# Multi-ancestry Genome Wide Association Study Meta-analysis of Non-syndromic Orofacial Clefts

**DOI:** 10.1101/2024.12.06.24318522

**Authors:** Zhonglin Jia, Nandita Mukhopadhyay, Zhenglin Yang, Azeez Butali, Jialin Sun, Yue You, Meilin Yao, Qi Zhen, Jian Ma, Miao He, Yongchu Pan, Azeez Alade, Yirui Wang, Mojisola Olujitan, Mengchun Qi, Wasiu Lanre Adeyemo, Carmen J. Buxó, Lord J.J Gowans, Mekonen Eshete, Yongqing Huang, Chenghao Li, Elizabeth J. Leslie, Lin Wang, Zhuan Bian, Jenna C. Carlson, Bing Shi, Seth M. Weinberg, Jeffrey C. Murray, Liangdan Sun, Mary L. Marazita, Rachel M. Freathy, Robin N. Beaumont

## Abstract

Non-syndromic orofacial clefts (NSOC) are common craniofacial birth defects, and result from both genetic and environmental factors. NSOC include three major sub-phenotypes: non-syndromic cleft lip with palate (NSCLP), non-syndromic cleft lip only (NSCLO) and non-syndromic cleft palate only (NSCPO), NSCLP and NSCLO are also sometimes grouped as non-syndromic cleft lip with or without cleft palate (NSCL/P) based on epidemiology. Currently known loci only explain a limited proportion of the heritability of NSOC. Further, differences in genetic susceptibility among the sub-phenotypes are poorly characterized. We performed a multi-ancestry GWAS meta-analysis on 44,094 individuals (9,381 cases, 28,510 controls, 2042 case-parent trios and 18 multiplex pedigrees) of East Asian, European, Latin and South American, and African ancestry for both NSOC and subtypes. We identified 50 loci, including 11 novel loci: four loci (*CALD1*, *SHH*, *NRG1* and *LINC00320*) associated with both NSOC and NSCL/P, two loci (*NTRK1* and *RUNX1*) only associated with NSOC, four loci (*HMGCR*, *PRICKLE1*, *SOX9* and *MYH9*) only associated with NSCL/P and one locus (*ALX1*) specifically associated with NSCLO. Five of the novel loci are located in regions containing genes associated with syndromic orofacial clefts (*SHH*, *NTRK1, CALD1, ALX1* and *SOX9*); seven of the novel loci are located in regions containing genes-implicated in craniofacial development (*HMGCR, SHH, PRICKLE1, ALX1, SOX9, RUNX1, MYH9*). Genetic correlation and colocalization analyses revealed an overlap between signals associated with NSCLO, NSCPO and NSCLP, but there were also notable differences, emphasizing the complexity of common and distinct genetic processes affecting lip and palate development.

## INTRODUCTION

Non-syndromic orofacial clefts (NSOC) are the most common disruption of normal facial structure, with an average incidence of 1/700 live births worldwide^1^. Prevalence of NSOC varies among different populations, with the highest prevalence in Asian or Amerindian populations [1/500], European-derived populations have an intermediate prevalence [1/1000], while African-derived populations have the lowest prevalence [1/2500]^1^.

The treatment of NSOC requires long-term care by teams of doctors in surgery, speech, hearing, orthodontics and psychology. In addition to its impact in early life, NSOC is associated with a lifetime increase in death from all causes as well as an increased risk for mental health conditions and cancer^2^. It imposes substantial economic burdens with expenses of more than $200,000 (US) lifetime per case and has a substantial impact on quality of life^3^.

NSOC is considered a complex trait and the non-syndromic (NS) refers to an absence of other associated structural or major cognitive abnormalities. It can be divided into three major sub-phenotypes: non-syndromic cleft lip with palate (NSCLP), non-syndromic cleft lip only (NSCLO) and non-syndromic cleft palate only (NSCPO). NSCLP and NSCLO are also sometimes grouped as non-syndromic cleft lip with or without cleft palate (NSCL/P) based on epidemiology (Supplementary Fig.1). Their phenotypic severity may be regulated by multiple genes and/or influenced by environmental triggers. Fogh-andersen et al. 1942 first proposed that genetics was a leading factor in the occurrence of NSOC^4^; twin and family studies of orofacial clefts consistently show higher concordance rates in monozygotic twins compared to dizygotic twins, and monozygotic twins had the concordance rate yielding estimates of heritability of 91% for NSOC^5^, which supports a major genetic component to its etiology.

Genome wide association studies (GWAS) and meta-analyses of GWASs have enabled progress in understanding the etiology of NSOC and its sub-phenotypes (NSCL/P, NSCLP, NSCLO and NSCPO) (Supplementary Fig.1), with 81 susceptibility loci reported^6–20^ (Supplementary Table 1). A few additional loci were first identified through linkage or candidate gene association studies and replicated independently^21,22^. However, current findings explain only a limited proportion of the heritability^23^. In addition, most previous studies used mixed samples of different subtypes of NSOC (NSCPO, NSCLO or NSCLP) to generate more power to detect variants shared across phenotypes, but might have missed true association signals for the specific sub-phenotypes as these could be diluted by other subtypes not at risk for those associated variants.

To uncover more NSOC susceptibility loci, advance current understanding of the genetic architecture and identify genetic components of the subphenotypes NSCLP, NSCLO and NSCPO, this study assembled 44,094 individuals (9381 cases, 28,510 controls and 6,203 individuals from 2042 case-parent trios and 18 multiplex pedigrees) across Asian, European, Latin and South American, and African populations (Supplementary Table 2), grouping cases into one shared (NSOC) and four subphenotypic groups (NSCL/P, NSCLP, NSCLO and NSCPO) (Supplementary Fig.1). We performed multi-ancestry genome wide association meta-analysis, genetic correlation and co-localization analysis, investigated the genetic basis of the phenotypic heterogeneity of NSOC, and investigated the potential predictive power of a polygenic risk score for NSOC (Fig.1).

**Fig. 1.**
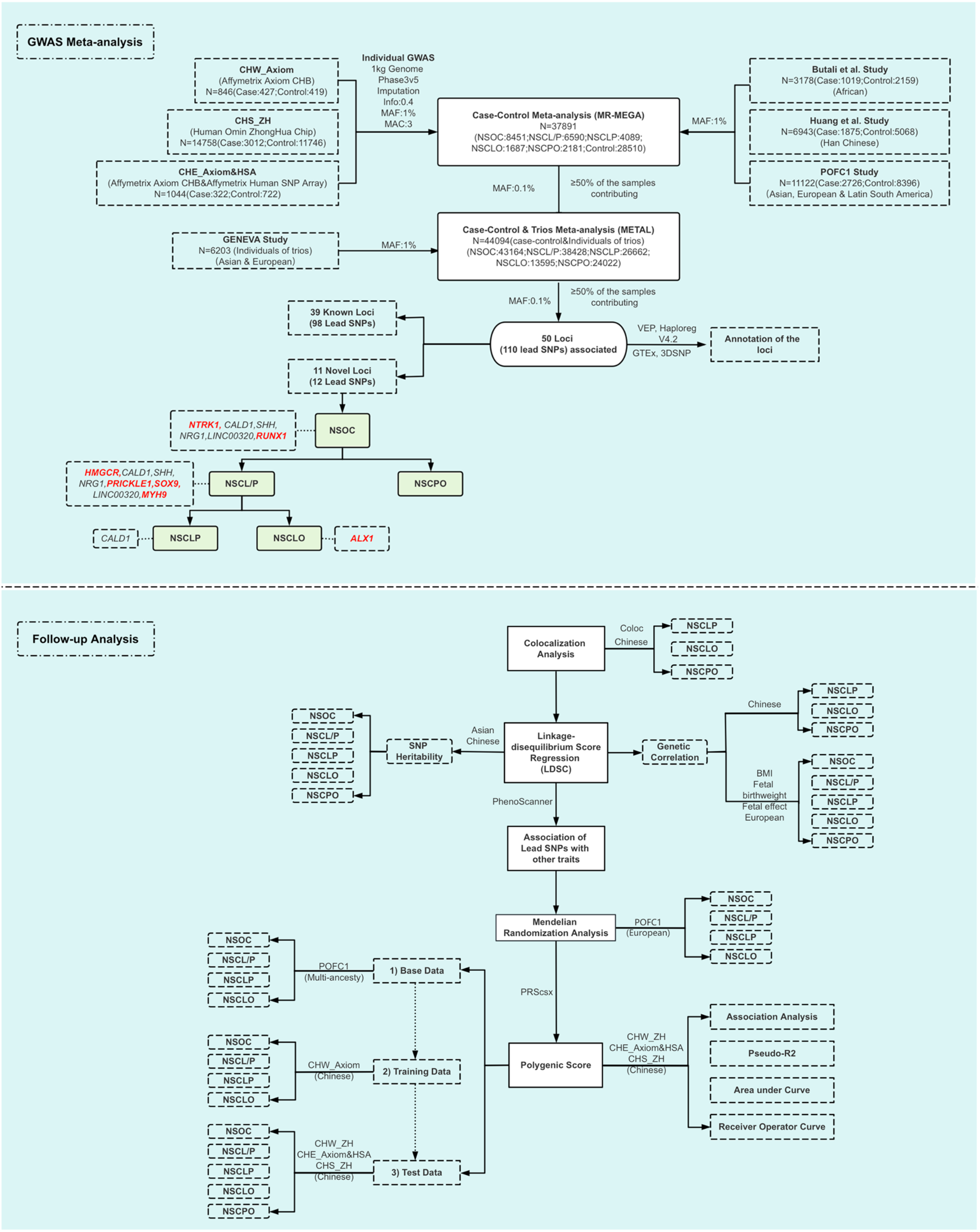
Flow Chart of the Multi-ancestry GWAS Meta-analysis

## RESULTS

### Multi-Ancestry Genome Wide Meta-analysis

We performed genome wide association (GWA) meta-analyses of seven independent datasets in three phases (Fig.1): In phase 1, we performed individual genome wide association studies in three independent datasets (CHW_Axiom, CHE_Axiom&HSA and CHS_ZH) of NSOC, NSCL/P NSCLP, NSCLO and NSCPO from a Han Chinese Population (Methods and Supplementary Tables 3 provide cohort information, data collection and genotyping details). After data cleaning and imputation, 8 million SNPs were analyzed. In phase 2, we performed GWA meta-analysis using the MR-MEGA software^24^ combining the case-control summary statistical results from Phase 1 with three independent previously published studies from Han Chinese (Huang et al. study)^14^, a mixed ancestry study (POFC1 study)^10^, and an African ancestry study^13^. In phase 3, we combined the filtered phase 2 results (MAF ≥0.1% and ≥50% of the samples contributing) with results from the GENEVA study (case-parent trio)^7^, in a p-value based meta-analysis using Metal software^25^. After quality control (Methods), to define independent lead SNPs we performed the clumping procedure implemented in the PLINK software^26^ based on the LD structure of 1000 genome phase 3v5 with default parameters. We defined novel loci as those in which the lead SNPs mapped >1 Mb away from the 81 loci reported at P<5.0E-08 in previous GWAS. Where loci fell within 1Mb of previously identified cleft locus which includes causal genes (syndromic orofacial cleft), known susceptibility genes (non-syndromic orofacial cleft) or genes that play roles during the craniofacial development, locus names were given to the cleft locus, otherwise the locus was named by the gene nearest to the lead SNP (Methods). We identified 50 independently associated loci (110 distinct lead SNPs; 152 SNP-trait associations) at genome-wide significant level (*P* < 5 × 10^−8^) (Fig.1, Fig.2, Table 1, Supplementary Table 4).

**Fig. 2.**
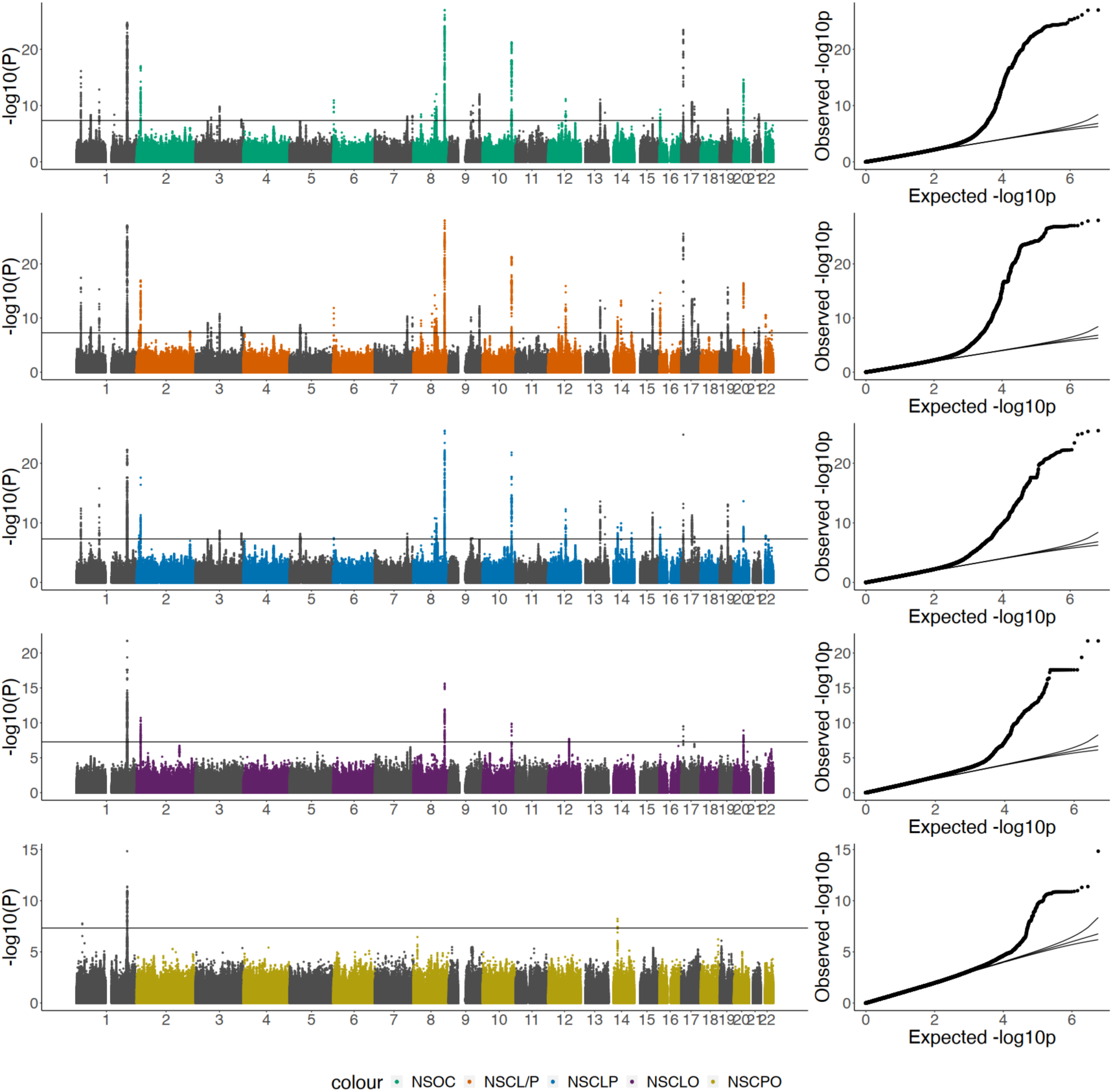
Manhattan and QQ Plot of the Multi-ancestry GWAS Meta-analysis Results NSOC, non-syndromic orofacial cleft cases by combining NSCLP,NSCLO and NSCPO together; NSCL/P, non-syndromic cleft lip with or without palate (NSCLP&NSCLO); NSCLP, non-syndromic cleft lip and palate; NSCLO, non-syndromic cleft lip only; NSCPO, non-syndromic cleft palate only.

**Table 1.**
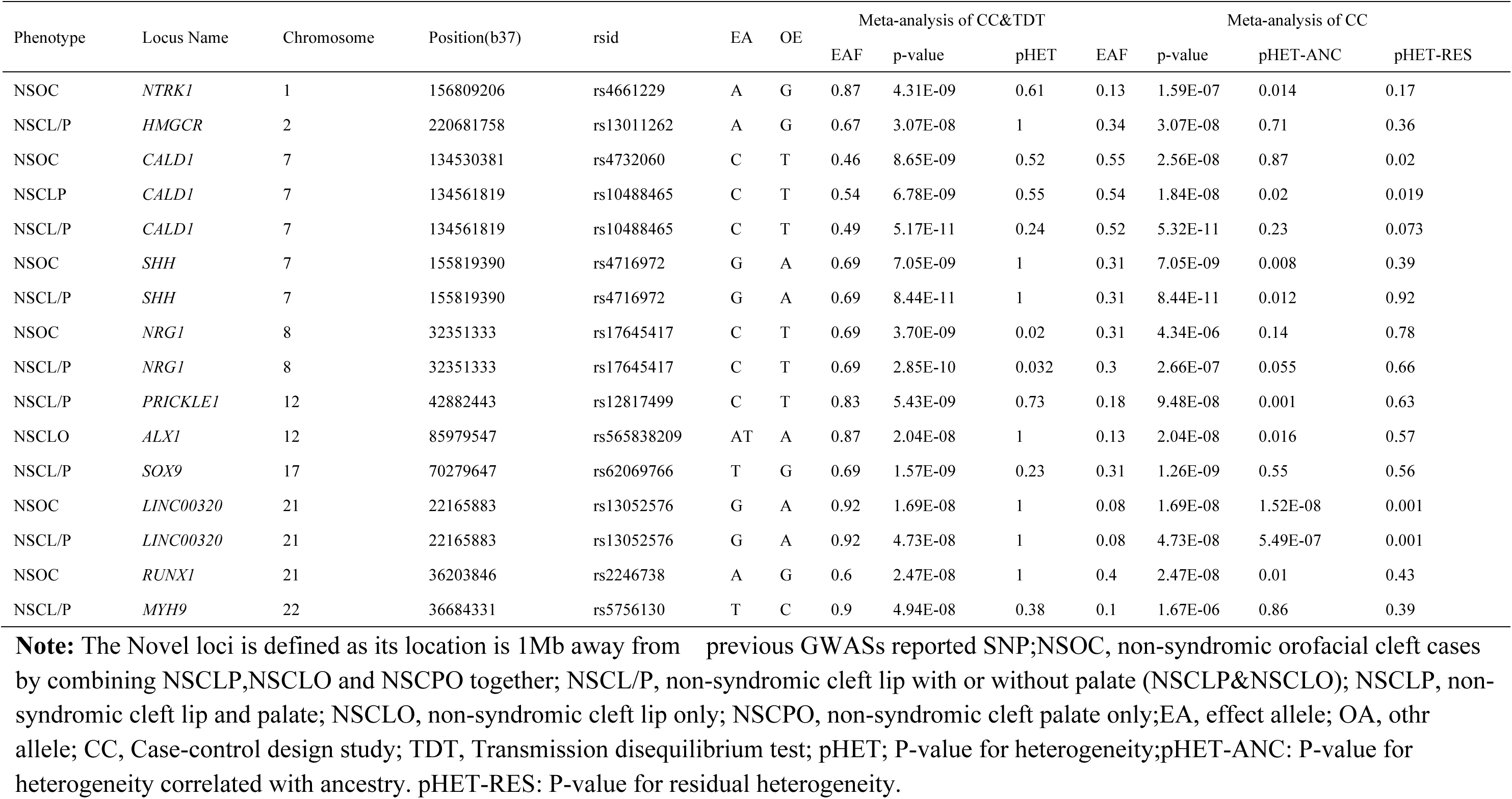
Summary of the lead SNPs in Novel Loci by Cleft Group.

We did not observe any substantial evidence of heterogeneity in any of the five phenotypic groups of the case-control meta-analysis [all lead SNPs have pHET-RES > 0.00045 (0.05/110)], However, six loci (five known and one novel locus) showed significant heterogeneity between ancestry groups in the case-control meta-analysis (pHET-ANC < 0.00045) (Table 1, Supplementary Table 4). Two known loci (two lead SNPs) showed significant heterogeneity in the final meta-analysis of the case-control and the GENEVA trio studies (pHET < 0.00045) (Supplementary Table 4).

### Functional annotation highlights links between loci identified here and known syndromic cleft genes

Of the 50 loci identified in this analysis, 39 loci were within 1Mb of SNPs previously shown to associate with NSOC (including 17 known SNPs and 81 newly identified SNPs) (Supplementary Table1, Supplementary Table 4). Eleven loci (12 SNPs) mapped >1 Mb away from the 81 loci reported at P<5.0E-08 in previous GWAS (Supplementary Table1, Table 1). Twelve of the 50 loci were only associated with one of the 5 NSOC phenotypes: three loci *(NTRK1*, *RIPK2* and *RUNX1*) associated with NSOC, six loci (*HMGCR*, *HYAL2*, *PRICKLE1*, *SOX9*, *MYH9* and *ARHGAP8*) with NSCL/P, one locus (*PTCH1*) with NSCLP, one locus (*ALX1)* with NSCLO, and one locus (*GRHL3*) with NSCPO (Table 1, Fig.1, Supplementary Table 4).

To explore the potential functions of the lead SNPs, we annotated them by using the Variant Effect Predictor (VEP) online tool from the Ensembl browser^27^, HaploReg v4.2^28^, GTEx database^29^ and 3DSNP^30^ (Fig.1). The majority of the lead SNPs were located in non-coding regions of genes or intergenic regions (Supplementary Table 5, Supplementary Table 6). A total of 63 lead SNPs (36 loci) were located in regulatory regions, 87 lead SNPs (46 loci) fall within transcription factor binding sites, 59 lead SNPs (31 loci) are expression quantitative trait loci (eQTL) and 28 lead SNPs (14 loci) are splicing quantitative trait loci (sQTL) (Supplementary Table 6). Four of the novel loci are located in regions containing genes associated with syndromic orofacial clefts [*NTRK1*^31^*, CALD1*^32^*, ALX1*^33^ and *SOX9*^34^ and seven of the novel loci are located in regions containing genes implicated in craniofacial development [*HMGCR*^35^*, SHH*^36^*, PRICKLE1*^37^*, ALX1*^38^*, SOX9*^39^*, RUNX1*^40^*, MYH9*^41^ (Supplementary Table 7).

One novel locus is of particular note, *CALD1*, which is located within the region known to cause syndromic orofacial clefts. Individuals with a maternally inherited 2.08 Mb deletion at chromosomal region 7q33 (chr7:133,176,651–135,252,871, hg19) containing 15 genes (including *CALD1*) have facial dysmorphism, including a narrow cleft palate and brachydactyly^32^. Both of rs4732060 and rs10488465 in the *CALD1* locus are eQTLs associated with expression levels of *CALD1*, *RP11-134L10.1*, and *C7orf49*, and are also sQTLs associated with splicing of *CALD1*, and *TMEM140* (supplementary Table 6). Of note the lead SNP rs4732060 is located within an enhancer region whose 3D interacting gene is *CALD1* in three cell lines (NHEK, epidermal keratinocytes), HepG2 (hepatocellular carcinoma) and HMEC (mammary epithelial cells) predicted by 3DSNP online software^30^(Fig.3a,Fig.3b,Fig.3c), to assess whether the rs4732060 affects the activity of its regulatory element, we conducted dual luciferase reporter assays, the results confirmed that the fragments containing rs4732060 function as enhancers, with the risk allele C significantly reduced enhancer function, whereas the non-risk allele T demonstrating strong enhancer activity, (P < 0.001) (Fig.3d). Expression of *CALD1* in several tissues (eg. Skin, Adipose et al.) with the C/C genotype of rs4732060 was significantly lower than in those with the T/T genotype (Fig.3e, Fig.3f). SNP rs4732060 is located within a region known to be bound by 14 transcription factors (e.g. Sox2, Pou2f2) (Supplementary Table 6, Fig.3c).

### Colocalization analyses between NSCLP, NSCLO and NSCPO

The *IRF6* locus is a known locus for both syndromic and NSOC, and in our analysis there were 11 SNPs associated with NSCLP, NSCLO, and/or NSCPO. In an attempt to elucidate the genetic architecture of this locus with these phenotypes, we performed colocalization analysis to determine the extent of overlap between our association signals for each trait (Fig.1). We found strong evidence for a shared association between NSCLP and NSCLO (PP.H4.abf=87.3%), and between NSCLP and NSCPO (PP.H4.abf=100%), while NSCLO showed evidence for distinct association signals from NSCPO (PP.H3.abf=77%) (Table 2, Supplementary Fig.2).

**Table 2.** Colocalisation Analysis on *IRF6* locus (chr1: 209800000-210670000, hg19) in Chinese.

| Traits_pairs | PP.H0.abf | PP.H1.abf | PP.H2.abf | PP.H3.abf | PP.H4.abf | PP abf for shared variant |
| --- | --- | --- | --- | --- | --- | --- |
| NSCLO_NSCPO | 1.82E-37 | 0.00% | 0.00% | 77.00% | 23.00% | 23.00% |
| NSCPO_NSCLP | 4.70E-47 | 0.00% | 0.00% | 0.05% | 100.00% | 100.00% |
| NSCLO_NSCLP | 8.48E-56 | 0.00% | 0.00% | 12.70% | 87.30% | 87.30% |

**Note:** NSCLO, non-syndromic cleft lip only; NSCPO, non-syndromic cleft palate only; NSCLP, non-syndromic cleft lip and palate; PP: posterior probabilities; H0: neither trait has a genetic association in the region; H1: only trait 1 has a genetic association in the region; H2: only trait 2 has a genetic association in the region; H3: both traits are associated, but with different causal variants; H4: both traits are associated and share a single causal variant.

### Correlations between Orofacial Clefts and BMI

To measure the proportion of phenotypic variance explained by all measured SNPs and to understand the degree to which common genetic variation influences these phenotypes (Fig.1), we estimated the SNP heritability by using the linkage-disequilibrium score regression (LDSC) method^42^ in Han Chinese and Asians separately. In Han Chinese, we found substantial heritability estimates in NSOC (*h*^2^ = 0.14, [95%CI: 0.11-0.18]), NSCL/P (*h*^2^ =0.21, [0.16-0.27]), NSCLP (*h*^2^ = 0.19, [0.14-0.25]), NSCLO (*h*^2^ = 0.25, [0.12-0.38]) and NSCPO (*h*^2^ = 0.26, [0.19-0.33]). The h^2^ estimates in Asians showed a similar trend (Fig.4a) (Supplementary Table 8).

We next estimated the genetic correlation between NSOC sub-phenotypes (NSCLO, NSCPO, and NSCLP) (Supplementary Fig.1) in the Chinese population to investigate how much of the genetic etiology may be shared across these phenotypes (Fig.1). NSCLO showed strong positive genetic correlations with NSCLP (*R*_g_ = 0.71, *P* = 1.92E-09). NSCLO and NSCPO showed weak positive genetic correlations (*R*_g_ =0.20, *P* = 0.38), while NSCLP and NSCPO showed weak inverse genetic correlations (*R*_g_ = −0.05, *P* = 0.65) (Fig.4b, Supplementary Table 9). These findings are similar to previous candidate gene studies, which found that NSCLP and NSCLO shared more susceptible loci and reflect the impact of shared genetic variants that influence both^14^.

There is an increased risk of having an offspring with an orofacial cleft in obese women^43,44^; fetuses with oral clefts are at elevated risk of having low and very low birth weight, and infants with low birth weight are more likely to have non-syndromic orofacial clefts^45^. To explore the extent to which known phenotypic association between maternal BMI and NSOC may be captured through shared genetic etiology, we estimated the genetic correlation of NSOC and its subtypes (NSCL/P, NSCLP, NSCLO and NSCPO) (Supplementary Fig.1), with BMI, the fetal GWAS of birth weight, and the fetal GWAS of birth weight conditional on maternal genotype using LDSC in a European ancestry Population (Fig.1). We found positive genetic correlation estimates between BMI and NSOC (*R*_g_ = 0.079, *P* = 0.030, 95%CI: 0.0076-0.15), NSCL/P (*R*_g_ = 0.077, *P* = 0.033, 95%CI:0.006-0.15) and NSCLP (*R*_g_ = 0.072, *P* = 0.04, 95%CI:0.0032-0.14). We found no evidence that NSOC or its sub-phenotypes are significantly correlated with fetal birth weight (either with or without conditioning on maternal genotype) (P>0.05) (Fig.4c, Supplementary Table 9).

### Associations of Lead SNPs with Other Traits

We performed a phenome-wide association analysis on our 110 lead SNPs using the online tool PhenoScanner^46^ to identify other phenotypes associated with our lead SNPs or those in LD (r^2^>0.8) (Fig.1). We found 34 lead SNPs with associations in PhenoScanner. As expected, 12 lead SNPs at 9 known loci had previous associations with orofacial clefts; in addition, we found 11 lead SNPs (7 loci) significantly associated with anthropometric traits (including BMI, weight, and height), 10 lead SNPs (5 loci) associated with blood cell traits, 3 lead SNPs associated with blood pressure (2 loci), 3 lead SNPs (3 loci) associated with bone mineral density, and 5 lead SNPs (3 loci) associated with other traits (supplementary Table 10). In addition to those mentioned above, several of our novel loci were also associated with normal-range human facial shape; e.g. the *ALX1* locus was associated with upper lip shape, and the *SOX9* locus was associated with nose shape ^47^.

### Mendelian Randomization Analysis

To test the effect of maternal BMI on risk of fetal orofacial clefts (NSOC, NSCL/P, NSCLP and NSCLO) we performed Two-sample Mendelian randomization (MR) analyses (Methods) (Supplementary Fig.3a) using 76 BMI-associated variants as instrumental variables (N_BMI_=339,224, N_cleft_=2,882). These results show weak but directionally consistent evidence that maternal genetic predisposition to a higher BMI may raise the risk of fetal orofacial cleft in NSOC (OR=1.59, 95%CI:0.37-6.74), NSCL/P group (OR=1.76, 95%CI:0.35-8.79), NSCLP (OR=1.68, 95%CI:0.30-9.23), and NSCLO (OR=1.18, 95%CI:0.12-11.93), although confidence intervals were wide and included the null. For NSOC and NSCL/P, the Weighted Median method showed some evidence of a causal effect, but the confidence intervals for these were very wide. Overall, while these results are consistent with the observational association, our results are inconclusive and require investigation in better powered analyses (Supplementary Table 11 and Supplementary Fig.3b, Supplementary Fig.3c).

### Polygenic Score

To investigate predictive/discriminative ability explained by the results from the present study, we aggregated thousands of SNP effects simultaneously into polygenic score (PGS) and assessed its association with NSOC and sub-phenotypes’ risk in two steps (Methods) (Supplementary Table 12, Fig.1). The PGS was associated strongly with NSOC, NSCL/P, NSCLP, NSCLO, and NSCPO (all P<0.05; Supplementary Table 13, Fig.5a). To indicate the predictive ability and the proportion of phenotypic variance explained by the polygenic scores (PGSs), we calculated the coefficient of determination R^2^ (pseudo-*R*^2^) by building two prediction models (with/without PGS). We found a relatively modest increase in R^2^ values in models including PGS indicating that the likely utility of PGS in prediction of NSOC is low with current summary statistics (Supplementary Table 13).

To further measure the discrimination of PGS in disease prediction, we calculated the area under the receiver-operator curve (AUC) with or without covariate sex in the logistic regression model adjusting for 10 genetic principal components (PCs). PGS achieves a best prediction accuracy with an AUC of 0.57 (95%CI:0.56-0.58) for NSOC, 0.61 (0.58-0.65) for NSCL/P, 0.61 (0.59-0.62) for NSCLP, 0.63 (0.57-0.68) for NSCLO, and 0.54 (0.51-0.56) for NSCPO. When adding the sex covariate in the model, the AUC value increased in both the CHW_ZH and CHS_ZH datasets, and increased substantially in the CHE_Axiom&HSA dataset which is a result of the biased distributions of sex in cases (Male: 214, Female:108) and controls (Male: 5, Female: 717) (Fig.5b, Fig.5c, Fig.5d, Supplementary Table 13).

## DISCUSSION

In our multi-ancestry meta-analysis of NSOC and its sub-phenotypes we identified 110 lead association signals at 50 genetic loci. This includes 12 lead SNPs at 11 novel loci, and 81 newly identified SNPs overlapping with known NSOC loci (Fig.1, Table 1, Supplementary Table 4). Compared with the other phenotypic subgroups, we found fewer association signals for NSCLO and NSCPO due to smaller sample sizes for these subphenotypes. In addition, the majority of the signals for NSCPO showed significant ancestry-based heterogeneity (Supplementary Table 4).

Among our 11 novel loci, 9 contain genes either implicated in syndromic orofacial clefts (*NTRK1, CALD1, ALX1, SHH* and *SOX9*), or genes involved in aspects of craniofacial development (*HMGCR, SHH, PRICKLE1, ALX1, SOX9, RUNX1, MYH9*), where abnormal expression can lead to an orofacial cleft (Supplementary Table 7).

Functional annotation showed that 102 out of 110 independent SNPs fall within transcription factor binding sites, regulatory regions, eQTLs or sQTLs (Supplementary Table 5, Supplementary Table 6,); of note among them are two lead SNPs within the *CALD1* locus. Both are eQTLs, and one (rs4732060) is located within an enhancer whose 3D interacting gene is *CALD1* in three cell lines (NHEK, epidermal keratinocytes), HepG2 (hepatocellular carcinoma) and HMEC (mammary epithelial cells) predicted by the 3DSNP software ^30^. Functional work confirmed that rs4732060 is located in an enhancer, with the risk allele C significantly reducing enhancer activity, whereas the non-risk allele T demonstrates strong enhancer function (P < 0.001) (Fig. 3d); expression of *CALD1* in several tissues (eg. skin, adipose et al.) with the C/C genotype of rs4732060 was significantly lower than in those with the T/T genotype (Fig.3e, Fig.3f). *CALD1* is also located within a region known to be bound by several transcription factors including Sox2 and, Pou2f2. Previous studies have demonstrated that deletion of *SOX2* in oral epithelium disrupts palatal shelf extension^48^. In addition, *POU2F2*, also known as *OCT2*, encodes a transcription factor and is differentially expressed in early embryogenesis^49^. It is involved in cellular proliferation and differentiation of various cell types^50–52^, suggesting that it may influence the morphogenetic pathways that shape the developing face. *CALD1*, Caldesmon 1, is a protein-coding gene, which encodes calmodulin and actin-binding proteins and plays an important role in the regulation of smooth muscle and non-muscle contraction. CALD1-related disorders include adenomyomas and cellular leiomyomas, associated with pathways including ERK development, angiotensin activation, and cardiac conduction. Additionally, patients with 7q33 CNV are reported to have an abnormal palatal phenotype, involving the duplication or deletion of *CALD1*^32^. *CALD1* overexpression is a key component in TGF-β-driven epithelial-mesenchymal transition (EMT)^53^; TGF-β signaling and TGF-β induced EMT processes are critical for the palate development, and more specifically for completing the process of palatal fusion^54,55^. In addition, EMT is vital in embryonic development, including craniofacial morphogenesis, where epithelial cells transform into mesenchymal cells to facilitate tissue formation. Abnormalities in EMT can lead to defects such as cleft lip and palate^56^. These results indicate that rs4732060 is a functional SNP, and support a role for it in the etiology of orofacial clefts by affecting *CALD1* expression.

**Fig. 3.**
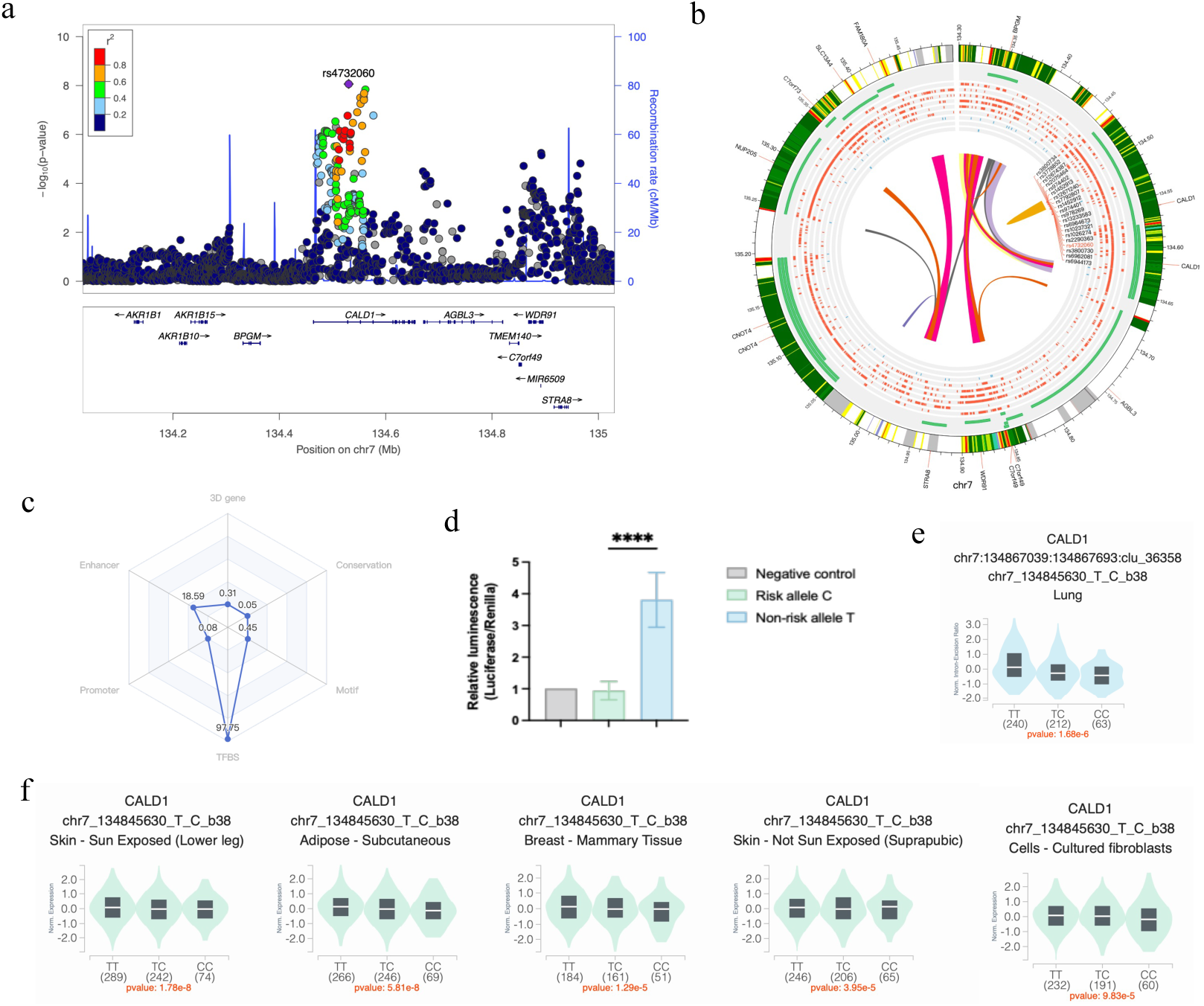
a, Regional LD plot of CALD1 locus; b, CIRCOS plot based on NHEK cell line, Circos tracks from outside to inside represent: Chromatin states, RefSeq genes, DHS and histone modifications, TFBS, associated SNPs and chromatin loops. Histone Modification makers: H3K27ac, H3K27me3, H3k36me, H3K4me1, H3K4me3, H3K9me3; Transcription Factor: CTCF,EZH2,POLR2A. c, radar chart showing the distribution of the six functionality scores for this SNP, the total score of functionality for this SNP is 117.24, 3D interacting genes: This SNP is linked to 2 genes via chromatin loops, the corresponding score is 0.31; Enhancer: It locates in Enhancer state in 32 cell types, the corresponding score is 18.59.TFBS: It locates in 77 transcription factor binding sites, the corresponding score is 97.75; Conservation: Its PhyloP score is -1.064, the corresponding score is 0.05; d, Dual-luciferase reporter assay of PGL3 promoter plasmids containing enhancer fragments with different rs4732060 alleles transfected into human GMSM-K cells. Data are normalized to Renilla luciferase activity (n = 8); risk allele (OR>1) was determined by performing the meta-analysis of the case control based GWASs with Metal software. e, Single-Tissue sQTLs for rs4732060; f,Single-Tissue eQTLs for rs4732060.

**Fig. 4.**
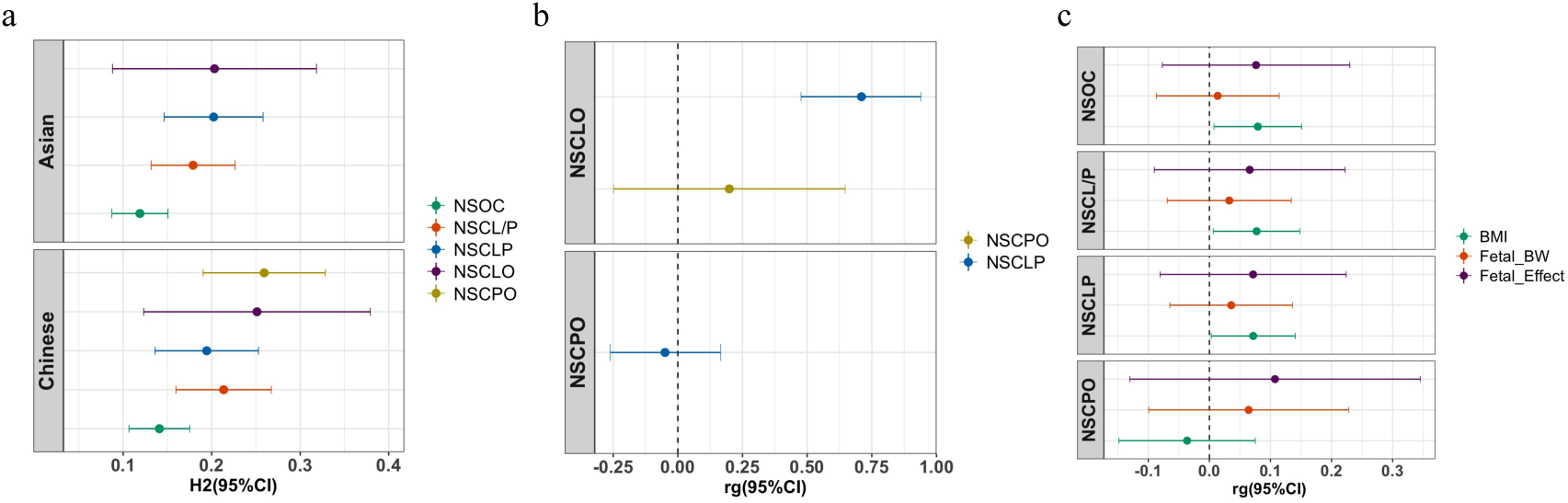
Heritability Estimates and Genetic Correlation a, SNP Heritability Estimated by phenotypic group in Asian and Chinese;b, Genetic Correlation among sub-phenotypes in Chinese;c, Genetic Correlation between orofacial clefts and adiposity phenotypes in European

Palate and lip originate from different tissues during embryonic development, so NSCLO and NSCPO are commonly regarded as two independent disorders supported by their different epidemiology. NSCLO and NSCLP however, are regarded as related phenotypes since they both involve lip deformities^57^ (Supplementary Fig.1). However, many studies have showed that there is a more complex relationship among NSCLO, NSCPO and NSCLP, with shared etiologies between the three, but also with their own unique features^58^. To further elucidate the extent of shared etiology within the largest risk locus, this study performed colocalization on the *IRF6* locus. As expected, we found high evidence for shared associations between NSCLO and NSCLP, with evidence for distinct association signals for NSCLO and NSCPO. There was also evidence that NSCLP shared association signals with NSCPO, although due to the muti-ancestry nature of our meta-analysis we were unable to implement the colocalization methods which allow for multiple causal variants at a single locus. Genetic correlations based on linkage-disequilibrium score regression further confirmed that NSCLO (in Han Chinese samples) has stronger positive genetic correlations with NSCLP (*R*_g_ = 0.71) than that with NSCPO (*R*_g_ =0.20). These findings further clarify the genetic background between NSCLO, NSCPO and NSCLP, reflecting the impact of shared genetic variants that influence risk of both NSCLO and NSCLP, but also the presence of distinct susceptibility genes for both traits which drive the different genetic heritability between the two traits with NSCPO higher (*h*^2^ = *0.26*) than NSCLP (*h*^2^ = 0.19) (Fig.4a, Supplementary Table 8).

Numerous epidemiological studies have reported associations between maternal BMI and orofacial clefts with obese mothers having an elevated risk for children with orofacial clefts when compared to normal weight mothers^43,44^. Obese mothers were at significantly increased odds of a pregnancy affected by either cleft palate or cleft lip and palate compared with mothers with BMI within the normal range; cleft lip showed weak evidence for association with maternal obesity^59^. Our MR analyses found weak but directionally consistent evidence for a causal role of higher maternal BMI in risk of fetal orofacial cleft consistent with estimates from the literature^60,61^. The large uncertainty in our estimates mean that further evidence is needed to confirm the association.

To investigate the discriminative power of a PGS for NSOC, we trained a PGS for NSCL/P using the multi-ancestry cohorts of POFC1 (Methods, Supplementary Table 12). We found that this PGS was significantly associated with the risk of NSOC sub-phenotypes (NSCL/P, NSCLP, NSCLO and NSCPO) (Supplementary Table 13, Fig.5a). AUC values of NSOC, NSCL/P, NSCLP, NSCLO and NSCPO ranges between 0.54 and 0.63 indicating that while the PGS provides some discriminative power, at present it does not reach a level which may be useful for clinical use (Supplementary Fig.1) (Fig. 5b, Fig. 5c, Fig. 5d, Supplementary Table 13).

**Fig. 5.**
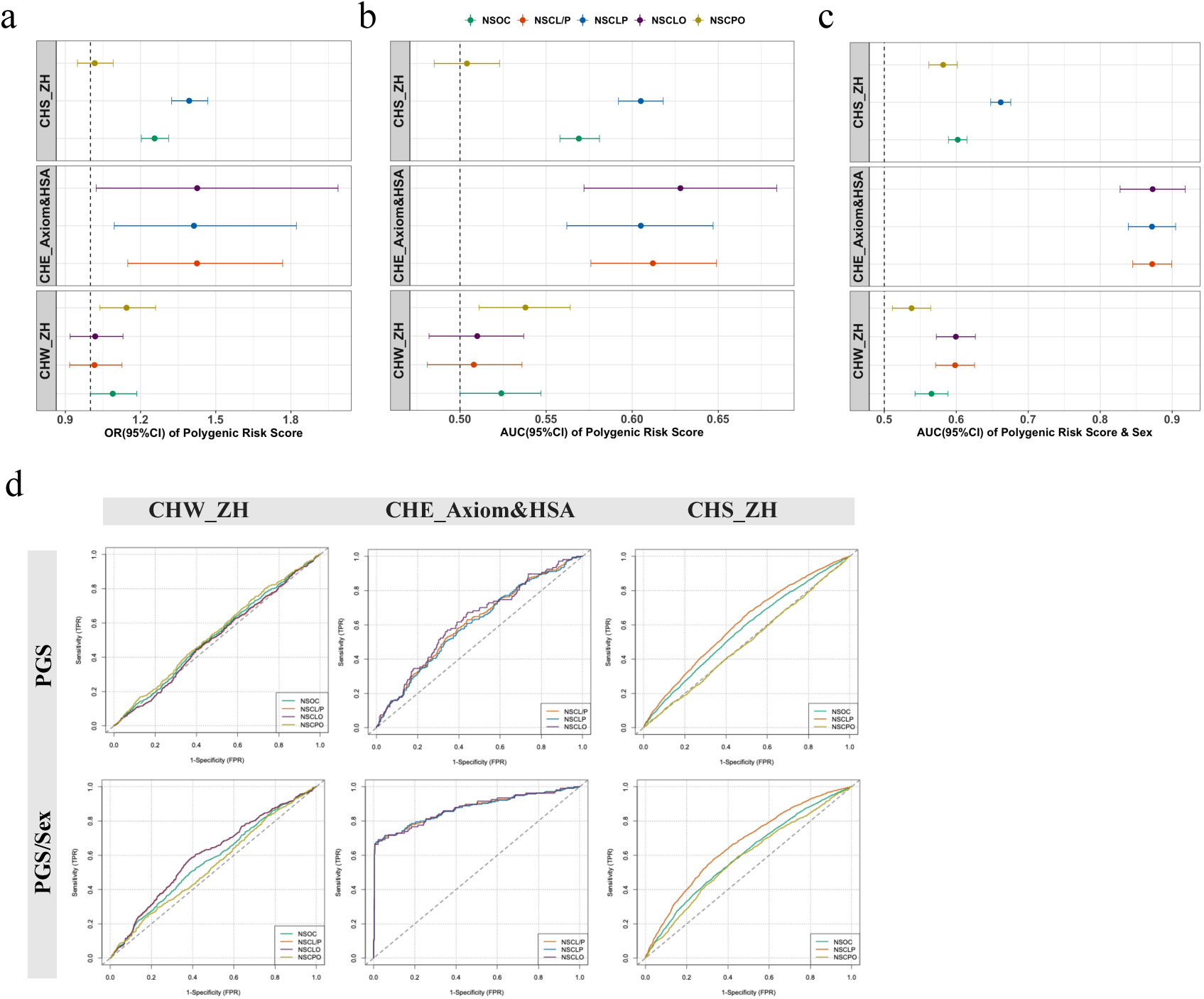
Forest plot of Associations between PGS and Non-syndromic orofacial clefts in Han Chinese Cohorts a, odds ratio (95%CI); b, AUC (area under the curve) and 95%CI of PGS; c, AUC and 95%CI of PGS &SEX; CHS_ZH in figure c is calculated among the individuals with SEX information; d, Receiver Operator Characteristic Curves for Different Models.

There are some limitations to this study. First, the colocalization analysis was performed under a single causal variant assumption so was only able to estimate the probability of colocalization of the strongest association signal for each trait. Methods which can extend the functionality by allowing more than one causal variant exist but due to the multi-ancestry nature of our analyses we were not able to fit these with reliability due to their reliance on accurate maps of linkage-disequilibrium. Second, the sample size is not big enough to test associations between the PGS and sub-phenotypes of NSOC. Third, the sample sizes are relatively small to test the relationship between maternal BMI and fetal orofacial clefts by performing MR analysis.

Our study complements large-scale GWA studies of orofacial clefts. While genetic correlation and colocalization analyses revealed an overlap between signals associated with NSCLO, NSCPO and NSCLP, there were also notable differences emphasizing the complexity of common and distinct genetic regulation and physiological processes affecting lip and palate development. Overall, our findings provide an improved understanding of the role of maternal BMI in orofacial clefts. Future research may focus on additional proxies for orofacial clefts and environmental influences, as well as the role of orofacial clefts for later-life health outcomes.

## Data Availability

All results produced in the present study are available upon reasonable request to the authors

## ACKNOWLEDGEMENTS

The authors thank all the participants who donated samples in this study. This project was supported by the National Science Funds of China (No.81271118, No. 81600849), Sichuan Province Science and Technology support program (No.2024NSFSC0649), the Research and Develop Program, West China Hospital of Stomatology Sichuan University (No. RD-03-202301); R.M.F. was supported by a Wellcome Senior Fellowship (WT220390). This research was funded in part, by the Wellcome Trust [Grant number WT220390]. For the purpose of open access, the authors have applied a CC BY public copyright licence to any author accepted manuscript version arising from this submission; Liangdan Sun was supported by Hebei Natural Science Foundation H2023209084 and S&T Program of Hebei 23372505D; Miao He was supported by the Joint Funds of the National Natural Science Foundation of China (No. U22A20313); Yongchu Pan was supported by the National Natural Science Foundation of China (82270496) and Key Research and Development Program of Jiangsu Province (BE2023831); Zhuan Bian was supported by the National Natural Science Foundation of China (No.82170944, No.81970923, No.82370966); The POFC1 cohorts were recruited and genotyped through grant support from the United States National Institutes of Health (NIH): grant numbers R01-DE016148, R01-DE008559, X01-HG000784. The GENEVA cohorts were genotyped through NIH grant support: grant number U01-DE-018993.

## METHODS

### Study Cohorts and Ethical Approval

Our studied enrolled 44,094 individuals (9381 cases, 28,510 controls, and 6,203 individuals of 2042 trios and 18 multiplex pedigrees multiplex pedigrees) across Asian, European, Latin South American, and African populations from 7 studies (Fig.1 and Supplementary Table 1): CHW_Axiom study including 846 individuals (427 cases and 419 Control), CHE_Axiom&HSA study including 1044 individuals (322 cases and 722 controls), CHS_ZH study including 14758 individuals (3012 cases and 11179 controls) and Huang et al. Study including 6943 individuals (1875 cases and 5068 controls) are from Han Chinese population^14^; Butali et al. study included 3178 individuals (1019 cases and 2159 normal controls) from African population^13^; the Marazita/Weinberg et al Pittsburgh Orofacial Cleft (POFC1) study included 11122 individuals (2726 cases and 8396 controls)^10^; the Beaty Johns Hopkins University GENEVA studies included 6203 individuals from 2042 trios and 18 multiplex pedigrees multiplex pedigrees ^7^. Study protocols were approved at each study center by the local ethics committee and written informed consent was obtained from all participants and/or their parent(s) or legal guardians. Study descriptions and phenotype information of samples are presented in Supplementary Table 1.

### Individual GWAS analysis

Genotypes in each study of phase 1 were obtained through high-density SNP arrays and up to ∼0.8 million autosomal SNPs were imputed to 1000 genome phase 3v5. Ancestry principal components for studies except the POFC1 study were calculated by flashPCA package^61^ and were included as covariables where necessary in the individual studies. Genome-wide association analyses of CHW_Axiom study and CHE_Axiom&HSA study were conducted using SNPTEST software^62^ adjusted by using sex and 10 PCs as covariates, and Genome-wide association analyses of CHS_ZH study was performed by regenie software^63^ using 10 PCs as covariates. SNPs with MAF (Minor allele frequency) ≥1% and info ≥ 0.4 of each study were included for the following case-control meta-analysis. For the POFC1 study, ancestry principal components calculated using the KING software^64^ were used to create four ancestry-based sample subsets. African, Asian, European and Central and South American. Genome-wide association analysis was conducted separately within each ancestry-subset, using the R GENESIS association package, utilizing a genetic relationship matrix (GRM) to account for familial relatedness.

### Meta-analysis Methods

Meta-analyses were performed in two steps based on the study design. Firstly, we conducted meta-analysis on case-control design studies by MR-MEGA (Meta-Regression of Multi-Ethnic Genetic Association) software^24^. SNPs with MAF ≥0.1% and ≥50% of the samples contributing were taken forward to the second stage, where we performed p-value based meta-analysis with case-parent trios from the POFC1^10^ and GENEVA studies^7^ by Metal software^25^. SNPs with MAF ≥0.1% and ≥50% of the samples contributing were selected to perform clump analysis to extract the independent lead SNPs by PLINK software^26^ based on the LD structure of 1000 genome phase 3v5 with default parameters. SNPs mapping >1 Mb away from the previously reported GWAS loci were defined as novel loci. Where the loci fell within 1Mb of genes known to cause syndromic orofacial cleft, known susceptibility gene for non-syndromic orofacial cleft, or playing roles during the craniofacial development, these genes were used as locus names, otherwise the locus was named by the nearest gene. The Manhattan plots and QQ plots by using R software ggplot2 package. Regional LD plots were generated by LocusZoom (hg19 Nov 2014 ASN Population)^65^.

### Dual-luciferase reporter assays

For the dual luciferase reporter assays, GMSM-K cells were co-transfected with PGL3 promoter plasmids containing different rs4732060 allele-specific enhancer fragments as transcriptional activators, along with Renilla luciferase as a normalization control. Whole-cell lysates were collected 48 hours post-transfection, and luciferase activity was measured using the Dual-Luciferase Reporter Assay Kit (Vazyme, DL101) following the manufacturer’s protocol.

### Colocalization Analysis

We performed colocalization analysis on *IRF6* locus in Chinese (using summary statistics of meta-analysis from Chinese population) to distinguish the genetic structure of NSCLP, NSCLO and NSCPO using the R library ‘coloc’ version 5.1.0 with the default prior probability for colocalization^66^. Signals were defined as colocalising if the posterior probability of shared association signals (P4) was >0.8, distinct if the posterior probability of independent signals (P3) was >0.8 and undetermined if neither P3 or P4 was >0.8.

### LD Score Regression

LD score regression^42^ was used, to calculate the SNP based heritability adjusted by sample prevalence and worldwide population prevalence of 0.1% for NSOC and its sub-phenotypes (NSCL/P, NSCLP, NSCLO and NSCPO) in Chinese and Asian population (using summary statistics of meta-analysis from Chinese and Asian population); to estimate the genetic correlation between NSCLP, NSCLO and NSCPO in Chinese; and genetic correlation between BMI^67^, Fetal birth weight^68^ and Fetal effect^68^ with orofacial clefts in a European population (POFC1).

### Associations of Lead SNPs with Other Traits

We looked at associations between our 110 independent NSOC (including its sub-phenotype groups) associated SNPs and other phenotypes within the phenoscanner^46^. We looked for associations with the SNP itself, and SNPs in LD (*r*^2^ ≥ 0.8).

### Mendelian Randomization Analysis

We performed two-sample mendelian randomization (MR) analyses with maternal BMI as the exposure, and risk of offspring orofacial clefts (NSOC, NSCL/P, NSCLP and NSCLO) as outcomes. The exposures used maternal BMI using 76 genetic variants previously associated with BMI as instruments from previous reported GWA study of BMI^69^ (N= 339,224). The SNP-orofacial clefts associations were taken from POFC1 GWASs in European population (N=2,882), and adjusted for fetal genotype (N=11,122) using WLM-adjusted analysis to account for confounding due to correlation between maternal and fetal genotype^70^. To obtain SNP estimates for offspring orofacial clefts conditional on offspring genotype, we first transformed the log ORs of maternal and fetal to the liability scale^71^, and then applied the WLM-adjusted analysis. The fetal orofacial cleft-independent maternal BMI effects were then transformed to log ORs for analysis. A population prevalence for orofacial clefts of 0.1% was used for the transformations. We applied the inverse-variance weighted MR method, with MR-Egger^72^, weighted median and penalized weighted median^73^ acting as sensitivity analysis to test for robustness to MR assumptions.

### Polygenic Scores

Step1, we calculated the base on POFC1 GWAS summary results of European, Latin/South American and Asian populations on four phenotypic groups (NSOC, NSCL/P, NSCLP and NSCLO)^10^ by PRScsx software^74^. Step2, the CHW_Axiom dataset (NSOC, NSCL/P, NSCLP and NSCLO) was used as training data to assess the most powerful base among the four phenotypic group of POFC1 GWAS, by comparing the association results of the training datasets, the NSCL/P of POFC1 derived PGS had the strongest association. Step3, we tested the evidence for association between PGS and NSOC and its sub-phenotypic groups in three independent datasets (CHW_ZH, CHE_Axiom&HSA and CHS_ZH), and evaluated its potential effect and estimated the effect size (OR and 95%CI) per unit (normalized of PGS which is calculated by the formula [PRS-mean(PRS)]/SD(PRS)).

To indicate the predictive ability and the proportion of phenotypic variance explained by the polygenic scores (PGSs). We calculated the coefficient of determination R^2^ (pseudo-*R*^2^) by building two prediction logistic regression models (with/without PGS) and calculated by R rms Package.

To further measure the discrimination of PGS in disease prediction, we calculated area under the curve (AUC) and plotted the receiver-operator curve (ROC) with or without covariates sex in the logistic regression model adjusting for 10 PCs by R ROCit Package.

## REFERENCE

1. Dixon, M.J., Marazita, M.L., Beaty, T.H. & Murray, J.C. Cleft lip and palate: understanding genetic and environmental influences. Nat Rev Genet 12, 167–78 (2011).

2. Christensen, K., Juel, K., Herskind, A.M. & Murray, J.C. Long term follow up study of survival associated with cleft lip and palate at birth. BMJ 328, 1405 (2004).

3. Wehby, G.L. & Cassell, C.H. The impact of orofacial clefts on quality of life and healthcare use and costs. Oral Dis 16, 3–10 (2010).

4. Inheritance of Harelip and Cleft Palate: Contribution to the Elucidation of the Etiology of the Congenital Clefts of the Face. Journal of the American Medical Association 133, 276–276 (1947).

5. Grosen, D. et al. Risk of oral clefts in twins. Epidemiology 22, 313–9 (2011).

6. Birnbaum, S. et al. Key susceptibility locus for nonsyndromic cleft lip with or without cleft palate on chromosome 8q24. Nat Genet 41, 473–7 (2009).

7. Beaty, T.H. et al. A genome-wide association study of cleft lip with and without cleft palate identifies risk variants near MAFB and ABCA4 (vol 42, pg 525, 2010). Nature Genetics 42, 727–727 (2010).

8. Ludwig, K.U. et al. Genome-wide meta-analyses of nonsyndromic cleft lip with or without cleft palate identify six new risk loci. Nat Genet 44, 968–71 (2012).

9. Sun, Y. et al. Genome-wide association study identifies a new susceptibility locus for cleft lip with or without a cleft palate. Nat Commun 6, 6414 (2015).

10. Leslie, E. et al. A multi-ethnic genome-wide association study identifies novel loci for non-syndromic cleft lip with or without cleft palate on 2p24.2, 17q23 and 19q13. Hum Mol Genet 25, 2862–2872 (2016).

11. Leslie, E.J. et al. Genome-wide meta-analyses of nonsyndromic orofacial clefts identify novel associations between FOXE1 and all orofacial clefts, and TP63 and cleft lip with or without cleft palate. Hum Genet 136, 275–286 (2017).

12. Yu, Y. et al. Genome-wide analyses of non-syndromic cleft lip with palate identify 14 novel loci and genetic heterogeneity. Nat Commun 8, 14364 (2017).

13. Butali, A. et al. Genomic analyses in African populations identify novel risk loci for cleft palate. Hum Mol Genet 28, 1038–1051 (2019).

14. Huang, L. et al. Genetic factors define CPO and CLO subtypes of nonsyndromicorofacial cleft. PLoS Genet 15, e1008357 (2019).

15. He, M. et al. Genome-wide Analyses Identify a Novel Risk Locus for Nonsyndromic Cleft Palate. J Dent Res 99, 1461–1468 (2020).

16. Mukhopadhyay, N. et al. Genome-Wide Association Study of Non-syndromic Orofacial Clefts in a Multiethnic Sample of Families and Controls Identifies Novel Regions. Frontiers in Cell and Developmental Biology 9, 13 (2021).

17. Avasthi, K.K., Muthuswamy, S., Asim, A., Agarwal, A. & Agarwal, S. Identification of Novel Genomic Variations in Susceptibility to Nonsyndromic Cleft Lip and Palate Patients. Pediatr Rep 13, 650–657 (2021).

18. Li, B. et al. Genome-wide analyses of nonsyndromic cleft lip with or without palate identify 20 new risk loci in the Chinese Han population. J Genet Genomics 49, 903–905 (2022).

19. Robinson, K. et al. Trio-based GWAS identifies novel associations and subtype-specific risk factors for cleft palate. HGG Adv 4, 100234 (2023).

20. Yu, Y. et al. Genome-wide meta-analyses identify five new risk loci for nonsyndromic orofacial clefts in the Chinese Han population. Mol Genet Genomic Med 11, e2226 (2023).

21. Zucchero, T.M. et al. Interferon regulatory factor 6 (IRF6) gene variants and the risk of isolated cleft lip or palate. N Engl J Med 351, 769–80 (2004).

22. Lidral, A.C. et al. Association of MSX1 and TGFB3 with nonsyndromic clefting in humans. Am J Hum Genet 63, 557–68 (1998).

23. Génin, E. Missing heritability of complex diseases: case solved? Hum Genet 139, 103–113 (2020).

24. Mägi, R. et al. Trans-ethnic meta-regression of genome-wide association studies accounting for ancestry increases power for discovery and improves fine-mapping resolution. Hum Mol Genet 26, 3639–3650 (2017).

25. Willer, C.J., Li, Y. & Abecasis, G.R. METAL: fast and efficient meta-analysis of genomewide association scans. Bioinformatics 26, 2190–1 (2010).

26. Purcell, S. et al. PLINK: a tool set for whole-genome association and population-based linkage analyses. Am J Hum Genet 81, 559–75 (2007).

27. McLaren, W. et al. The Ensembl Variant Effect Predictor. Genome Biol 17, 122 (2016).

28. Ward, L.D. & Kellis, M. HaploReg: a resource for exploring chromatin states, conservation, and regulatory motif alterations within sets of genetically linked variants. Nucleic Acids Res 40, D930–4 (2012).

29. The Genotype-Tissue Expression (GTEx) project. Nat Genet 45, 580-5 (2013).

30. Quan, C., Ping, J., Lu, H., Zhou, G. & Lu, Y. 3DSNP 2.0: update and expansion of the noncoding genomic variant annotation database. Nucleic Acids Res 50, D950–d955 (2022).

31. Aleksiūnienė, B., Preiksaitiene, E., Morkūnienė, A., Ambrozaitytė, L. & Utkus, A. A de novo 1q22q23.1 Interstitial Microdeletion in a Girl with Intellectual Disability and Multiple Congenital Anomalies Including Congenital Heart Defect. Cytogenet Genome Res 154, 6–11 (2018).

32. Lopes, F. et al. The contribution of 7q33 copy number variations for intellectual disability. Neurogenetics 19, 27–40 (2018).

33. Uz, E. et al. Disruption of ALX1 causes extreme microphthalmia and severe facial clefting: expanding the spectrum of autosomal-recessive ALX-related frontonasal dysplasia. Am J Hum Genet 86, 789–96 (2010).

34. Benko, S. et al. Highly conserved non-coding elements on either side of SOX9 associated with Pierre Robin sequence. Nat Genet 41, 359–64 (2009).

35. Signore, I.A. et al. Inhibition of the 3-hydroxy-3-methyl-glutaryl-CoA reductase diminishes the survival and size of chondrocytes during orofacial morphogenesis in zebrafish. Orthod Craniofac Res 26, 378–386 (2023).

36. Kurosaka, H., Iulianella, A., Williams, T. & Trainor, P.A. Disrupting hedgehog and WNT signaling interactions promotes cleft lip pathogenesis. Journal of Clinical Investigation 124, 1660–1671 (2014).

37. Yang, T. et al. Analysis of PRICKLE1 in human cleft palate and mouse development demonstrates rare and common variants involved in human malformations. Mol Genet Genomic Med 2, 138–51 (2014).

38. Nguyen, T.T. et al. TFAP2 paralogs regulate midfacial development in part through a conserved ALX genetic pathway. Development 151(2024).

39. Bi, W. et al. Haploinsufficiency of Sox9 results in defective cartilage primordia and premature skeletal mineralization. Proc Natl Acad Sci U S A 98, 6698–703 (2001).

40. Yamashiro, T., Kurosaka, H. & Inubush, T. The Association Between Runx Signaling and Craniofacial Development and Disease. Curr Osteoporos Rep 20, 120–126 (2022).

41. Martinelli, M. et al. Cleft lip with or without cleft palate: implication of the heavy chain of non-muscle myosin IIA. J Med Genet 44, 387–92 (2007).

42. Bulik-Sullivan, B.K. et al. LD Score regression distinguishes confounding from polygenicity in genome-wide association studies. Nat Genet 47, 291–5 (2015).

43. Izedonmwen, O.M., Cunningham, C. & Macfarlane, T.V. What is the Risk of Having Offspring with Cleft Lip/Palate in Pre-Maternal Obese/Overweight Women When Compared to Pre-Maternal Normal Weight Women? A Systematic Review and Meta-Analysis. J Oral Maxillofac Res 6, e1 (2015).

44. Wehby, G.L. et al. Interaction between smoking and body mass index and risk of oral clefts. Ann Epidemiol 27, 103–107.e2 (2017).

45. Wyszynski, D.F., Sarkozi, A., Vargha, P. & Czeizel, A.E. Birth weight and gestational age of newborns with cleft lip with or without cleft palate and with isolated cleft palate. J Clin Pediatr Dent 27, 185–90 (2003).

46. Kamat, M.A. et al. PhenoScanner V2: an expanded tool for searching human genotype-phenotype associations. Bioinformatics 35, 4851–4853 (2019).

47. White, J.D, et al. Insights into the genetic architecture of the human face. Nat Genet 53, 45–53 (2021).

48. Sweat, Y.Y. et al. Sox2 Controls Periderm and Rugae Development to Inhibit Oral Adhesions. J Dent Res 99, 1397–1405 (2020).

49. Schöler, H.R., Hatzopoulos, A.K., Balling, R., Suzuki, N. & Gruss, P. A family of octamer-specific proteins present during mouse embryogenesis: evidence for germline-specific expression of an Oct factor. Embo j 8, 2543–50 (1989).

50. Doane, A.S. et al. OCT2 pre-positioning facilitates cell fate transition and chromatin architecture changes in humoral immunity. Nat Immunol 22, 1327–1340 (2021).

51. García-Caballero, D., Hart, J.R. & Vogt, P.K. The MYC-regulated lncRNA LNROP (ENSG00000254887) enables MYC-driven cell proliferation by controlling the expression of OCT2. Cell Death Dis 14, 168 (2023).

52. Hodson, D.J. et al. Regulation of normal B-cell differentiation and malignant B-cell survival by OCT2. Proc Natl Acad Sci U S A 113, E2039–46 (2016).

53. Morita, T., Mayanagi, T. & Sobue, K. Dual roles of myocardin-related transcription factors in epithelial mesenchymal transition via slug induction and actin remodeling. J Cell Biol 179, 1027–42 (2007).

54. Gritli-Linde, A. Molecular control of secondary palate development. Dev Biol 301, 309–26 (2007).

55. Jalali, A., Zhu, X., Liu, C. & Nawshad, A. Induction of palate epithelial mesenchymal transition by transforming growth factor β3 signaling. Dev Growth Differ 54, 633–48 (2012).

56. Lu, Y. et al. Mutations of GADD45G in rabbits cause cleft lip by the disorder of proliferation, apoptosis and epithelial-mesenchymal transition (EMT). Biochim Biophys Acta Mol Basis Dis 1865, 2356–2367 (2019).

57. Marazita, M.L. The Evolution of Human Genetic Studies of Cleft Lip and Cleft Palate. in Annual Review of Genomics and Human Genetics, Vol 13, Vol. 13 (eds. Chakravarti, A. & Green, E.) 263–283 (Annual Reviews, Palo Alto, 2012).

58. Carlson, J.C. et al. A systematic genetic analysis and visualization of phenotypic heterogeneity among orofacial cleft GWAS signals. Genetic Epidemiology 43, 704–716 (2019).

59. Stothard, K.J., Tennant, P.W., Bell, R. & Rankin, J. Maternal overweight and obesity and the risk of congenital anomalies: a systematic review and meta-analysis. Jama 301, 636–50 (2009).

60. Kutbi, H. et al. Maternal underweight and obesity and risk of orofacial clefts in a large international consortium of population-based studies. Int J Epidemiol 46, 190–199 (2017).

61. Francisco, I., Caramelo, F., Fernandes, M.H. & Vale, F. Parental Risk Factors and Child Birth Data in a Matched Year and Sex Group Cleft Population: A Case-Control Study. Int J Environ Res Public Health 18(2021).

## REFERENCE

61. Abraham, G., Qiu, Y. & Inouye, M. FlashPCA2: principal component analysis of Biobank-scale genotype datasets. Bioinformatics 33, 2776–2778 (2017).

62. Marchini, J., Howie, B., Myers, S., McVean, G. & Donnelly, P. A new multipoint method for genome-wide association studies by imputation of genotypes. Nat Genet 39, 906–13 (2007).

63. Mbatchou, J. et al. Computationally efficient whole-genome regression for quantitative and binary traits. Nat Genet 53, 1097–1103 (2021).

64. Manichaikul, A. et al. Robust relationship inference in genome-wide association studies. Bioinformatics 26, 2867–73 (2010).

65. Pruim, R.J. et al. LocusZoom: regional visualization of genome-wide association scan results. Bioinformatics 26, 2336–7 (2010).

66. Giambartolomei, C. et al. Bayesian test for colocalisation between pairs of genetic association studies using summary statistics. PLoS Genet 10, e1004383 (2014).

67. Yengo, L. et al. Meta-analysis of genome-wide association studies for height and body mass index in ∼700000 individuals of European ancestry. Hum Mol Genet 27, 3641–3649 (2018).

68. Warrington, N.M. et al. Maternal and fetal genetic effects on birth weight and their relevance to cardio-metabolic risk factors. Nat Genet 51, 804–814 (2019).

69. Locke, A.E. et al. Genetic studies of body mass index yield new insights for obesity biology. Nature 518, 197–206 (2015).

70. Beaumont RN., Genome-wide association study of placental weight identifies distinct and shared genetic influences between placental and fetal growth. Nat Genet 55,1807–1819 (2023)

71. Gillett, A.C., Vassos, E. & Lewis, C.M. Transforming Summary Statistics from Logistic Regression to the Liability Scale: Application to Genetic and Environmental Risk Scores. Hum Hered 83, 210–224 (2018).

72. Bowden, J., Davey Smith, G. & Burgess, S. Mendelian randomization with invalid instruments: effect estimation and bias detection through Egger regression. Int J Epidemiol 44, 512–25 (2015).

73. Bowden, J., Davey Smith, G., Haycock, P.C. & Burgess, S. Consistent Estimation in Mendelian Randomization with Some Invalid Instruments Using a Weighted Median Estimator. Genet Epidemiol 40, 304–14 (2016).

74. Ruan, Y. et al. Improving polygenic prediction in ancestrally diverse populations. Nat Genet 54, 573–580 (2022).

