## Supplementary Figures for "Multi-ancestry Genome Wide Association Study Meta-analysis of Non-syndromic Orofacial Clefts"

**ABSTRACT**

Non-syndromic orofacial clefts (NSOC) are common craniofacial birth defects, and result from both genetic and environmental factors. NSOC include three major sub-phenotypes: non-syndromic cleft lip with palate (NSCLP), non-syndromic cleft lip only (NSCLO) and non-syndromic cleft palate only (NSCPO), NSCLP and NSCLO are also sometimes grouped as non-syndromic cleft lip with or without cleft palate (NSCL/P) based on epidemiology. Currently known loci only explain a limited proportion of the heritability of NSOC. Further, differences in genetic susceptibility among the sub-phenotypes are poorly characterized. We performed a multi-ancestry GWAS meta-analysis on 44,094 individuals (9,381 cases, 28,510 controls, 2042 case-parent trios and 18 multiplex pedigrees) of East Asian, European, Latin and South American, and African ancestry for both NSOC and subtypes. We identified 50 loci, including 11 novel loci: four loci (*CALD1*, *SHH*, *NRG1* and *LINC00320*) associated with both NSOC and NSCL/P, two loci (*NTRK1* and *RUNX1*) only associated with NSOC, four loci (*HMGCR*, *PRICKLE1*, *SOX9* and *MYH9*) only associated with NSCL/P and one locus (*ALX1*) specifically associated with NSCLO. Five of the novel loci are located in regions containing genes associated with syndromic orofacial clefts (*SHH*, *NTRK1, CALD1, ALX1* and *SOX9*); seven of the novel loci are located in regions containing genes implicated in craniofacial development (*HMGCR, SHH, PRICKLE1, ALX1, SOX9, RUNX1, MYH9*). Genetic correlation and colocalization analyses revealed an overlap between signals associated with NSCLO, NSCPO and NSCLP, but there were also notable differences, emphasizing the complexity of common and distinct genetic processes affecting lip and palate development.

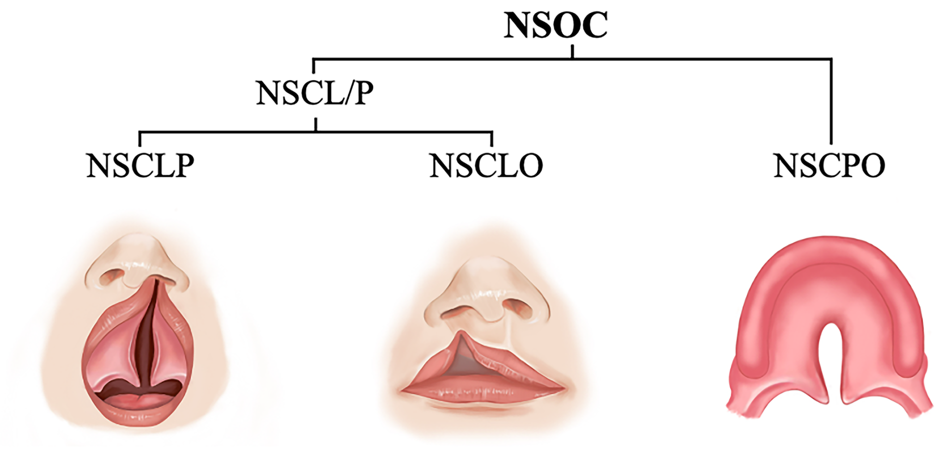


**Supplementary Fig.1** NSOC and its sub-phenotypes

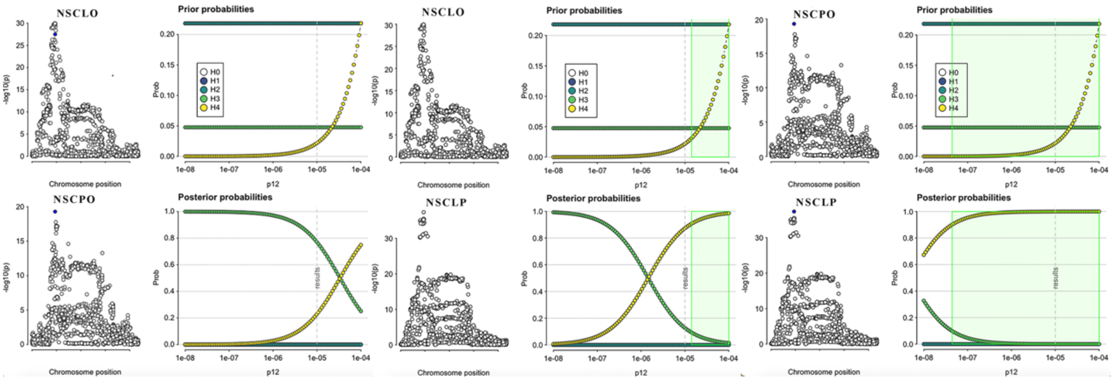


**Supplementary Fig. 2** Colocalization Analysis on IRF6 locus (Chr1: 209800000-210670000) in Chinese population

p12, Prior probability a random SNP in the region is jointly causal for both traits; The green region shows the region -the set of values of p12 for which H4>0.9 the rule that was specified. blue point on the left input data indicate the posterior probabilities that a SNP is causal if H4 were true.

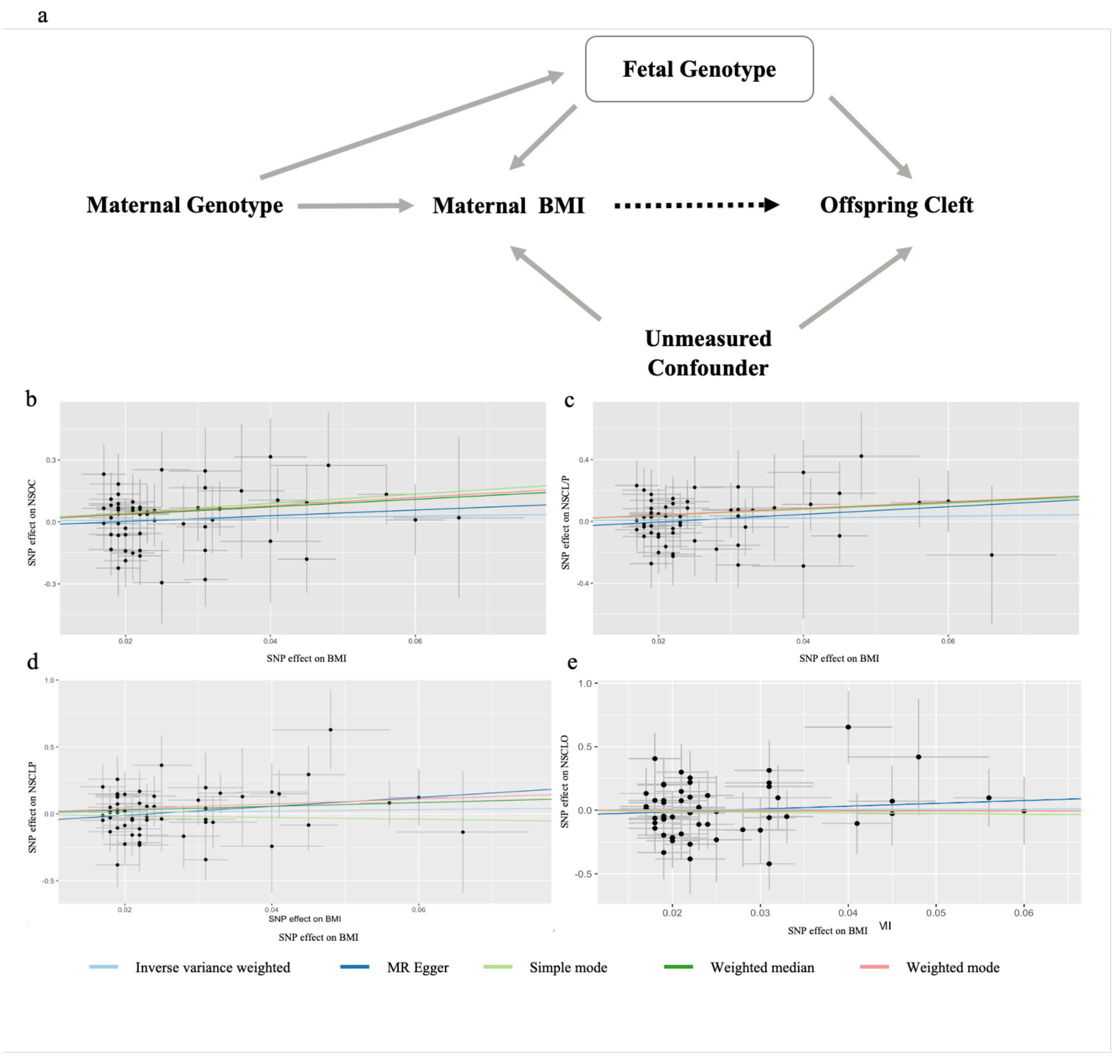


**Supplementary Fig.3** Mendelian randomization analyses

a, Diagram illustrating the Mendelian randomization analyses used to test for a causal; b–e, Results of two-sample Mendelian randomization analyses testing the effect of maternal BMI on fetal orofacial clefts. Points represent SNP effect estimates, and error bars show 95% confidence intervals.

NSOC, non-syndromic orofacial cleft cases by combining NSCLP, NSCLO and NSCPO together; NSCL/P, non-syndromic cleft lip with or without palate (NSCLP&NSCLO); NSCLP, non-syndromic cleft lip and palate; NSCLO, non-syndromic cleft lip only; NSCPO, non-syndromic cleft palate only.
