## Supplementary Tables for "Multi-ancestry Genome Wide Association Study Meta-analysis of Non-syndromic Orofacial Clefts"

ABSTRACT

Non-syndromic orofacial clefts (NSOC) are common craniofacial birth defects, and result from both genetic and environmental factors. NSOC include three major sub-phenotypes: non-syndromic cleft lip with palate (NSCLP), non-syndromic cleft lip only (NSCLO) and non-syndromic cleft palate only (NSCPO), NSCLP and NSCLO are also sometimes grouped as non-syndromic cleft lip with or without cleft palate (NSCL/P) based on epidemiology. Currently known loci only explain a limited proportion of the heritability of NSOC. Further, differences in genetic susceptibility among the sub-phenotypes are poorly characterized. We performed a multi-ancestry GWAS meta-analysis on 44,094 individuals (9,381 cases, 28,510 controls, 2042 case-parent trios and 18 multiplex pedigrees) of East Asian, European, Latin and South American, and African ancestry for both NSOC and subtypes. We identified 50 loci, including 11 novel loci: four loci (CALD1, SHH, NRG1 and LINC00320) associated with both NSOC and NSCL/P, two loci (NTRK1 and RUNX1) only associated with NSOC, four loci (HMGCR, PRICKLE1, SOX9 and MYH9) only associated with NSCL/P and one locus (ALX1) specifically associated with NSCLO. Five of the novel loci are located in regions containing genes associated with syndromic orofacial clefts (SHH, NTRK1, CALD1, ALX1 and SOX9); seven of the novel loci are located in regions containing genes implicated in craniofacial development (HMGCR, SHH, PRICKLE1, ALX1, SOX9, RUNX1, MYH9). Genetic correlation and colocalization analyses revealed an overlap between signals associated with

Supplementary Table 1. 81 Loci reported by previous GWASs

| No.Loci | Chromosome | Position (b37) | SNP | Gene | Associated Phen | Published GWAS | PMID | Data Multi-Ancestry GWAS Meta |
|---|---|---|---|---|---|---|---|---|
| 1 | 1 | 3047252 | rs2817178 | *PRDM16* | NSCL/P | Li et al.2022 | 35217232 | No significance |
| 2 | 1 | 18972776 | rs9439713 | *PAX7* | NSCL/P,NSOC | Leslie et al.2017 | 28054174 | Confirmed |
| 2 | 1 | 18976489 | rs9439714 | *PAX7* | NSCL/P | Mukhopadhyay et al.20 | 33898419 | Confirmed |
| 2 | 1 | 18978372 | rs4920524 | *PAX7* | NSCL/P | Leslie et al.2016 | 27033726 | Confirmed |
| 2 | 1 | 18979874 | rs742071 | *PAX7* | NSCL/P | Ludwig et al.2012 | 22863734 | Confirmed |
| 3 | 1 | 24669457 | rs41268753 | *GRHL3* | NSCPO | Leslie et al.2017;Leslie | 28054174;27033 | Confirmed |
| 4 | 1 | 59832057 | rs11588505 | *FGGY* | NSOC | Yu et al.2023 | 37326468 | Confirmed |
| 5 | 1 | 62673037 | rs7542665 | *L1TD1* | NSCL/P | Li et al.2022 | 35217232 | No significance |
| 5 | 1 | 64627002 | rs11208372 | *ROR1, UBE2U* | NSCL/P | Li et al.2022 | 35217232 | No significance |
| 6 | 1 | 94553438 | rs560426 | *ARHGAP29* | NSCL/P | Beaty et al.2010;Ludw | 20436469;228637 | Confirmed |
| 6 | 1 | 94558110 | rs66515264 | *ARHGAP29* | NSCL/P,NSOC | Leslie et al.2017 | 28054174;228637 | Confirmed |
| 6 | 1 | 94558256 | rs4147882 | *ARHGAP29* | NSCL/P | Leslie et al.2016 | 27033726 | Confirmed |
| 6 | 1 | 94570016 | rs481931 | *ARHGAP29* | NSCL/P | Beaty et al.2010;Li et a | 20436469;352172 | Confirmed |
| 6 | 1 | 94575056 | rs4147811 | *ARHGAP29* | NSCL/P | Beaty et al.2010 | 20436469 | Confirmed |
| 7 | 1 | 209947548 | rs11119345 | *IRF6* | NSCL/P | Leslie et al.2016 | 27033726 | Confirmed |
| 7 | 1 | 209962794 | rs2073485 | *IRF6* | NSCL/P | Beaty et al.2010 | 20436469 | Confirmed |
| 7 | 1 | 209964080 | rs2235371 | *IRF6* | NSCL/P,NSCLO | Sun et al.2015;Huang e | 25775280;31609 | Confirmed |
| 7 | 1 | 209966629 | rs17015217 | *IRF6* | NSCL/P | Mukhopadhyay et al.20 | 33898419 | Confirmed |
| 7 | 1 | 209968684 | rs2013162 | *IRF6* | NSCL/P | Beaty et al.2010 | 20436469 | Confirmed |
| 7 | 1 | 209975392 | rs2235377 | *IRF6* | NSCLO | Huang et al.2019 | 31609978 | Confirmed |
| 7 | 1 | 209977111 | rs861020 | *IRF6* | NSCL/P,NSCLP | Beaty et al.2010;Ludw | 20436469;228637 | Confirmed |
| 7 | 1 | 209977844 | rs12405750 | *IRF6* | NSCLO | Huang et al.2019 | 31609978 | Confirmed |
| 7 | 1 | 209984470 | rs75477785 | *IRF6* | NSCL/P | Leslie et al.2017* | 28054174 | Confirmed |
| 7 | 1 | 209988047 | rs10863790 | *IRF6* | NSCL/P,NSCLO | Beaty et al.2010;Sun et | 20436469;257752 | Confirmed |
| 7 | 1 | 209988527 | rs77249837 | *IRF6* | NSCL/P | Li et al.2022 | 35217232 | Confirmed |
| 7 | 1 | 209989092 | rs72741048 | *IRF6* | NSCLO | Huang et al.2019 | 31609978 | Confirmed |
| 7 | 1 | 209989270 | rs642961 | *IRF6* | NSCLP | Yu et al.2017 | 28232668 | Confirmed |
| 7 | 1 | 209992501 | rs1109430 | *IRF6* | NSOC | Leslie et al.2017* | 28054174 | Confirmed |
| 7 | 1 | 210048819 | rs2064163 | *IRF6* | NSCLP | Yu et al.2017 | 28232668 | Confirmed |
| 7 | 1 | 210211912 | rs189000 | *IRF6* | NSCLO | Huang et al.2019 | 31609978 | Confirmed |
| 7 | 1 | 210226819 | rs1387934 | *IRF6* | NSCLO | Huang et al.2019 | 31609978 | Confirmed |
| 8 | 2 | 9983008 | rs72782605 | *TAF1B* | NSCL/P | Li et al.2022 | 35217232 | Confirmed |
| 9 | 2 | 16710565 | rs4832468 | *FAM49A* | NSCLO | Huang et al.2019 | 31609978 | Confirmed |
| 9 | 2 | 16713395 | rs6745357 | *FAM49A* | NSCL/P | Mukhopadhyay et al.20 | 22863734 | Confirmed |

| | | | | | | | | |
|---|---|---|---|---|---|---|---|---|
| 9 | 2 | 16729357 | rs7566780 | *FAM49A* | NSCL/P,NSOC | Leslie et al.2017* | 28054174 | Confirmed |
| 9 | 2 | 16733054 | rs10172734 | *FAM49A* | NSCLP | Yu et al.2017 | 28232668 | Confirmed |
| 9 | 2 | 16733928 | rs7552 | *FAM49A* | NSCL/P,NSCLO | Leslie et al.2016;Huang | 27033726;316099 | Confirmed |
| 10 | 2 | 43540125 | rs7590268 | | NSCL/P | Ludwig et al.2012 | 22863734 | No significance |
| 11 | 2 | 45127854 | rs11887207 | *SIX3* | NSCL/P | Li et al.2022 | 35217232 | No significance |
| 12 | 2 | 82025185 | rs80004662 | *CTNNA2* | NSCPO | Butali et al.2019 | 30452639 | NA |
| 12 | 2 | 82028390 | rs113691307 | *CTNNA2* | NSCPO | Butali et al.2019 | 30452639 | NA |
| 13 | 2 | 130176577 | rs17491637 | - | NSCLP | Yu et al.2017 | 28232668 | No significance |
| 14 | 3 | 50344178 | rs78138837 | *HYAL1* | NSOC | Yu et al.2023 | 37326468 | Confirmed |
| 15 | 3 | 54903701 | rs2703028 | *CACNA2D3* | NSCL/P | Li et al.2022 | 35217232 | No significance |
| 16 | 3 | 56063735 | rs10601122 | *ERC2* | NSOC | Yu et al.2023 | 37326468 | No significance |
| 17 | 3 | 64625244 | rs61021131 | *ADAMTS9* | NSOC | Yu et al.2023 | 37326468 | Confirmed |
| 18 | 3 | 89534377 | rs7632427 | - | NSCL/P | Ludwig et al.2012 | 22863734 | No significance |
| 19 | 3 | 99711816 | rs9873878 | *FILIP1L* | NSCL/P | Li et al.2022 | 35217232 | Confirmed |
| 20 | 3 | 150092627 | rs1450341 | *TSC22D2* | NSCL/P | Li et al.2022 | 35217232 | No significance |
| 21 | 3 | 189553372 | rs76479869 | *TP63* | NSCL/P | Leslie et al.2017* | 28054174 | Confirmed |
| 22 | 3 | 196777238 | rs71161992 | *DLG1* | NSCL/P | Li et al.2022 | 35217232 | No significance |
| 23 | 4 | 4852849 | rs735510 | *MSX1* | NSCL/P | Li et al.2022 | 35217232 | No significance |
| 24 | 4 | 124807935 | rs17007553 | *LOC285419* | NSCL/P | Li et al.2022 | 35217232 | No significance |
| 25 | 5 | 44068846 | rs10462065 | *FGF10* | NSCL/P | Li et al.2022 | 35217232 | Confirmed |
| 26 | 5 | 73317553 | rs6453037 | | NSCPO | He et al.2020 | 32758111 | No significance |
| 27 | 5 | 132847845 | rs17166739 | | NSCPO | He et al.2020 | 32758111 | No significance |
| 28 | 6 | 1945408 | rs3800146 | *GMDS* | NSCL/P | Li et al.2022 | 35217232 | Confirmed |
| 29 | 6 | 9442330 | rs7771515 | *OFCC1* | NSCLP | Yu et al.2017 | 28232668 | No significance |
| 29 | 6 | 9469238 | rs9381107 | *OFCC1* | NSCLP | Yu et al.2017 | 28232668 | No significance |
| 30 | 6 | 162671456 | rs12175475 | *PARK2* | NSCPO | Leslie et al.2016 | 27033726 | NA |
| 31 | 8 | 8865923 | rs2979260 | *ERI1* | NSCL/P | Li et al.2022 | 35217232 | No significance |
| 32 | 8 | 77496113 | rs11993273 | *ZFHX4* | NSCL/P | Li et al.2022 | 35217232 | Confirmed |
| 33 | 8 | 88868340 | rs12543318 | *DCAF4L2* | NSCL/P/NSOC | Ludwig et al.2012;Lesl | 22863734;280541 | Confirmed |
| 34 | 8 | 90231591 | rs318271 | *RIPK2* | NSCL/P | Li et al.2022 | 35217232 | Confirmed |
| 35 | 8 | 95541302 | rs957448 | *VIRMA* | NSCLP | Yu et al.2017 | 28232668 | Confirmed |
| 35 | 8 | 95594720 | rs12682601 | *VIRMA* | NSCL/P | Li et al.2022 | 35217232 | Confirmed |
| 36 | 8 | 100369465 | rs3110405 | *STK3, OSR2, VF* | NSCL/P | Li et al.2022 | 35217232 | No significance |
| 37 | 8 | 129933720 | rs72728734 | *LINC00824* | NSCL/P | Leslie et al.2016 | 27033726 | Confirmed |
| 37 | 8 | 129946154 | rs987525 | *LINC00824* | NSCL/P | Birnbaum et al.2009;Beaty et al.2010;Ludwig et al.2012 | 19270707;20436469;22863734 | Confirmed |

| | | | | | | | | |
|---|---|---|---|---|---|---|---|---|
| 37 | 8 | 129951769 | rs7815993 | *LINC00824* | NSCL/P | Li et al.2022 | 35217232 | Confirmed |
| 37 | 8 | 129964873 | rs17242358 | *LINC00824* | NSOC | Leslie et al.2017* | 28054174 | Confirmed |
| 37 | 8 | 129976136 | rs55658222 | *LINC00824* | NSCL/P | Leslie et al.2016;Leslie | 27033726;280541 | Confirmed |
| 37 | 8 | 129990382 | rs72728755 | *LINC00824* | NSCL/P | Mukhopadhyay et al.2( | 33898419 | Confirmed |
| 38 | 9 | 89289528 | rs1331446 | *ZCCHC6* | NSCL/P | Li et al.2022 | 35217232 | No significance |
| 39 | 9 | 92212750 | rs1475537 | *GADD45G* | NSCL/P | Li et al.2022 | 35217232 | Confirmed |
| 40 | 9 | 98280788 | rs28469297 | *PTCH1* | NSCL/P | Li et al.2022 | 35217232 | Confirmed |
| 41 | 9 | 100619719 | rs12347191 | *FOXE1* | NSOC | Leslie et al.2017* | 28054174 | Confirmed |
| 42 | 9 | 127327763 | rs10114038 | *NR6A1* | NSCL/P | Li et al.2022 | 35217232 | Confirmed |
| 43 | 9 | 129944999 | rs7035976 | *ANGPTL2* | Cleft Hard Palate | Robinson et al.2023 | 37719664 | No significance |
| 44 | 10 | 29233160 | rs1832845 | *LOC10050760* | NSCL/P | Li et al.2022 | 35217232 | No significance |
| 45 | 10 | 99695194 | rs474558 | | NSCLP | Yu et al.2017 | 28232668 | No significance |
| 46 | 10 | 102657733 | rs3050320 | *FAM178A* | NSCL/P | Li et al.2022 | 35217232 | No significance |
| 47 | 10 | 118715399 | rs7902527 | *VAX1* | NSCLO | Huang et al.2019 | 31609978 | Confirmed |
| 47 | 10 | 118827560 | rs7078160 | *VAX1* | NSCL/P | Ludwig et al.2012 | 22863734 | Confirmed |
| 47 | 10 | 118834991 | rs4752028 | *VAX1* | NSCLO | Huang et al.2019 | 31609978 | Confirmed |
| 47 | 10 | 118846294 | rs10886040 | *VAX1* | NSCL/P,NSOC | Leslie et al.2017;Li et a | 28054174;352172 | Confirmed |
| 47 | 10 | 118893231 | rs6585429 | *VAX1* | NSCLP | Yu et al.2017 | 28232668 | Confirmed |
| 48 | 11 | 44843134 | rs11038167 | | NSCPO | He et al.2020 | 32758111 | No significance |
| 49 | 11 | 102079713 | rs117496742 | *YAP1* | NSCPO | Leslie et al.2016 | 27033726 | NA |
| 50 | 12 | 53357335 | rs2363632 | *KRT18* | NSCL/P | Li et al.2022 | 35217232 | No significance |
| 51 | 12 | 56384687 | rs705698 | *RAB5B* | NSCL/P | Li et al.2022 | 35217232 | No significance |
| 52 | 12 | 71962975 | rs10784927 | *LGR5* | NSCL/P | Li et al.2022 | 35217232 | Confirmed |
| 53 | 12 | 111414461 | rs12229654 | | NSCLP | Yu et al.2017 | 28232668 | NA |
| 54 | 12 | 112518803 | rs11066150 | | NSCLP | Yu et al.2017 | 28232668 | NA |
| 54 | 12 | 112923393 | rs12229892 | | NSCLP | Yu et al.2017 | 28232668 | NA |
| 55 | 13 | 74316318 | rs12865628 | *KLF12* | NSOC | Yu et al.2023 | 37326468 | No significance |
| 56 | 13 | 80679302 | rs11841646 | *SPRY2* | NSCL/P,NSOC | Leslie et al.2017* | 28054174 | Confirmed |
| 56 | 13 | 80692811 | rs8001641 | *SPRY2* | NSCL/P | Ludwig et al.2012 | 22863734 | Confirmed |
| 56 | 13 | 80701485 | rs1854110 | *SPRY2* | NSCL/P | Li et al.2022 | 35217232 | Confirmed |
| 57 | 13 | 99395594 | rs4646211 | *DOCK9* | NSCPO | Huang et al.2019 | 31609978 | No significance |
| 58 | 13 | 100510372 | rs7982561 | *CLYBL* | NSCL/P | Li et al.2022 | 35217232 | Confirmed |
| 59 | 14 | 37239147 | rs730643 | *PAX9* | NSCPO | Huang et al.2019 | 31609978 | Confirmed |
| 59 | 14 | 37239854 | rs2415363 | *PAX9* | NSCPO | Huang et al.2019 | 31609978 | Confirmed |
| 60 | 14 | 51855553 | rs4901116 | *LINC00640* | NSCL/P | Li et al.2022 | 35217232 | Confirmed |
| 61 | 14 | 95375543 | rs1243567 | *GSC* | NSCL/P | Li et al.2022 | 35217232 | Confirmed |
| 62 | 15 | 33043657 | rs1919362 | *GREM1* | NSCL/P | Li et al.2022 | 35217232 | No significance |
| 63 | 15 | 63312632 | rs1873147 | | NSCL/P | Ludwig et al.2012 | 22863734 | No significance |

| | | | | | | | | |
|---|---|---|---|---|---|---|---|---|
| 64 | 15 | 74744399 | rs2289187 | *UBL7-DT* | NSCLP | Yu et al.2017 | 28232668 | Confirmed |
| 64 | 15 | 74853488 | rs8029215 | *UBL7-DT* | NSCL/P | Li et al.2022 | 35217232 | Confirmed |
| 64 | 15 | 74889163 | rs11072494 | *UBL7-DT* | NSCL/P | Leslie et al.2017* | 28054174 | Confirmed |
| 64 | 15 | 74899500 | rs6495117 | *UBL7-DT* | NSCLP | Yu et al.2017 | 28232668 | Confirmed |
| 65 | 16 | 3969297 | rs2283483 | *CREBBP* | NSCL/P | Li et al.2022 | 35217232 | Confirmed |
| 66 | 17 | 8930222 | rs11273201 | *NTN1* | NSCL/P | Leslie et al.2016 | 27033726 | Confirmed |
| 66 | 17 | 8930225 | rs7406226 | *NTN1* | NSCL/P | Leslie et al.2016 | 27033726 | Confirmed |
| 66 | 17 | 8932082 | rs4791331 | *NTN1* | NSCL/P | Li et al.2022 | 35217232 | Confirmed |
| 66 | 17 | 8932119 | rs4791774 | *NTN1* | NSCL/P | Sun et al.2015 | 25775280 | Confirmed |
| 66 | 17 | 8947708 | rs12944377 | *NTN1* | NSCL/P,NSOC | Leslie et al.2017* | 28054174 | Confirmed |
| 66 | 17 | 8948104 | rs16957821 | *NTN1* | NSCL/P | Mukhopadhyay et al.2( | 33898419 | Confirmed |
| 67 | 17 | 44977040 | rs12600562 | *WNT9B* | NSCL/P | Li et al.2022 | 35217232 | Confirmed |
| 68 | 17 | 54752926 | rs8182331 | *NOG* | NSCL/P | Li et al.2022 | 35217232 | Confirmed |
| 68 | 17 | 54773238 | rs227731 | *NOG* | NSCL/P | Ludwig et al.2012;Lesl | 22863734;280541 | Confirmed |
| 69 | 17 | 61076428 | rs1588366 | *TANC2* | NSCL/P | Leslie et al.2016;Mukh | 27033726;338984 | No significance |
| 70 | 17 | 73717421 | rs16967277 | *RECQL5* | NSCL/P | Li et al.2022 | 35217232 | No significance |
| 71 | 17 | 77267377 | rs1975866 | *RBFOX3* | NSCL/P | Mukhopadhyay et al.2( | 33898419 | No significance |
| 72 | 18 | 34116874 | rs4799867 | *FHOD3* | NSCL/P | Li et al.2022 | 35217232 | No significance |
| 73 | 18 | 37254234 | rs36019844 | *MIR924HG* | NSCLP | Avasthi et al.2021 | 34941638 | NA |
| 74 | 19 | 33521150 | rs73039428 | *RHPN2* | NSCL/P | Leslie et al.2016 | 27033726 | Confirmed |
| 74 | 19 | 33546283 | rs2003950 | *RHPN2* | NSCL/P | Li et al.2022 | 35217232 | Confirmed |
| 75 | 19 | 45882693 | rs8101662 | *KLC3* | NSCL/P | Li et al.2022 | 35217232 | No significance |
| 76 | 20 | 39261054 | rs6072081 | *MAFB* | NSCL/P,NSOC | Beaty et al.2010;Leslie | 20436469;280541 | Confirmed |
| 76 | 20 | 39261979 | rs6065259 | *MAFB* | NSCL/P | Beaty et al.2010 | 20436469 | Confirmed |
| 76 | 20 | 39268516 | rs17820943 | *MAFB* | NSCL/P,NSCLO | Beaty et al.2010;Huang | 20436469;316099 | Confirmed |
| 76 | 20 | 39269074 | rs13041247 | *MAFB* | NSCL/P | Beaty et al.2010;Ludw | 20436469;228637 | Confirmed |
| 76 | 20 | 39270816 | rs11696257 | *MAFB* | NSCL/P | Beaty et al.2010 | 20436469 | Confirmed |
| 76 | 20 | 39272959 | rs4812450 | *MAFB* | NSCL/P | Li et al.2022 | 35217232 | Confirmed |
| 76 | 20 | 39281629 | rs6102085 | *MAFB* | NSCL/P | Beaty et al.2010 | 20436469 | Confirmed |
| 77 | 20 | 50404452 | rs1018422 | *ATP9A* | NSCL/P | Li et al.2022 | 35217232 | No significance |
| 78 | 21 | 40014437 | rs463269 | *ERG* | NSCL/P | Li et al.2022 | 35217232 | Confirmed |
| 79 | 22 | 19799410 | rs2073764 | *ARVCF* | NSCLP | Yu et al.2017 | 28232668 | Confirmed |
| 79 | 22 | 19976845 | rs756653 | *ARVCF* | NSCL/P | Li et al.2022 | 35217232 | Confirmed |
| 80 | 22 | 31558820 | rs5753486 | *PLA2G3* | NSCL/P | Li et al.2022 | 35217232 | No significance |
| 81 | 22 | 45149303 | rs7284589 | *ARHGAP8* | NSCL/P | Li et al.2022 | 35217232 | Confirmed |

Note:NSOC, non-syndromic orofacial cleft cases by combining NSCLP,NSCLO and NSCPO together; NSCL/P, non-syndromic cleft lip with or without palate (NSCLP&NSCLO); NSCLP, non-syndromic cleft lip and palate; NSCLO, non-syndromic cleft lip only; NSCPO, non-syndromic cleft palate only; only listing the loci (lead SNPs) achiving genome -wide significant level (5.0E-08) in the discovery phase.

Supplementary Table 2. Sample information of multi-ancestry GWAS Meta-analysis

| Phenotype | Study | Population | Case | Control | Total |
|---|---|---|---|---|---|
| NSOC | POFC1 | Asian | 583 | 1589 | 2172 |
| | | European | 845 | 3343 | 4188 |
| | | Latin South American | 1298 | 3464 | 4762 |
| | CHW_Axiom | Han Chinese | 427 | 419 | 846 |
| | Huang et al | Han Chinese | 945 | 5068 | 6013 |
| | CHE_Axiom&HSA | Han Chinese | 322 | 722 | 1044 |
| | CHS_ZH | Han Chinese | 3012 | 11746 | 14758 |
| | Butail et al | African | 1019 | 2159 | 3178 |
| | GENEVA* | Mix Ancestry(Aisan&European) | | | 6203 (2042trios &18 multiplex pedigrees) |
| NSCL/P | POFC1 | Asian | 445 | 1246 | 1691 |
| | | European | 569 | 2704 | 3273 |
| | | Latin South American | 1050 | 2988 | 4038 |
| | CHW_Axiom | Han Chinese | 420 | 419 | 839 |
| | Huang et al | Han Chinese | 945 | 5068 | 6013 |
| | CHE_Axiom&HSA | Han Chinese | 322 | 722 | 1044 |
| | CHS_ZH | Han Chinese | 2025 | 11746 | 13771 |
| | Butail et al | African | 814 | 2159 | 2973 |
| | GENEVA* | Mix Ancestry(Aisan&European) | | | 4786 (1579trios &12 multiplex pedigrees) |
| NSCLP | POFC1 | Asian | 274 | 1055 | 1329 |
| | | European | 416 | 2454 | 2870 |
| | | Latin South American | 884 | 2766 | 3650 |
| | CHW_Axiom | Han Chinese | 275 | 419 | 694 |
| | CHE_Axiom&HSA | Han Chinese | 215 | 722 | 937 |
| | CHS_ZH | Han Chinese | 2025 | 11746 | 13771 |
| | GENEVA* | Mix Ancestry(Aisan&European) | | | 3411 (1126trios&8 multiplex pedigrees) |
| NSCLO | POFC1 | Asian | 171 | 785 | 956 |
| | | European | 153 | 1917 | 2070 |
| | | Latin South American | 166 | 1622 | 1788 |
| | CHW_Axiom | Han Chinese | 145 | 419 | 564 |
| | Huang et al. | Han Chinese | 945 | 5068 | 6013 |
| | CHE_Axiom&HSA | Han Chinese | 107 | 722 | 829 |
| | GENEVA* | Mix Ancestry(Aisan&European) | | | 1375 (453trios &4 multiplex pedigrees) |
| NSCPO | POFC1 | European | 38 | 835 | 873 |
| | | Latin South American | 21 | 626 | 647 |
| | Huang et al | Han Chinese | 930 | 5068 | 5998 |
| | CHS_ZH | Han Chinese | 987 | 11746 | 12733 |
| | Butail et al | African | 205 | 2159 | 2364 |
| | GENEVA* | Mix Ancestry(Aisan&European) | | | 1407 (460trios&6multiplex pedigrees) |

**Note**: NSOC, non-syndromic orofacial cleft cases by combining NSCLP,NSCLO and NSCPO together; NSCL/P, non-syndromic cleft lip with or without palate (NSCLP&NSCLO); NSCLP, non-syndromic cleft lip and palate; NSCLO, non-syndromic cleft lip only; NSCPO, non-syndromic cleft palate only. *, Individuals of the trios from Geneva study (Asian and European).

Supplementary Table 3. Cohort Studies Description, Genotypes

Descriptive genotyping statistics for studies that contributed to the GWAS meta-analysis, see column description under the table. Tests for deviation from Hardy Weinberg are 2-tailed.

| Study | Genotyping array(s) | Sample quality control | | SNP scaffold quality control | | | Prephasing | Imputation | | Association analysis | | Lambda | | | | |
|---|---|---|---|---|---|---|---|---|---|---|---|---|---|---|---|---|
| | | Call rate | Additional filters | Call rate | HWE P-value | MAF | software | Software | Reference panel | Software | Covariates or adjustment | NSOC | NSCL/P | NSCLP | NSCLO | NSCPO |
| CHW_Axiom | Affymetrix Axiom CHB | >98% | relatedness, sex discrepancy | >98% | >1e-6 | >1% | Eagle v2.4 | Minimac4 | 1000 genomes phase 3 | SNPTEST | PC1-10; Sex | 1.03093 | 1.030716 | 1.03536 | 0.98202 | -- |
| CHE_Axiom&HSA | Affymetrix Axiom CHB; Affymetrix Human SNP Ar | >98% | relatedness, nsex discrepancy | >98% | >1e-6 | >1% | Eagle v2.4 | Minimac4 | 1000 genomes phase 3 | SNPTEST | PC1-10; Sex | -- | 1.049698 | 1.040456 | NA | -- |
| CHS_ZH | Illumina HumanOmniZhongHua-8 BeadChip | >98% | relatedness, sex discrepancy | >98% | >1e-6 | >1% | Eagle v2.4 | Minimac4 | 1000 genomes phase 3 | regenie | PC1-10 | 1.109684 | -- | 1.091924 | -- | 1.0463 |
| CHW_ZH | Illumina HumanOmniZhongHua-8 BeadChip | >98% | relatedness, sex discrepancy | >98% | >1e-6 | >1% | Eagle v2.4 | Minimac4 | 1000 genomes phase 3 | SNPTEST | PC1-10; Sex | 1.015681 | 1.044482 | -- | 1.0485 | 1.00343 |
| Huang et al. 2019 | Illumina HumanOmniZhongHua-8 BeadChip | None | relatedness | >90% | >1e-6 | >1% | SHAPEIT2 | Minimac3 | 1000 Genomes phase 1 (ve | PLINK | PC1-4; Sex | -- | -- | -- | 1.031 | 1.016 |
| Butali et al.2019 | Illumina MEGA v2 15070954 A2 | None | batch effects, identification of large chromosomal anomalies, confirmation of relatedness (i.e. identity by descent) and establishment of continental ancestry with respect to HapMap samples | >98% | > 10−3 | >1% | IMPUTE2 | IMPUTE2 | 1000 genomes phase 3 | GMAAT package | population structure (the first seven eigenvectors of the genotypes), relationships between participants (using the computed genetic relatedness matrix) and covariates (sex and study site) | -- | -- | 0.996 | -- | 1 |
| POFC1 | Illumina HumanCore+Exome array | | | | > 0.0001 | >1% | SHAPEIT2 | IMPUTE2 | 1000 Genomes Phase 3 | PLINK | Genetic relationship matrix | 0.975035 | 0.987923 | 0.998601 | 1.08958 | 1.01313 |
| GENEVA | Illumina Human610-Quadv.1_B BeadChip | | | | > 0.0001 | >1% | SHAPEIT | IMPUTE2 | 1000 Genomes Phase 1 | PLINK | | 1.017371 | 1.020683 | 1.017371 | 1.00842 | 0.99534 |

**Note:**NSOC, non-syndromic orofacial cleft cases by combining NSCLP,NSCLO and NSCPO together; NSCL/P, non-syndromic cleft lip with or without palate (NSCLP&NSCLO); NSCLP, non-syndromic cleft lip and palate; NSCLO, non-syndromic cleft lip only; NSCPO, non-syndromic cleft palate only

| Column | Description |
|---|---|
| Study | The abbreviated name of the contributing study |
| Genotyping array(s) | The genotyping array used |
| Sample quality control | Call rate and other quality control filters |
| SNP scaffold quality control | SNP filters (HWE: Hardy-Weinberg equilibrium; MAF: minor allele frequency) |
| Prephasing software | Name and version of the software used for genotypes prephasing |
| Imputation | Software and reference panel used for genotypes imputation |
| Association analysis | Software and general covariates used for association analyses (PC: principal component) |
| Lambda | Inflation factor (lambda) for the adjusted GWASes |

SupTable 4. Summary of the lead SNPs in Known Loci by Cleft Group

| Phenotype | Locus Name | Chr | Position(b37) | rsid | EA | OE | Meta-analysis of CC&TDT | | | Meta-analsysi of CC | | | | Classification |
|---|---|---|---|---|---|---|---|---|---|---|---|---|---|---|
| | | | | | | | EAF | p-value | pHET | EAF | p-value | pHET-ANC | pHET-RES | |
| NSCLP | PAX7 | 1 | 18957297 | rs569017380 | G | GCCTCCT | 0.84 | 5.00E-08 | 1 | 0.16 | 5.00E-08 | 0.34 | 0.058 | Known Loci |
| **NSOC** | PAX7 | 1 | 18972776 | rs9439713 | A | G | 0.84 | 7.48E-17 | 0.62 | 0.15 | 1.30E-13 | 0.74 | 0.22 | Known SNP |
| NSCL/P | PAX7 | 1 | 18972776 | rs9439713 | A | G | 0.85 | 3.86E-18 | 0.082 | 0.14 | 2.01E-13 | 0.7 | 0.26 | Known SNP |
| NSCLP | PAX7 | 1 | 18972776 | rs9439713 | A | G | 0.87 | 4.09E-13 | 0.22 | 0.12 | 2.42E-10 | 0.91 | 0.015 | Known SNP |
| NSCPO | GRHL3 | 1 | 24667423 | rs12568609 | A | G | 0.86 | 1.69E-08 | 0.3 | 0.14 | 2.45E-07 | 0.009 | 0.71 | Known Loci |
| **NSOC** | FGGY | 1 | 59887483 | rs35110579 | G | A | 0.66 | 5.11E-09 | 1 | 0.34 | 5.11E-09 | 0.88 | 0.59 | Known Loci |
| NSCL/P | FGGY | 1 | 59887673 | rs11577647 | C | T | 0.66 | 5.28E-09 | 1 | 0.34 | 5.28E-09 | 0.9 | 0.68 | Known Loci |
| **NSOC** | ARHGAP29 | 1 | 94558110 | rs66515264 | T | G | 0.87 | 1.43E-13 | 0.005 | 0.13 | 7.58E-09 | 0.64 | 0.79 | Known SNP |
| NSCL/P | ARHGAP29 | 1 | 94558110 | rs66515264 | T | G | 0.87 | 5.08E-16 | 0.002 | 0.13 | 9.37E-11 | 0.38 | 0.74 | Known SNP |
| NSCLP | ARHGAP29 | 1 | 94558110 | rs66515264 | T | G | 0.85 | 1.63E-16 | 0.017 | 0.15 | 7.53E-12 | 0.77 | 0.88 | Known SNP |
| NSCL/P | ARHGAP29 | 1 | 94601200 | rs138088826 | C | CG | 0.65 | 1.36E-10 | 1 | 0.35 | 1.36E-10 | 0.001 | 0.69 | Known Loci |
| **NSOC** | ARHGAP29 | 1 | 94606077 | rs7551877 | A | G | 0.69 | 8.02E-09 | 1 | 0.31 | 8.02E-09 | 0.005 | 0.87 | Known Loci |
| NSCLP | ARHGAP29 | 1 | 94615394 | rs11290844 | A | AT | 0.61 | 1.95E-09 | 1 | 0.39 | 1.95E-09 | 0.009 | 0.2 | Known Loci |
| NSCLO | IRF6 | 1 | 209894822 | rs201481822 | AAG | A | 0.76 | 2.82E-13 | 1 | 0.24 | 2.82E-13 | 0.96 | 0.83 | Known Loci |
| **NSOC** | IRF6 | 1 | 209913473 | rs12120361 | G | T | 0.74 | 4.71E-18 | 0.8 | 0.26 | 2.41E-15 | 0.001 | 0.093 | Known Loci |
| NSCL/P | **IRF6** | **1** | **209913473** | **rs12120361** | **G** | **T** | 0.73 | **5.45E-22** | **0.14** | **0.26** | **1.91E-17** | **2.42E-08** | **0.26** | **Known Loci** |
| NSCLP | IRF6 | 1 | 209943893 | rs139818084 | G | GGTGT | 0.73 | 2.60E-18 | 1 | 0.27 | 2.60E-18 | 0.94 | 0.42 | Known Loci |
| NSCLO | IRF6 | 1 | 209974232 | rs622832 | A | C | 0.72 | 1.90E-22 | 0.11 | 0.28 | 2.60E-18 | 0.2 | 1 | Known Loci |
| NSCL/P | IRF6 | 1 | 209982738 | rs17015255 | G | A | 0.67 | 8.82E-28 | 0.000042 | 0.36 | 1.91E-17 | 0.001 | 0.14 | Known Loci |
| NSCPO | **IRF6** | **1** | **209989092** | **rs72741048** | **T** | **A** | 0.57 | **1.44E-15** | **0.3** | **0.44** | **1.19E-13** | **5.82E-11** | **0.37** | **Known SNP** |
| NSCLP | **IRF6** | **1** | **209989270** | **rs642961** | **A** | **G** | **0.76** | **5.42E-23** | **0.17** | **0.24** | **2.60E-18** | **0.000391** | **0.99** | **Known SNP** |
| **NSOC** | **IRF6** | **1** | **209992501** | **rs1109430** | **A** | **G** | 0.67 | **1.84E-25** | **0.025** | **0.36** | **1.21E-17** | **3.01E-08** | **0.012** | **Known SNP** |
| NSCLP | IRF6 | 1 | 210043322 | rs3028171 | C | A | 0.19 | 8.20E-10 | 0.43 | 0.19 | 1.78E-09 | 0.009 | 0.37 | Known Loci |
| NSCPO | **IRF6** | **1** | **210048819** | **rs2064163** | **T** | **G** | 0.57 | **1.13E-08** | **0.28** | **0.44** | **6.89E-09** | **1.60E-08** | **0.95** | **Known SNP** |
| NSCL/P | IRF6 | 1 | 210248548 | rs4844943 | T | C | 0.63 | 1.23E-24 | 0.013 | 0.4 | 1.91E-17 | 0.27 | 0.82 | Known Loci |
| **NSOC** | **IRF6** | **1** | **210253684** | **rs10863809** | **T** | **G** | 0.63 | **1.10E-20** | **0.37** | **0.4** | **5.36E-16** | **7.28E-05** | **0.16** | **Known Loci** |
| NSCLP | **IRF6** | **1** | **210301331** | **rs7516554** | **T** | **C** | **0.59** | **2.06E-21** | **0.68** | **0.4** | **2.60E-18** | **1.38E-07** | **0.44** | **Known Loci** |
| NSCLO | IRF6 | 1 | 210301331 | rs7516554 | T | C | 0.59 | 4.62E-14 | 0.28 | 0.4 | 1.01E-11 | 0.24 | 0.35 | Known Loci |
| NSCLP | IRF6 | 1 | 210352641 | rs2485896 | G | C | 0.2 | 1.11E-09 | 0.47 | 0.2 | 2.68E-09 | 0.003 | 0.71 | Known Loci |
| NSCPO | **IRF6** | **1** | **210355896** | **rs12087757** | **A** | **G** | 0.57 | **4.11E-12** | **0.36** | **0.44** | **1.20E-10** | **1.01E-09** | **0.57** | **Known Loci** |
| NSCL/P | IRF6 | 1 | 210372114 | rs2494175 | G | A | 0.61 | 2.90E-13 | 1 | 0.39 | 2.90E-13 | 0.075 | 0.31 | Known Loci |
| NSCPO | **IRF6** | **1** | **210372114** | **rs2494175** | **G** | **A** | 0.56 | **4.94E-10** | **1** | **0.44** | **4.94E-10** | **3.69E-09** | **0.73** | **Known Loci** |
| NSCL/P | TAF1B | 2 | 9967620 | rs528634626 | T | TA | 0.61 | 1.97E-08 | 1 | 0.39 | 1.97E-08 | 0.006 | 0.7 | Known Loci |
| NSCLP | TAF1B | 2 | 10028440 | rs71389302 | ATT | A | 0.34 | 1.32E-08 | 1 | 0.34 | 1.32E-08 | 0.11 | 0.44 | Known Loci |
| NSCLP | FAM49A | 2 | 16705500 | rs140385594 | TGTGA | T | 0.27 | 2.60E-18 | 1 | 0.27 | 2.60E-18 | 0.013 | 0.55 | Known Loci |
| NSCL/P | FAM49A | 2 | 16713395 | rs6745357 | G | C | 0.37 | 1.21E-17 | 1 | 0.63 | 1.21E-17 | 0.002 | 0.043 | Known SNP |
| NSCLO | FAM49A | 2 | 16713395 | rs6745357 | G | C | 0.43 | 1.95E-11 | 1 | 0.57 | 1.95E-11 | 0.31 | 0.52 | Known SNP |
| **NSOC** | FAM49A | 2 | 16733525 | rs5007483 | T | G | 0.47 | 9.97E-18 | 0.15 | 0.54 | 2.26E-17 | 0.12 | 0.011 | Known Loci |
| NSCL/P | HYAL2 | 3 | 50318486 | rs11434969 | GA | G | 0.5 | 8.70E-10 | 1 | 0.5 | 8.70E-10 | 0.14 | 0.57 | Known Loci |
| **NSOC** | ADAMTS9 | 3 | 64613278 | rs6445418 | C | G | 0.49 | 1.37E-08 | 0.7 | 0.51 | 3.26E-07 | 0.27 | 0.3 | Known Loci |
| NSCL/P | ADAMTS9 | 3 | 64615873 | rs9838262 | A | G | 0.53 | 6.69E-09 | 0.44 | 0.46 | 2.55E-07 | 0.24 | 0.78 | Known Loci |
| **NSOC** | FILIP1L | 3 | 99680753 | rs11290599 | AG | A | 0.76 | 1.54E-10 | 1 | 0.24 | 1.54E-10 | 0.22 | 0.68 | Known Loci |
| NSCL/P | FILIP1L | 3 | 99680753 | rs11290599 | AG | A | 0.76 | 1.82E-11 | 1 | 0.25 | 1.82E-11 | 0.12 | 0.22 | Known Loci |
| NSCLP | FILIP1L | 3 | 99680753 | rs11290599 | AG | A | 0.27 | 1.97E-09 | 1 | 0.27 | 1.97E-09 | 0.026 | 0.21 | Known Loci |
| NSCL/P | TP63 | 3 | 189544776 | rs75834417 | C | A | 0.96 | 5.08E-09 | 0.003 | 0.04 | 3.09E-05 | 0.63 | 0.36 | Known Loci |
| NSCLP | TP63 | 3 | 189549082 | rs79792381 | T | C | 0.97 | 6.15E-09 | 0.4 | 0.03 | 2.93E-07 | 0.8 | 0.91 | Known Loci |
| **NSOC** | TP63 | 3 | 189551340 | rs55660938 | T | C | 0.96 | 3.08E-08 | 0.05 | 0.04 | 3.05E-05 | 0.18 | 0.72 | Known Loci |
| NSCLP | NNT | 5 | 43856675 | rs34871596 | A | G | 0.85 | 6.96E-09 | 0.69 | 0.16 | 2.10E-07 | 0.62 | 0.65 | Known Loci |
| NSCL/P | NNT | 5 | 43862510 | rs34378343 | C | T | 0.84 | 1.97E-09 | 0.45 | 0.16 | 1.70E-07 | 0.6 | 0.96 | Known Loci |
| NSCLP | GMDS | 6 | 1944939 | rs11242726 | A | G | 0.53 | 3.66E-08 | 0.15 | 0.47 | 3.79E-06 | 0.18 | 0.92 | Known Loci |
| **NSOC** | GMDS | 6 | 1960651 | rs140236874 | TA | T | 0.86 | 1.18E-11 | 1 | 0.14 | 1.17E-11 | 0.58 | 0.75 | Known Loci |
| NSCL/P | GMDS | 6 | 1960651 | rs140236874 | TA | T | 0.86 | 1.41E-12 | 1 | 0.14 | 1.41E-12 | 0.26 | 0.91 | Known Loci |
| NSCLP | ZFHX4 | 8 | 77496113 | rs11993273 | G | A | 0.5 | 2.22E-08 | 1 | 0.5 | 2.22E-08 | 0.078 | 0.029 | Known SNP |
| **NSOC** | ZFHX4 | 8 | 77498222 | rs144305105 | ATTATT | A | 0.5 | 5.59E-09 | 1 | 0.5 | 5.59E-09 | 0.014 | 0.099 | Known Loci |
| NSCL/P | ZFHX4 | 8 | 77498222 | rs144305105 | ATTATT | A | 0.5 | 1.67E-11 | 1 | 0.5 | 1.66E-11 | 0.01 | 0.13 | Known Loci |
| **NSOC** | DCAF4L2 | 8 | 88946414 | rs5893033 | TA | T | 0.61 | 1.80E-11 | 1 | 0.39 | 1.80E-11 | 0.013 | 0.28 | Known Loci |
| NSCL/P | DCAF4L2 | 8 | 88946414 | rs5893033 | TA | T | 0.61 | 6.30E-15 | 1 | 0.39 | 6.30E-15 | 0.029 | 0.058 | Known Loci |
| NSCLP | DCAF4L2 | 8 | 88946414 | rs5893033 | TA | T | 0.38 | 1.68E-11 | 1 | 0.38 | 1.68E-11 | 0.65 | 0.3 | Known Loci |
| **NSOC** | RIPK2 | 8 | 90232609 | rs141180188 | CT | C | 0.57 | 1.76E-08 | 1 | 0.43 | 1.76E-08 | 0.69 | 0.7 | Known Loci |
| **NSOC** | VIRMA | 8 | 95503542 | rs58089312 | CA | C | 0.61 | 9.52E-13 | 1 | 0.39 | 9.52E-13 | 0.17 | 0.62 | Known Loci |
| NSCL/P | VIRMA | 8 | 95503542 | rs58089312 | CA | C | 0.6 | 2.02E-12 | 1 | 0.4 | 2.01E-12 | 0.23 | 0.15 | Known Loci |
| NSCLP | VIRMA | 8 | 95663822 | rs10089502 | C | T | 0.17 | 1.86E-11 | 0.89 | 0.17 | 2.62E-10 | 0.007 | 0.17 | Known Loci |
| NSCL/P | VIRMA | 8 | 95739642 | rs67763258 | T | G | 0.78 | 2.31E-08 | 0.107 | 0.22 | 3.18E-06 | 0.036 | 0.068 | Known Loci |
| **NSOC** | LINC00824 | 8 | 129699660 | rs7839784 | T | C | 0.85 | 1.53E-19 | 0.67 | 0.15 | 2.31E-16 | 0.001 | 0.44 | Known Loci |
| NSCL/P | LINC00824 | **8** | **129699660** | **rs7839784** | **T** | **C** | 0.85 | **5.78E-20** | **0.34** | **0.15** | **1.99E-16** | **5.97E-05** | **0.29** | **Known Loci** |
| NSCLO | LINC00824 | **8** | **129699660** | **rs7839784** | **T** | **C** | 0.79 | **5.15E-09** | **0.55** | **0.22** | **8.89E-08** | **0.00038** | **0.77** | **Known Loci** |
| NSCLP | LINC00824 | 8 | 129703990 | rs35379093 | T | C | 0.73 | 3.90E-14 | 0.39 | 0.26 | 1.40E-11 | 0.0021 | 0.042 | Known Loci |
| **NSOC** | LINC00824 | **8** | **129911532** | **rs10631848** | **GGTGTGT** | **G** | 0.8 | **1.91E-17** | **1** | **0.2** | **1.91E-17** | **3.58E-05** | **0.14** | **Known Loci** |
| NSCL/P | LINC00824 | **8** | **129911532** | **rs10631848** | **GGTGTGT** | **G** | 0.81 | **1.91E-17** | **1** | **0.2** | **1.91E-17** | **5.98E-05** | **0.033** | **Known Loci** |
| NSCLP | LINC00824 | 8 | 129911532 | rs10631848 | GGT | G | 0.63 | 1.02E-09 | 1 | 0.63 | 1.02E-09 | 0.76 | 0.057 | Known Loci |
| NSCLO | LINC00824 | 8 | 129950399 | rs1372450 | T | C | 0.84 | 2.39E-16 | 0.049 | 0.15 | 8.67E-13 | 0.25 | 0.54 | Known Loci |
| **NSOC** | LINC00824 | 8 | 129964873 | rs17242358 | A | G | 0.92 | 1.06E-27 | 0.001 | 0.08 | 1.21E-17 | 0.15 | 0.2 | Known SNP |
| NSCL/P | LINC00824 | 8 | 129976136 | rs55658222 | A | G | 0.92 | 9.30E-29 | 3.93E-06 | 0.08 | 1.91E-17 | 0.26 | 0.22 | Known SNP |
| NSCLP | LINC00824 | 8 | 129976136 | rs55658222 | A | G | 0.93 | 3.34E-26 | 0.001 | 0.07 | 2.60E-18 | 0.36 | 0.26 | Known SNP |
| NSCL/P | GADD45G | 9 | 92210595 | rs7024024 | T | A | 0.74 | 7.76E-11 | 0.55 | 0.26 | 4.19E-09 | 0.65 | 0.028 | Known Loci |
| NSCLP | GADD45G | 9 | 92210595 | rs7024024 | T | A | 0.26 | 4.09E-08 | 0.56 | 0.26 | 8.98E-07 | 0.96 | 0.075 | Known Loci |
| **NSOC** | GADD45G | 9 | 92211862 | rs10429497 | A | G | 0.73 | 1.23E-09 | 0.38 | 0.27 | 1.24E-07 | 0.029 | 0.27 | Known Loci |
| NSCLP | PTCH1 | 9 | 98225056 | rs28377268 | T | G | 0.82 | 3.91E-08 | 1 | 0.18 | 3.91E-08 | 0.12 | 0.32 | Known Loci |
| **NSOC** | FOXE1 | 9 | 100508754 | rs541315146 | CT | C | 0.86 | 9.91E-11 | 1 | 0.14 | 9.91E-11 | 0.049 | 0.049 | Known Loci |
| **NSOC** | FOXE1 | 9 | 100619090 | rs35060625 | TCTC | T | 0.75 | 4.83E-08 | 1 | 0.25 | 4.83E-08 | 0.3 | 0.1 | Known Loci |
| **NSOC** | NR6A1 | 9 | 127369279 | rs77346093 | C | T | 0.88 | 9.72E-13 | 0.56 | 0.13 | 2.82E-10 | 0.46 | 0.4 | Known Loci |
| NSCL/P | NR6A1 | 9 | 127369279 | rs77346093 | C | T | 0.88 | 6.81E-13 | 0.42 | 0.13 | 1.78E-10 | 0.45 | 0.68 | Known Loci |
| NSCLP | VAX1 | 10 | 118831210 | rs10886037 | T | A | 0.37 | 1.57E-22 | 0.27 | 0.38 | 2.60E-18 | 0.001 | 0.19 | Known Loci |
| NSCLO | VAX1 | 10 | 118836682 | rs11197887 | A | T | 0.72 | 1.30E-10 | 0.93 | 0.28 | 9.22E-10 | 0.97 | 0.004 | Known Loci |
| NSCL/P | VAX1 | 10 | 118846294 | rs10886040 | G | C | 0.64 | 5.16E-22 | 0.29 | 0.36 | 1.91E-17 | 0.004 | 0.02 | Known SNP |
| **NSOC** | VAX1 | 10 | 118850264 | rs17095834 | G | A | 0.65 | 5.66E-22 | 0.47 | 0.35 | 1.91E-17 | 0.093 | 0.25 | Known Loci |
| **NSOC** | LGR5 | 12 | 71967309 | rs67687642 | A | AT | 0.67 | 7.07E-12 | 1 | 0.33 | 7.07E-12 | 0.66 | 0.13 | Known Loci |
| NSCL/P | LGR5 | 12 | 71967309 | rs67687642 | A | AT | 0.66 | 1.27E-16 | 1 | 0.34 | 1.27E-16 | 0.084 | 0.006 | Known Loci |
| NSCLP | LGR5 | 12 | 71967309 | rs67687642 | A | AT | 0.66 | 5.37E-13 | 1 | 0.34 | 5.37E-13 | 0.13 | 0.035 | Known Loci |
| **NSOC** | SPRY2 | 13 | 80679302 | rs11841646 | A | T | 0.75 | 8.55E-12 | 0.47 | 0.24 | 1.50E-09 | 0.15 | 0.55 | Known SNP |
| NSCL/P | SPRY2 | 13 | 80679302 | rs11841646 | A | T | 0.76 | 6.38E-14 | 0.054 | 0.23 | 2.33E-10 | 0.104 | 0.32 | Known Loci |
| NSCLP | SPRY2 | 13 | 80679302 | rs11841646 | A | T | 0.75 | 2.60E-14 | 0.27 | 0.24 | 1.80E-11 | 0.16 | 0.91 | Known SNP |
| **NSOC** | CLYBL | 13 | 100512107 | rs9557318 | A | G | 0.68 | 1.88E-09 | 0.84 | 0.33 | 4.18E-08 | 0.23 | 0.63 | Known Loci |
| NSCL/P | CLYBL | 13 | 100512107 | rs9557318 | A | G | 0.67 | 1.79E-12 | 0.76 | 0.34 | 8.72E-11 | 0.28 | 0.048 | Known Loci |
| NSCLP | CLYBL | 13 | 100512107 | rs9557318 | A | G | 0.65 | 1.11E-11 | 0.92 | 0.36 | 1.79E-10 | 0.024 | 0.49 | Known Loci |
| NSCPO | **PAX9** | **14** | **37227795** | **rs555814798** | **CA** | **C** | 0.34 | **5.86E-09** | **1** | **0.66** | **5.86E-09** | **4.88E-05** | **0.81** | **Known Loci** |
| NSCL/P | PAX9 | 14 | 37458914 | rs10483482 | T | C | 0.8 | 7.13E-11 | 0.67 | 0.2 | 5.13E-10 | 0.6 | 0.034 | Known Loci |
| NSCLP | PAX9 | 14 | 37464217 | rs75088305 | A | G | 0.8 | 5.49E-10 | 0.77 | 0.21 | 1.79E-08 | 0.64 | 0.058 | Known Loci |

| | | | | | | | | | | | | | | |
|---|---|---|---|---|---|---|---|---|---|---|---|---|---|---|
| NSCLP | ***PAX9*** | **14** | **37514564** | **rs10150336** | **A** | **T** | **0.5** | **4.74E-08** | **1** | **0.5** | **4.74E-08** | **0.00035** | **0.025** | **Known Loci** |
| NSCL/P | *LINC00640* | 14 | 51855553 | rs4901116 | T | G | 0.63 | 6.54E-14 | 0.78 | 0.37 | 4.63E-12 | 0.13 | 0.72 | Known SNP |
| NSCLP | *LINC00640* | 14 | 51856064 | rs144433632 | AAT | A | 0.44 | 1.20E-10 | 1 | 0.44 | 1.20E-10 | 0.31 | 0.99 | Known Loci |
| NSCL/P | *GSC* | 14 | 95378540 | rs61665130 | T | TG | 0.52 | 4.78E-08 | 1 | 0.48 | 4.78E-08 | 0.047 | 0.084 | Known Loci |
| NSCLP | *GSC* | 14 | 95378540 | rs61665130 | T | TG | 0.5 | 5.33E-09 | 1 | 0.5 | 5.33E-09 | 0.015 | 0.56 | Known Loci |
| **NSOC** | *UBL7-AS1* | 15 | 74759689 | rs559701024 | CT | C | 0.6 | 1.72E-08 | 1 | 0.4 | 1.72E-08 | 0.29 | 0.87 | Known Loci |
| NSCL/P | *UBL7-AS1* | 15 | 74759689 | rs559701024 | CT | C | 0.59 | 6.78E-14 | 1 | 0.41 | 6.78E-14 | 0.033 | 0.36 | Known Loci |
| NSCLP | *UBL7-AS1* | 15 | 74759689 | rs559701024 | CT | C | 0.59 | 2.07E-12 | 1 | 0.41 | 2.07E-12 | 0.062 | 0.17 | Known Loci |
| **NSOC** | *CREBBP* | 16 | 3963187 | rs9923088 | C | G | 0.43 | 5.45E-10 | 1 | 0.57 | 5.45E-10 | 0.054 | 0.51 | Known Loci |
| NSCL/P | *CREBBP* | 16 | 3963187 | rs9923088 | C | G | 0.42 | 2.25E-15 | 1 | 0.58 | 2.25E-15 | 0.061 | 0.8 | Known Loci |
| NSCLP | *CREBBP* | 16 | 3963187 | rs9923088 | C | G | 0.44 | 5.98E-10 | 1 | 0.56 | 5.98E-10 | 0.1 | 0.53 | Known Loci |
| NSCLP | *NTN1* | 17 | 8930279 | rs35023427 | T | TA | 0.72 | 1.66E-08 | 1 | 0.28 | 1.66E-08 | 0.053 | 0.49 | Known Loci |
| NSCLO | *NTN1* | 17 | 8943063 | rs9911652 | T | C | 0.82 | 3.14E-10 | 0.064 | 0.18 | 7.64E-08 | 0.16 | 0.4 | Known Loci |
| **NSOC** | *NTN1* | 17 | 8947708 | rs12944377 | C | T | 0.43 | 3.55E-24 | 0.063 | 0.57 | 1.91E-17 | 0.006 | 0.54 | Known SNP |
| NSCL/P | ***NTN1*** | **17** | **8947708** | **rs12944377** | **C** | **T** | 0.43 | **2.66E-26** | **0.001** | **0.57** | **1.21E-17** | **0.00025** | **0.086** | **Known SNP** |
| NSCLP | *NTN1* | 17 | 8947708 | rs12944377 | C | T | 0.63 | 1.58E-25 | 0.004 | 0.63 | 2.60E-18 | 0.45 | 0.16 | Known SNP |
| NSCL/P | WNT9B | 17 | 44993128 | rs4968248 | G | A | 0.53 | 3.19E-14 | 0.84 | 0.47 | 2.03E-12 | 0.63 | 0.52 | Known Loci |
| NSCLP | WNT9B | 17 | 45005703 | rs3785888 | C | T | 0.48 | 5.42E-12 | 0.62 | 0.48 | 3.68E-11 | 0.39 | 0.8 | Known Loci |
| **NSOC** | WNT9B | 17 | 45042079 | rs3760377 | A | G | 0.5 | 2.35E-11 | 0.47 | 0.5 | 3.38E-09 | 0.84 | 0.52 | Known Loci |
| NSCL/P | *NOG* | 17 | 54773238 | rs227731 | G | T | 0.62 | 2.92E-14 | 0.14 | 0.38 | 4.34E-11 | 0.018 | 0.46 | Known SNP |
| NSCLP | *NOG* | 17 | 54773238 | rs227731 | G | T | 0.36 | 2.23E-08 | 0.59 | 0.35 | 4.88E-07 | 0.62 | 0.33 | Known SNP |
| **NSOC** | *NOG* | 17 | 54776955 | rs227727 | T | A | 0.62 | 1.57E-10 | 0.9 | 0.38 | 2.39E-09 | 0.017 | 0.12 | Known Loci |
| **NSOC** | *RHPN2* | 19 | 33515581 | rs145403118 | C | CT | 0.85 | 4.85E-10 | 1 | 0.15 | 4.85E-10 | 0.038 | 0.31 | Known Loci |
| NSCL/P | *RHPN2* | 19 | 33515581 | rs145403118 | C | CT | 0.85 | 2.45E-16 | 1 | 0.15 | 2.45E-16 | 0.013 | 0.96 | Known Loci |
| NSCLP | *RHPN2* | 19 | 33517152 | rs28570619 | A | G | 0.88 | 8.92E-14 | 1 | 0.12 | 8.91E-14 | 1 | 0.7 | Known Loci |
| **NSOC** | *RHPN2* | 19 | 33525145 | rs10409307 | A | G | 0.53 | 3.61E-08 | 1 | 0.47 | 3.61E-08 | 0.92 | 0.42 | Known Loci |
| NSCL/P | *RHPN2* | 19 | 33525145 | rs10409307 | A | G | 0.53 | 1.53E-09 | 1 | 0.47 | 1.53E-09 | 0.68 | 0.34 | Known Loci |
| NSCLP | *RHPN2* | 19 | 33545042 | rs12462348 | T | C | 0.68 | 1.59E-08 | 0.99 | 0.32 | 1.36E-07 | 0.095 | 0.62 | Known Loci |
| NSCLO | *MAFB* | 20 | 39265737 | rs6029242 | T | C | 0.48 | 1.25E-09 | 0.54 | 0.52 | 2.63E-08 | 0.007 | 0.33 | Known Loci |
| NSCL/P | *MAFB* | 20 | 39270795 | rs6124259 | G | A | 0.6 | 3.73E-17 | 0.011 | 0.39 | 2.84E-12 | 0.031 | 0.64 | Known Loci |
| **NSOC** | *MAFB* | 20 | 39272739 | rs4812449 | G | C | 0.55 | 2.46E-15 | 1 | 0.45 | 2.45E-15 | 0.006 | 0.05 | Known Loci |
| NSCL/P | ***MAFB*** | **20** | **39274534** | **rs71193616** | **GGGA** | **G** | 0.63 | **1.25E-12** | **1** | **0.37** | **1.25E-12** | **0.00013** | **0.28** | **Known Loci** |
| NSCLP | *MAFB* | 20 | 39281629 | rs6102085 | G | A | 0.48 | 2.35E-14 | 0.006 | 0.48 | 7.67E-10 | 0.008 | 0.28 | Known SNP |
| **NSOC** | *ERG* | 21 | 40031924 | rs9647180 | T | C | 0.76 | 3.58E-09 | 1 | 0.24 | 3.58E-09 | 0.75 | 0.37 | Known Loci |
| NSCL/P | *ERG* | 21 | 40034216 | rs59685837 | GA | G | 0.76 | 6.86E-09 | 1 | 0.24 | 6.86E-09 | 0.75 | 0.96 | Known Loci |
| NSCLP | *ARVCF* | 22 | 19972118 | rs9606203 | A | C | 0.81 | 1.42E-08 | 0.57 | 0.19 | 3.48E-07 | 0.004 | 0.18 | Known Loci |
| NSCL/P | *ARVCF* | 22 | 19976845 | rs756653 | A | G | 0.81 | 3.00E-11 | 0.44 | 0.19 | 2.78E-09 | 0.008 | 0.54 | Known SNP |
| NSCL/P | *ARHGAP8* | 22 | 45150659 | rs5765956 | T | C | 0.52 | 2.24E-08 | 0.006 | 0.48 | 3.97E-05 | 0.21 | 0.31 | Known Loci |

Note: The known loci is defined as its location within 1Mb from previous GWASs reported SNP.NSOC, non-syndromic orofacial cleft cases by combining NSCLP,NSCLO and NSCPO together; NSCL/P, non-syndromic cleft lip with or without palate (NSCLP&NSCLO); NSCLP, non-syndromic cleft lip and palate; NSCLO, non-syndromic cleft lip only; NSCPO, non-syndromic cleft palate only;EA, effect allele; OA, othr allele; CC, Case-control design Study; TDT, Transmission disequilibrium test; pHET; P-value for heterogeneity;pHET-ANC: P-value for heterogeneity correlated with ancestry. pHET-RES: P-value for residual heterogeneity.

Supplementary Table 5. Funcational Annotation on the lead SNPs by VEP from Ensemble

| Classification | Locus Name | Chromosome | Position(b37) | rsid | Function | Feature_type | BIOTYPE | Consequence |
|---|---|---|---|---|---|---|---|---|
| Novel | *NTRK1* | 1 | *156839414* | rs4661229 | *intronic* | Transcript;RegulatoryFeature | protein_coding;protein_coding_CDS_not_defined;nonsense_mediated_decay;retained_intron;CTCF_binding_site;enhancer | downstream_gene_variant;intron_variant;intron_variant,non_coding_transcript_variant;intron_variant,NMD_transcript_variant;regulatory_region_variant |
| Novel | *HMGCR* | 2 | *219817037* | rs13011262 | | - | - | intergenic_variant |
| Novel | *CALD1* | 7 | *134845630* | rs4732060 | *intronic* | Transcript;RegulatoryFeature;MotifFeature | protein_coding;nonsense_mediated_decay;retained_intron;protein_coding_CDS_not_defined;lncRNA;enhancer;CTCF_binding_site | intron_variant;intron_variant,NMD_transcript_variant;intron_variant,non_coding_transcript_variant;upstream_gene_variant;regulatory_region_variant;TF_binding_site_variant |
| Novel | *CALD1* | 7 | *134877068* | rs10488465 | *intronic* | Transcript | protein_coding;nonsense_mediated_decay;retained_intron;protein_coding_CDS_not_defined;lncRNA | intron_variant;intron_variant,NMD_transcript_variant;intron_variant,non_coding_transcript_variant |
| Novel | *SHH* | 7 | *156026696* | rs4716972 | | Transcript | lncRNA | downstream_gene_variant;intron_variant,non_coding_transcript_variant |
| Novel | *NRG1* | 8 | *32493817* | rs17645417 | *intronic* | Transcript | protein_coding;protein_coding_CDS_not_defined;nonsense_mediated_decay | intron_variant;intron_variant,non_coding_transcript_variant;intron_variant,NMD_transcript_variant |
| Novel | *PRICKLE1* | 12 | *42488641* | rs12817499 | *intronic* | Transcript | protein_coding;lncRNA;protein_coding_CDS_not_defined | intron_variant;upstream_gene_variant;intron_variant,non_coding_transcript_variant |
| Novel | *ALX1* | 12 | *85585770* | rs565838209 | | Transcript | lncRNA | intron_variant,non_coding_transcript_variant |
| Novel | *SOX9* | 17 | *72283506* | rs62069766 | | RegulatoryFeature | open_chromatin_region | regulatory_region_variant;intergenic_variant |
| Novel | *LINC00320* | 21 | *20793565* | rs13052576 | *intronic* | Transcript;RegulatoryFeature | lncRNA;enhancer | intron_variant,non_coding_transcript_variant;regulatory_region_variant |
| Novel | *RUNX1* | 21 | *34831549* | rs2246738 | *intronic* | Transcript | protein_coding;retained_intron;protein_coding_CDS_not_defined;nonsense_mediated_decay;lncRNA | intron_variant;downstream_gene_variant;upstream_gene_variant;intron_variant,NMD_transcript_variant |
| Novel | *MYH9* | 22 | 36288285 | rs5756130 | synonymous | Transcript;RegulatoryFeature | protein_coding;protein_coding_CDS_not_defined;retained_intron;miRNA;enhancer | synonymous_variant;upstream_gene_variant;non_coding_transcript_exon_variant;regulatory_region_variant;intergenic_variant |
| Known SNP | *PAX7* | 1 | 18972776 | rs9439713 | | Transcript | protein_coding | intron_variant |
| Known SNP | *ARHGAP29* | 1 | 94558110 | rs66515264 | Intronic | Transcript | protein_coding | intron_variant |
| Known SNP | *IRF6* | 1 | 209989092 | rs72741048 | | RegulatoryFeature | enhancer | regulatory_region_variant;intergenic_variant |
| Known SNP | *IRF6* | 1 | 209989270 | rs642961 | | RegulatoryFeature | enhancer | regulatory_region_variant;intergenic_variant |
| Known SNP | *IRF6* | 1 | 209992501 | rs1109430 | | - | - | intergenic_variant |
| Known SNP | *IRF6* | 1 | 210048819 | rs2064163 | | RegulatoryFeature | enhancer;CTCF_binding_site | regulatory_region_variant;intergenic_variant |
| Known SNP | *FAM49A* | 2 | 16713395 | rs6745357 | Intronic | Transcript | lncRNA | downstream_gene_variant;intron_variant,non_coding_transcript_variant |
| Known SNP | ZFHX4 | 8 | 77496113 | rs11993273 | Intronic | Transcript | lncRNA | intron_variant,non_coding_transcript_variant;upstream_gene_variant |
| Known SNP | *LINC00824* | 8 | 129964873 | rs17242358 | | Transcript | lncRNA | downstream_gene_variant;intron_variant,non_coding_transcript_variant |
| Known SNP | *LINC00824* | 8 | 129976136 | rs55658222 | Intronic | Transcript | lncRNA | intron_variant,non_coding_transcript_variant |
| Known SNP | *VAX1* | 10 | 118846294 | rs10886040 | Intronic | RegulatoryFeature;Transcript | enhancer;protein_coding | regulatory_region_variant;intron_variant |
| Known SNP | *SPRY2* | 13 | 80679302 | rs11841646 | | - | - | intergenic_variant |
| Known SNP | *LINC00640* | 14 | 51855553 | rs4901116 | Intronic | Transcript | lncRNA | downstream_gene_variant;intron_variant,non_coding_transcript_variant;upstream_gene_variant |
| Known SNP | *NTN1* | 17 | 8947708 | rs12944377 | Intronic | Transcript | protein_coding | intron_variant |
| Known SNP | *NOG* | 17 | 54773238 | rs227731 | | - | - | intergenic_variant |
| Known SNP | *MAFB* | 20 | 39281629 | rs6102085 | | - | - | intergenic_variant |
| Known SNP | *ARVCF* | 22 | 19976845 | rs756653 | Intronic | Transcript | protein_coding;protein_coding_CDS_not_defined;retained_intron | intron_variant;upstream_gene_variant;intron_variant,non_coding_transcript_variant |
| Known Loci | *PAX7* | 1 | 18957297 | rs569017380 | | Transcript;RegulatoryFeature | protein_coding;promoter | upstream_gene_variant;regulatory_region_variant |
| Known Loci | *GRHL3* | 1 | 24667423 | rs12568609 | Intronic | Transcript;RegulatoryFeature | protein_coding;retained_intron;protein_coding_CDS_not_defined;enhancer | intron_variant;intron_variant,non_coding_transcript_variant;downstream_gene_variant;regulatory_region_variant |

| | | | | | | | | |
|---|---|---|---|---|---|---|---|---|
| Known Loci | *FGGY* | 1 | *59421811* | rs35110579 | *intronic* | Transcript;RegulatoryFeature | protein_coding;nonsense_mediated_decay;protein_coding_CDS_not_defined;retained_intron;misc_RNA;enhancer | intron_variant;intron_variant,NMD_transcript_variant;intron_variant,non_coding_transcript_variant;5_prime_UTR_variant;regulatory_region_variant |
| Known Loci | *FGGY* | 1 | *59422001* | rs11577647 | *intronic* | Transcript | protein_coding;nonsense_mediated_decay;protein_coding_CDS_not_defined;retained_intron;misc_RNA | intron_variant;intron_variant,NMD_transcript_variant;intron_variant,non_coding_transcript_variant;5_prime_UTR_variant |
| Known Loci | *ARHGAP29* | 1 | 94601200 | rs138088826 | | - | - | intergenic_variant |
| Known Loci | *ARHGAP29* | 1 | 94606077 | rs7551877 | | Transcript | lncRNA | upstream_gene_variant |
| Known Loci | *ARHGAP29* | 1 | 94615394 | rs11290844 | Exonic | Transcript | lncRNA;retained_intron;nonsense_mediated_decay;misc_RNA | downstream_gene_variant;non_coding_transcript_exon_variant;downstream_gene_variant;upstream_gene_variant |
| Known Loci | *IRF6* | 1 | 209894822 | rs201481822 | Intronic | Transcript;RegulatoryFeature | protein_coding;lncRNA;enhancer | intron_variant;intron_variant,non_coding_transcript_variant;regulatory_region_variant |
| Known Loci | *IRF6* | 1 | 209913473 | rs12120361 | | Transcript | processed_pseudogene | upstream_gene_variant |
| Known Loci | *IRF6* | 1 | 209943893 | rs139818084 | Intronic | Transcript;RegulatoryFeature | protein_coding;retained_intron;nonsense_mediated_decay;CTCF_binding_site;promoter | intron_variant;intron_variant,non_coding_transcript_variant;intron_variant,NMD_transcript_variant;regulatory_region_variant |
| Known Loci | *IRF6* | 1 | 209974232 | rs622832 | Intronic | Transcript | protein_coding;nonsense_mediated_decay | intron_variant;intron_variant,NMD_transcript_variant |
| Known Loci | *IRF6* | 1 | 209982738 | rs17015255 | | Transcript | protein_coding;nonsense_mediated_decay | upstream_gene_variant |
| Known Loci | *IRF6* | 1 | 210043322 | rs3028171 | | RegulatoryFeature | enhancer | regulatory_region_variant;intergenic_variant |
| Known Loci | *IRF6* | 1 | 210248548 | rs4844943 | Intronic | Transcript | protein_coding;nonsense_mediated_decay | intron_variant;intron_variant,NMD_transcript_variant |
| Known Loci | *IRF6* | 1 | 210253684 | rs10863809 | Intronic | Transcript | protein_coding;nonsense_mediated_decay | intron_variant;intron_variant,NMD_transcript_variant |
| Known Loci | *IRF6* | 1 | 210301331 | rs7516554 | Intronic | Transcript | protein_coding;nonsense_mediated_decay | intron_variant;intron_variant,NMD_transcript_variant |
| Known Loci | *IRF6* | 1 | 210352641 | rs2485896 | | - | - | intergenic_variant |
| Known Loci | *IRF6* | 1 | 210355896 | rs12087757 | | - | - | intergenic_variant |
| Known Loci | *IRF6* | 1 | 210372114 | rs2494175 | | - | - | intergenic_variant |
| Known Loci | *TAF1B* | 2 | 9967620 | rs528634626 | | - | - | intergenic_variant |
| Known Loci | *TAF1B* | 2 | 10028440 | rs71389302 | Intronic | Transcript | nonsense_mediated_decay; protein_coding | intron_variant,NMD_transcript_variant;intron_variant |
| Known Loci | *FAM49A* | 2 | 16705500 | rs140385594 | Intronic | Transcript | lncRNA | intron_variant,non_coding_transcript_variant |
| Known Loci | *FAM49A* | 2 | 16733525 | rs5007483 | Exonic | Transcript | protein_coding;lncRNA | 3_prime_UTR_variant;downstream_gene_variant;intron_variant,non_coding_transcript_variant |
| Known Loci | *HYAL2* | 3 | *50281058* | rs11434969 | *intronic* | Transcript | protein_coding;retained_intron;protein_coding_CDS_not_defined;misc_RNA | intron_variant;downstream_gene_variant |
| Known Loci | *ADAMTS9* | 3 | *64627602* | rs6445418 | *intronic* | Transcript;RegulatoryFeature | protein_coding;retained_intron;misc_RNA;enhancer | intron_variant;intron_variant,non_coding_transcript_variant;regulatory_region_variant |
| Known Loci | *ADAMTS9* | 3 | *64630197* | rs9838262 | *intronic* | Transcript | protein_coding;retained_intron;misc_RNA | intron_variant;intron_variant,non_coding_transcript_variant |
| Known Loci | *FILIP1L* | 3 | 99680753 | rs11290599 | Intronic | Transcript;RegulatoryFeature | protein_coding;nonsense_mediated_decay;protein_coding_CDS_not_defined;miRNA;enhancer | intron_variant;intron_variant,NMD_transcript_variant;intron_variant,non_coding_transcript_variant;downstream_gene_variant;regulatory_region_variant |
| Known Loci | *TP63* | 3 | 189544776 | rs75834417 | Intronic | Transcript | protein_coding;miRNA;retained_intron | intron_variant;upstream_gene_variant;intron_variant,non_coding_transcript_variant |
| Known Loci | *TP63* | 3 | 189549082 | rs79792381 | Intronic | Transcript | protein_coding;miRNA;retained_intron | intron_variant;downstream_gene_variant;intron_variant,non_coding_transcript_variant |
| Known Loci | *TP63* | 3 | 189551340 | rs55660938 | Intronic | Transcript;RegulatoryFeature | protein_coding;miRNA;retained_intron;open_chromatin_region | intron_variant;downstream_gene_variant;intron_variant,non_coding_transcript_variant;regulatory_region_variant |
| Known Loci | *NNT* | 5 | 43856675 | rs34871596 | | - | - | intergenic_variant |
| Known Loci | *NNT* | 5 | 43862510 | rs34378343 | | - | - | intergenic_variant |
| Known Loci | *GMDS* | 6 | 1944939 | rs11242726 | Intronic | Transcript | protein_coding;protein_coding_CDS_not_defined | intron_variant;intron_variant,non_coding_transcript_variant |

| | | | | | | | | |
|---|---|---|---|---|---|---|---|---|
| Known Loci | *GMDS* | 6 | 1960651 | rs140236874 | Intronic | Transcript | protein_coding;retained_intron;protein_coding_CDS_not_defined | intron_variant;intron_variant,non_coding_transcript_variant |
| Known Loci | ZFHX4 | 8 | 77498222 | rs144305105 | Intronic | Transcript | lncRNA | intron_variant,non_coding_transcript_variant |
| Known Loci | *DCAF4L2* | 8 | 88946414 | rs5893033 | | - | - | intergenic_variant |
| Known Loci | *RIPK2* | 8 | 90232609 | rs141180188 | | - | - | intergenic_variant |
| Known Loci | *VIRMA* | 8 | 95503542 | rs58089312 | Intronic | Transcript | protein_coding;nonsense_mediated_decay;retained_intron | intron_variant;intron_variant,NMD_transcript_variant;intron_variant,non_coding_transcript_variant;downstream_gene_variant |
| Known Loci | *VIRMA* | 8 | 95663822 | rs10089502 | Intronic | Transcript | protein_coding;retained_intron;nonsense_mediated_decay;misc_RNA | intron_variant;intron_variant,non_coding_transcript_variant;intron_variant,NMD_transcript_variant;upstream_gene_variant |
| Known Loci | *VIRMA* | 8 | 95739642 | rs67763258 | Intronic | Transcript | protein_coding;nonsense_mediated_decay | intron_variant;intron_variant,NMD_transcript_variant |
| Known Loci | *LINC00824* | 8 | 129699660 | rs7839784 | Intronic | Transcript | lncRNA | intron_variant,non_coding_transcript_variant |
| Known Loci | *LINC00824* | 8 | 129703990 | rs35379093 | Intronic | Transcript | lncRNA | intron_variant,non_coding_transcript_variant |
| Known Loci | *LINC00824* | 8 | 129911532 | rs10631848 | Intronic | Transcript;RegulatoryFeature | lncRNA;enhancer | intron_variant,non_coding_transcript_variant;regulatory_region_variant |
| Known Loci | *LINC00824* | 8 | 129950399 | rs1372450 | Intronic | Transcript;RegulatoryFeature | lncRNA;open_chromatin_region | intron_variant,non_coding_transcript_variant;regulatory_region_variant |
| Known Loci | *GADD45G* | 9 | 92210595 | rs7024024 | Intronic | Transcript | lncRNA | downstream_gene_variant;intron_variant,non_coding_transcript_variant |
| Known Loci | *GADD45G* | 9 | 92211862 | rs10429497 | Intronic | Transcript | lncRNA | downstream_gene_variant;intron_variant,non_coding_transcript_variant |
| Known Loci | *PTCH1* | 9 | 98225056 | rs28377268 | Intronic | Transcript;MotifFeature;RegulatoryFeature | nonsense_mediated_decay;promoter;protein_coding;protein_coding_CDS_not_defined;retained_intron | TF_binding_site_variant;intron_variant,NMD_transcript_variant;regulatory_region_variant;intron_variant;upstream_gene_variant;downstream_gene_variant |
| Known Loci | *FOXE1* | 9 | 100508754 | rs541315146 | Intronic | Transcript | lncRNA | downstream_gene_variant;intron_variant,non_coding_transcript_variant |
| Known Loci | *FOXE1* | 9 | 100619090 | rs35060625 | | Transcript | lncRNA;protein_coding | upstream_gene_variant;downstream_gene_variant |
| Known Loci | *NR6A1* | 9 | 127369279 | rs77346093 | Intronic | Transcript | misc_RNA;protein_coding | upstream_gene_variant;intron_variant |
| Known Loci | *VAX1* | 10 | 118831210 | rs10886037 | Intronic | RegulatoryFeature;Transcript | enhancer;protein_coding | regulatory_region_variant;intron_variant |
| Known Loci | *VAX1* | 10 | 118836682 | rs11197887 | Intronic | Transcript | protein_coding | intron_variant |
| Known Loci | *VAX1* | 10 | 118850264 | rs17095834 | Intronic | RegulatoryFeature;Transcript | open_chromatin_region;protein_coding | regulatory_region_variant;intron_variant |
| Known Loci | *LGR5* | 12 | 71967309 | rs67687642 | Intronic | Transcript | protein_coding;retained_intron | intron_variant;downstream_gene_variant;intron_variant,non_coding_transcript_variant |
| Known Loci | *CLYBL* | 13 | 100512107 | rs9557318 | Intronic | RegulatoryFeature;Transcript | enhancer;lncRNA;nonsense_mediated_decay;protein_coding | regulatory_region_variant;downstream_gene_variant;intron_variant,non_coding_transcript_variant;upstream_gene_variant;intron_variant,NMD_transcript_variant;intron_variant |
| Known Loci | *PAX9* | 14 | 37227795 | rs555814798 | Intronic | Transcript | protein_coding;retained_intron | intron_variant;intron_variant,non_coding_transcript_variant |
| Known Loci | *PAX9* | 14 | 37458914 | rs10483482 | Intronic | Transcript | protein_coding;protein_coding_CDS_not_defined | intron_variant;intron_variant,non_coding_transcript_variant |
| Known Loci | *PAX9* | 14 | 37464217 | rs75088305 | Intronic | Transcript | protein_coding;protein_coding_CDS_not_defined | intron_variant;intron_variant,non_coding_transcript_variant |
| Known Loci | *PAX9* | 14 | 37514564 | rs10150336 | Intronic | Transcript | protein_coding;protein_coding_CDS_not_defined | intron_variant;intron_variant,non_coding_transcript_variant |
| Known Loci | *LINC00640* | 14 | 51856064 | rs144433632 | Intronic | Transcript | lncRNA | downstream_gene_variant;intron_variant,non_coding_transcript_variant;upstream_gene_variant |
| Known Loci | *GSC* | 14 | 95378540 | rs61665130 | | - | - | intergenic_variant |
| Known Loci | *UBL7-AS1* | 15 | 74759689 | rs559701024 | Intronic | Transcript | lncRNA | downstream_gene_variant;intron_variant,non_coding_transcript_variant |
| Known Loci | *CREBBP* | 16 | 3963187 | rs9923088 | | - | - | intergenic_variant |
| Known Loci | *NTN1* | 17 | 8930279 | rs35023427 | Intronic | Transcript | protein_coding | intron_variant |

| | | | | | | | | |
|---|---|---|---|---|---|---|---|---|
| Known Loci | *NTN1* | 17 | 8943063 | rs9911652 | Intronic | Transcript | protein_coding | intron_variant |
| Known Loci | *WNT9B* | 17 | 44993128 | rs4968248 | | Transcript | lncRNA | downstream_gene_variant;upstream_gene_variant |
| Known Loci | *WNT9B* | 17 | 45005703 | rs3785888 | Intronic | Transcript | nonsense_mediated_decay;protein_coding;protein_coding_CDS_not_defined;retained_intron | intron_variant,NMD_transcript_variant;upstream_gene_variant;intron_variant;intron_variant,non_coding_transcript_variant |
| Known Loci | *WNT9B* | 17 | 45042079 | rs3760377 | Exonic;Intronic | MotifFeature;Transcript;RegulatoryFeature | nonsense_mediated_decay;protein_coding;protein_coding_CDS_not_defined;TF_binding_site | TF_binding_site_variant;3_prime_UTR_variant,NMD_transcript_variant;intron_variant,NMD_transcript_variant;intron_variant;downstream_gene_variant;non_coding_transcript_exon_variant;regulatory_region_variant |
| Known Loci | *NOG* | 17 | 54776955 | rs227727 | | - | - | intergenic_variant |
| Known Loci | *RHPN2* | 19 | 33515581 | rs145403118 | Intronic | Transcript | nonsense_mediated_decay;protein_coding;retained_intron | intron_variant,NMD_transcript_variant;intron_variant;intron_variant,non_coding_transcript_variant |
| Known Loci | *RHPN2* | 19 | 33517152 | rs28570619 | Intronic | MotifFeature;RegulatoryFeature;Transcript | enhancer;nonsense_mediated_decay;protein_coding;retained_intron | TF_binding_site_variant;regulatory_region_variant;intron_variant,NMD_transcript_variant;intron_variant;intron_variant,non_coding_transcript_variant |
| Known Loci | *RHPN2* | 19 | 33525145 | rs10409307 | Intronic | Transcript | nonsense_mediated_decay;protein_coding | intron_variant,NMD_transcript_variant;intron_variant |
| Known Loci | *RHPN2* | 19 | 33545042 | rs12462348 | Intronic | RegulatoryFeature;Transcript | enhancer;nonsense_mediated_decay;processed_pseudogene;protein_coding | regulatory_region_variant;intron_variant,NMD_transcript_variant;downstream_gene_variant;intron_variant |
| Known Loci | *MAFB* | 20 | 39265737 | rs6029242 | | - | - | intergenic_variant |
| Known Loci | *MAFB* | 20 | 39270795 | rs6124259 | | RegulatoryFeature | enhancer | regulatory_region_variant;intergenic_variant |
| Known Loci | *MAFB* | 20 | 39272739 | rs4812449 | | RegulatoryFeature | enhancer | regulatory_region_variant;intergenic_variant |
| Known Loci | *MAFB* | 20 | 39274534 | rs71193616 | | - | - | intergenic_variant |
| Known Loci | *ERG* | 21 | 40031924 | rs9647180 | Intronic | Transcript | protein_coding;protein_coding_CDS_not_defined;retained_intron | intron_variant;intron_variant,non_coding_transcript_variant |
| Known Loci | *ERG* | 21 | 40034216 | rs59685837 | | Transcript | protein_coding;protein_coding_CDS_not_defined;retained_intron | upstream_gene_variant |
| Known Loci | *ARVCF* | 22 | 19972118 | rs9606203 | Intronic | MotifFeature;RegulatoryFeature;Transcript | promoter;protein_coding;protein_coding_CDS_not_defined;retained_intron | TF_binding_site_variant;regulatory_region_variant;intron_variant;intron_variant,non_coding_transcript_variant;upstream_gene_variant |
| Known Loci | *ARHGAP8* | 22 | 45150659 | rs5765956 | Intronic | Transcript | nonsense_mediated_decay;protein_coding;retained_intron | intron_variant,NMD_transcript_variant;intron_variant;intron_variant,non_coding_transcript_variant |

Supplementary Table 6. Functional Annotation of the lead SNPs by Haploreg V4.2 and GTEx database

| *Classification* | *Locus Name* | Chromosome | Position(b37) | **rsid** | **Ref** | **Alt** | **Promoter histone marks** | **Enhancer histone marks** | **DNAse** | **Motifs changed** | Enhancer_Encode | Promoter_Encode | eQTL_GTEx | sQTL_GTEx |
|---|---|---|---|---|---|---|---|---|---|---|---|---|---|---|
| Novel | *NTRK1* | 1 | 156839414 | rs4661229 | G | A | | 10 tissues | PLCNT | | | | MRPL24 | *MRPL24;RRNAD1;ISG20L2* |
| Novel | *HMGCR* | 2 | 219817037 | rs13011262 | | | | | | | | | GMPPA | |
| Novel | *CALD1* | 7 | 134845630 | rs4732060 | T | C | | 15 tissues | 25 tissues | Foxo_2,Nanog_disc2,Pou2f2_disc1,Pou2f2_known5,Pou3f3,Pou5f1_disc2,Sox_10,Sox_13,Sox_14,Sox_18,Sox_19,Sox_2,Sox_4,Sox_7 | NKEK;HepG2;HMEC | | CALD1;RP11-134L10.1;C7orf49 | *TMEM140;CALD1* |
| Novel | *CALD1* | 7 | 134877068 | rs10488465 | T | C | | | | GATA,Pou2f2 | | | CALD1;RP11-134L10.1 | *CALD1* |
| Novel | *SHH* | 7 | 156026696 | rs4716972 | G | A | | BRN | | BDP1,CTCF,MAZR,MZF1::1-4,Myc,RXRA | | | | |
| Novel | *NRG1* | 8 | 32493817 | rs17645417 | T | C | | ESDR, ESC, BRN | ESDR,GI | HDAC2,NRSF | | | | |
| Novel | *PRICKLE1* | 12 | 42488641 | rs12817499 | T | C | | | | Foxd3,Foxp1,HDAC2,Pou2f2,p300 | | | PRICKLE1 | |
| Novel | *ALX1* | 12 | 85585770 | rs565838209 | | | | | | | | | | |
| Novel | *SOX9* | 17 | 72283506 | rs62069766 | G | T | | BRST, SKIN | IPSC,BRST,BRST | BAF155,CTCF,Myc,YY1,Znf143 | | | | |
| Novel | *LINC00320* | 21 | 20793565 | rs13052576 | A | G | | 4 tissues | ESC | HMG-IY | | | | |
| Novel | *RUNX1* | 21 | 34831549 | rs2246738 | G | A | | FAT, BLD | MUS,MUS,SKIN | AP-1,SETDB1 | | | | |
| Novel | *MYH9* | 22 | 36288285 | rs5756130 | C | T | | 5 tissues | 6 tissues | Ets,NF-I,Pax-3,TCF12 | | | | |
| Known SNP | *PAX7* | 1 | 18972776 | rs9439713 | G | A | | 10 tissues | ESDR,ESC | BATF,CIZ,Foxa,Irf,NF-kappaB | | | | |
| Known SNP | *ARHGAP29* | 1 | 94558110 | rs66515264 | G | T | BLD, MUS | 17 tissues | 20 tissues | CCNT2,NF-kappaB,PLAG1,RREB-1,RXRA,Spz1,ZNF219,Zfp281,Zfp740 | HUVEC,NHLF | | ARHGAP29 | |
| Known SNP | *IRF6* | 1 | 209989092 | rs72741048 | A | T | SKIN | 6 tissues | 4 tissues | Arid3a,Cdx,HMG-IY,Mef2,SIX5,TATA | HMEC,NHEK | | IRF6,DIEXF | IRF6,C1orf74 |
| Known SNP | *IRF6* | 1 | 209989270 | rs642961 | A | G | BRST, SKIN | 11 tissues | 5 tissues | | HMEC,NHEK | | IRF6,DIEXF,C1orf74 | |
| Known SNP | *IRF6* | 1 | 209992501 | rs1109430 | G | A | | 4 tissues | ESDR,GI | | | | TRAF3IP3,DIEXF | SERTAD4-AS1 |
| Known SNP | *IRF6* | 1 | 210048819 | rs2064163 | G | T | BRN, BONE | 13 tissues | BRN,LNG | Nanog,Pou5f1,Sox,TCF4 | HMEC,HSMM,NHEK | | IRF6,C1orf74,SERTAD4-AS1 | |
| Known SNP | *FAM49A* | 2 | 16713395 | rs6745357 | C | G | | KID | | Sox | | | AC104623.2,RP11-542H15.1 | |
| Known SNP | *ZFHX4* | 8 | 77496113 | rs11993273 | A | G | | STRM, SKIN | | Foxa,Irf | | | | |
| Known SNP | LINC00824 | 8 | 129964873 | rs17242358 | G | A | | | | | | | | |
| Known SNP | LINC00824 | 8 | 129976136 | rs55658222 | G | A | | | | Pou5f1,Sox | | | | |
| Known SNP | *VAX1* | 10 | 118846294 | rs10886040 | C | G | | | SKIN,CRVX,BRST | AP-1,Irf,PU.1,ZEB1 | | | RP11-539I5.1,SHTN1 | |
| Known SNP | *SPRY2* | 13 | 80679302 | rs11841646 | T | A | | | | Arid3a,Esx1,HMG-IY,Hoxa3,Hoxa5,Hoxa7,Hoxb3,Hoxb4,Hoxb6,Hoxc6,Nobox,Pax7,Pou2f2,Pou3f2,Pou4f3,Prrx1,Prrx2,Zfp187 | | | | |
| Known SNP | *LOC283553* | 14 | 51855553 | rs4901116 | G | T | | MUS | BLD,MUS | Irf,STAT,p300 | | | RP11-255G12.2 | LINC00640 |
| Known SNP | *NTN1* | 17 | 8947708 | rs12944377 | T | C | | LIV | | Pax-4,TCF12 | | | NTN1 | |
| Known SNP | *NOG* | 17 | 54773238 | rs227731 | T | G | | | | Dobox4 | | | NOG | |
| Known SNP | *MAFB* | 20 | 39281629 | rs6102085 | G | A | | ESDR, STRM, MUS | KID | Bcl6b,ELF1,Ets,Foxa,Myf,PU.1,SPIB,STAT,TATA,p300 | | | | |
| Known SNP | *ARVCF* | 22 | 19976845 | rs756653 | G | A | | 7 tissues | BLD,BLD | NRSF,Sin3Ak-20 | | | ARVCF,COMT,TANGO2 | ARVCF,COMT |
| Known Loci | *PAX7* | 1 | 18957297 | rs569017380 | | | | | | | | | | |
| Known Loci | *GRHL3* | 1 | 24667423 | rs12568609 | G | A | | 9 tissues | ESDR,ESDR,IPSC | E2A,LBP-9,NRSF,ZEB1 | | | NIPAL3,RCAN3,STPG1 | STPG1 |
| Known Loci | *FGGY* | 1 | 59421811 | rs35110579 | A | G | | 10 tissues | ESDR,HRT,MUS | | | | HSD52 | |
| Known Loci | *FGGY* | 1 | 59422001 | rs11577647 | T | C | | 8 tissues | ESDR | | | | HSD52 | |
| Known Loci | *ARHGAP29* | 1 | 94601200 | rs138088826 | C | CG | | | | EWSR1-FLI1,NF-AT1 | | | RP11-148B18.1,ABCD3 | |
| Known Loci | *ARHGAP29* | 1 | 94606077 | rs7551877 | G | A | | | | HDAC2,RXRA,XBP-1 | | | | |
| Known Loci | *ARHGAP29* | 1 | 94615394 | rs11290844 | AT | A | | ESDR, ESC | | HMG-IY,Mef2 | | | ABCD3,ARHGAP29 | |
| Known Loci | *IRF6* | 1 | 209894822 | rs201481822 | A | AAG | | 8 tissues | | Evi-1,FAC1,Foxa,Foxd3,Foxp1,HDAC2,HMG-IY,Irf,NF-kappaB,Otx2,Pax-4,Zfp105,p300 | | | | |
| Known Loci | *IRF6* | 1 | 209913473 | rs12120361 | T | G | | | | CHOP::CEBPalpha | | | TRAF3IP3,SYT14,IRF6,ADORA2BP1 | TRAF3IP3 |
| Known Loci | *IRF6* | 1 | 209943893 | rs139818084 | GGTGT | G | 9 tissues | 15 tissues | 5 tissues | RREB-1,TEF-1 | HUVEC | | | |
| Known Loci | *IRF6* | 1 | 209974232 | rs622832 | A | C | LIV, HRT | 5 tissues | BRST,BRST,SKIN | Zfx | HUVEC,HMEC | | C1orf74,DIEXF,SYT14,TRAF3IP3 | |
| Known Loci | *IRF6* | 1 | 209982738 | rs17015255 | A | G | | 4 tissues | BRST,SKIN,BRST | CDP,Sin3Ak-20 | HMEC,NHEK | | DIEXF,TRAF3IP3 | SERTAD4-AS1 |
| Known Loci | *IRF6* | 1 | 210043322 | rs3028171 | A | 23-mer | | 9 tissues | SKIN,MUS | | HMEC,NHEK | | | |
| Known Loci | *IRF6* | 1 | 210248548 | rs4844943 | C | T | | ESC, IPSC, BRN | BRN | ATF3,TAL1 | | | | SERTAD4-AS1 |
| Known Loci | *IRF6* | 1 | 210253684 | rs10863809 | G | T | | | | Crx,GATA,Nrf1 | | | | SERTAD4-AS1 |
| Known Loci | *IRF6* | 1 | 210301331 | rs7516554 | C | T | | | | CHOP::CEBPalpha,E2F,YY1 | | | IRF6,TRAF3IP3 | |
| Known Loci | *IRF6* | 1 | 210352641 | rs2485896 | C | G | | | | | | | IRF6,DIEXF,SERTAD4-AS1,SYT14 | |
| Known Loci | *IRF6* | 1 | 210355896 | rs12087757 | G | A | | | | Myc,Pax-5 | | | IRF6,LAMB3,SERTAD4-AS1 | IRF6,C1orf74 |
| Known Loci | *IRF6* | 1 | 210372114 | rs2494175 | A | G,T | | SKIN | SKIN | | | | | |
| Known Loci | *TAF1B* | 2 | 9967620 | rs528634626 | | | | | | | | | | |
| Known Loci | *TAF1B* | 2 | 10028440 | rs71389302 | A | ATT | | | | Foxp1,HDAC2,Zfp105 | | | | |
| Known Loci | *FAM49A* | 2 | 16705500 | rs140385594 | T | TGTGA | | ESDR, HRT, LNG | ESDR | FAC1, Foxp1,GR,Zfp105 | | | | |
| Known Loci | *FAM49A* | 2 | 16733525 | rs5007483 | G | T | | | | RFX5 | | | AC104623.2,RP11-542H15.1 | |
| Known Loci | *HYAL2* | 3 | 50281058 | rs11434969 | G | GA | | | | HDAC2,HMG-IY,PRDM1,Pax-4,Zfp105,p300 | | | HYAL1;NAT6;HYAL3;CYB561D2;ZMYND10;AMT;RASSF1;CACNA2D2 | *HYAL1;CISH* |
| Known Loci | *ADAMTS9* | 3 | 64627602 | rs6445418 | G | C,T | | 9 tissues | ESDR,VAS | | | | ADAMTS9-AS1;ATXN7 | *ADAMTS9-AS1;ADAMTS9* |
| Known Loci | *ADAMTS9* | 3 | 64630197 | rs9838262 | A | G | | | | T3R | | | ADAMTS9-AS1 | *ADAMTS9-AS1;ADAMTS9* |
| Known Loci | *FILIP1L* | 3 | 99680753 | rs11290599 | AG | A | | 9 tissues | | Egr-1,GR | | | CMSS1,COL8A1,FILIP1L,LNP1,RP11-383I23.2,TBC1D23,TMEM30CP | |
| Known Loci | *TP63* | 3 | 189544776 | rs75834417 | A | C | | 5 tissues | SKIN | Foxm1,STAT | HMEC,NHEK | | | |
| Known Loci | *TP63* | 3 | 189549082 | rs79792381 | C | T | | BRST, SKIN | | CDP,Evi-1,FXR,Fox,Foxa,Foxc1,Foxd3,Foxf1,Foxj1,Foxj2,Foxk1,Foxp1,HDAC2,HNF1,Nkx6-2,PLZF,Sox | NHEK | | | |
| Known Loci | *TP63* | 3 | 189551340 | rs55660938 | C | T | SKIN | BRST, SKIN | SKIN,SKIN | CEBPB,NF-Y | HMEC,NHEK | | | |

| | | | | | | | | | | | | | |
|---|---|---|---|---|---|---|---|---|---|---|---|---|---|
| Known Loci | *NNT* | 5 | 43856675 | rs34871596 | G | A | | HRT, LNG | | Foxp1,RREB-1 | | | |
| Known Loci | *NNT* | 5 | 43862510 | rs34378343 | T | C | | | | FAC1,Foxp1,GR,Zfp105 | | NNT,NNT-AS1,PAIP1,TMEM267 | NNT-AS1 |
| Known Loci | *GMDS* | 6 | 1944939 | rs11242726 | G | A | | | | BHLHE40,DMRT2,Pax-6,TCF4 | | | |
| Known Loci | *GMDS* | 6 | 1960651 | rs140236874 | G | GA | | SKIN | PLCNT | Cdx,FAC1,Foxa,Foxd3,Foxp1,HDAC2,Ik-2,NF-AT,Pax-4,Pou2f2,Pou3f2,STAT,TATA,TEF,Zfp105 | NHEK | | |
| Known Loci | *ZFHX4* | 8 | 77498222 | rs144305105 | A | ATTATT | | | | Cdx2,Dbx1,Evi-1,Fox,Foxa,HDAC2,Hlx1,Hoxa10,Hoxa9,Hoxb13,Hoxb9,Hoxd10,Lhx3,Ncx,PLZF,Pou3f4,Sox,Zfp105 | | | |
| Known Loci | *DCAF4L2* | 8 | 88946414 | rs5893033 | G | GA | | | | Foxo,HDAC2,Ik-1,Mef2,RBP-Jkappa,Sox | | | |
| Known Loci | *RIPK2* | 8 | 90232609 | rs141180188 | C | CT | | | | Foxm1,Pou3f2,Sox | | | |
| Known Loci | VIRMA | 8 | 95503542 | rs58089312 | C | CA | | | | Hoxa4,Zfp105 | | | |
| Known Loci | VIRMA | 8 | 95663822 | rs10089502 | T | C | | | | Maf,Pou5f1,Sox | | ESRP1,DPY19L4,RP11-22C11.2 | |
| Known Loci | VIRMA | 8 | 95739642 | rs67763258 | G | T | | ADRL, BLD | | HMG-IY,Mef2 | GM12878 | DPY19L4,ESRP1 | DPY19L4 |
| Known Loci | LINC00824 | 8 | 129699660 | rs7839784 | T | C | | BLD | | AIRE,Foxa,SF1,p300 | | | |
| Known Loci | LINC00824 | 8 | 129703990 | rs35379093 | T | C | | ESC, IPSC | | Cdx,Foxp1,PLZF | | | |
| Known Loci | LINC00824 | 8 | 129911532 | rs10631848 | G | 11-mer | | | 4 tissues | | HSMM | | |
| Known Loci | LINC00824 | 8 | 129950399 | rs1372450 | C | T | | STRM, MUS | MUS | GR,PTF1-beta | HSMM | | |
| Known Loci | *GADD45G* | 9 | 92210595 | rs7024024 | A | T | | | | PLZF,Sox | | UNQ6494 | |
| Known Loci | *GADD45G* | 9 | 92211862 | rs10429497 | G | A | | | ADRL | | | UNQ6494,GADD45G | |
| Known Loci | *PTCH1* | 9 | 98225056 | rs28377268 | G | T | 17 tissues | 14 tissues | 46 tissues | AP-1,Egr-1,GR,MAZ,MZF1::1-4,Pou2f2,SEF-1,SP1,STAT,Sp4,TATA,TFII-I,ZNF263,Zfp281,p300 | GM12878,K562, HUVEC,HSMM, NHLF NHEK | PTCH1 | |
| Known Loci | *FOXE1* | 9 | 100508754 | rs541315146 | | | | | | | | | |
| Known Loci | *FOXE1* | 9 | 100619090 | rs35060625 | TCTC | T | | | | AP-3,CEBPG | | FOXE1,KRT18P13,NANS,PTCSC2,RP11-244N9.4,TRMO,XPA | TRMO |
| Known Loci | *NR6A1* | 9 | 127369279 | rs77346093 | T | C | | | | Maf | | | |
| Known Loci | *VAX1* | 10 | 118831210 | rs10886037 | A | T | | | | Mef2,p300 | | RP11-539I5.1,SHTN1 | |
| Known Loci | *VAX1* | 10 | 118836682 | rs11197887 | T | A | | | | HDAC2,Myc,Nanog,Nkx2 | | RP11-539I5.1,SHTN1 | |
| Known Loci | *VAX1* | 10 | 118850264 | rs17095834 | A | G | | | | EWSR1-FLI1,Ets,Foxo,Irf,p300 | | | |
| Known Loci | *LGR5* | 12 | 71967309 | rs67687642 | AT | A | | BLD | | Dbx1,HMG-IY,Pou2f2 | | TMEM19 | |
| Known Loci | *CLYBL* | 13 | 100512107 | rs9557318 | G | A | | 4 tissues | BLD,BLD,BLD | STAT | | | |
| Known Loci | *PAX9* | 14 | 37227795 | rs555814798 | | | | | | | | | |
| Known Loci | *PAX9* | 14 | 37458914 | rs10483482 | C | T | | | MUS | | | | |
| Known Loci | *PAX9* | 14 | 37464217 | rs75088305 | G | A | | IPSC | | Ets,Irf,STAT,TATA,YY1,p300 | | | |
| Known Loci | *PAX9* | 14 | 37514564 | rs10150336 | A | T | | | | Dbx1,Hdx,PLZF,Pou2f2,TATA | | MIPOL1,RP11-356O9.2,SLC25A21,SLC25A21-AS1 | MIPOL1 |
| Known Loci | LINC00640 | 14 | 51856064 | rs144433632 | AAT | A | | | | Pax-4 | | TMX1 | |
| Known Loci | *GSC* | 14 | 95378540 | rs61665130 | T | TG | | | | Barhl1,Barx1,Barx2,CHX10,Dbx1,Dlx2,Dlx3,Dlx5,En-1,Esx1,Evi-1,Foxa,Foxo,Foxp1,Gbx1,Gbx2,HDAC2,HNF1,Hoxa3,Hoxa9,Hoxb3,Hoxb4,Hoxd10,Lhx4,Lhx8,Mef2,Msx-1,Msx2,Ncx,Nkx6-1,Nkx6-2,Pax-4,Pax-6,Pou2f2,Pou3f2,Pou3f4,Pou6f1,Prrx1,Prrx2 | | GSC | |
| Known Loci | UBL7-AS1 | 15 | 74759689 | rs559701024 | | | | | | | | | |
| Known Loci | *CREBBP* | 16 | 3963187 | rs9923088 | C | G | | | | PPAR,Pax-4 | | RP11-462G12.1 | |
| Known Loci | *NTN1* | 17 | 8930279 | rs35023427 | TA | T | GI | 5 tissues | 5 tissues | ERalpha-a,TR4 | | NTN1 | |
| Known Loci | *NTN1* | 17 | 8943063 | rs9911652 | C | T | | | | | | NTN1 | |
| Known Loci | *WNT9B* | 17 | 44993128 | rs4968248 | G | A | | BRST, BRN, SKIN | PLCNT | AP-2 | | GOSR2,KANSL1,LINC01974,LRRC37A17P,MYL4,RP11-156P1.3,RPRML,WNT3 | GOSR2,RP11-156P1.3 |
| Known Loci | *WNT9B* | 17 | 45005703 | rs3785888 | C | T | ESDR, GI | 16 tissues | GI,LNG | | HUVEC | ARL17A,ARL17B,GOSR2 | GOSR2,RP11-156P1.3,LRRC46,WNT3 |
| Known Loci | *WNT9B* | 17 | 45042079 | rs3760377 | G | A | | ESDR | | Maf,Mrg,NERF1a,Tgif1 | | ARL17A,ARL17B,GOSR2,KANSL1,LINC01974,LRRC37A17P,RP11-156P1.3,RPRML,WNT3 | GOSR2,RP11-156P1.3,WNT3 |
| Known Loci | *NOG* | 17 | 54776955 | rs227727 | A | T | 11 tissues | 14 tissues | 32 tissues | Crx,Dmbx1,Obox3,Otx2,Pitx3,Spz1 | HUVEC,HMEC, HSMM,NHEK, NHLF | NOG | |
| Known Loci | *RHPN2* | 19 | 33515581 | rs145403118 | CT | C | | 4 tissues | ESDR,PLCNT | LUN-1 | | GPATCH1,LRP3,RHPN2 | GPATCH1 |
| Known Loci | *RHPN2* | 19 | 33517152 | rs28570619 | G | A | LIV | LIV, GI, LNG | | Foxj2,Ik-2,Pitx2 | HepG2 | GPATCH1,RHPN2 | GPATCH1 |
| Known Loci | *RHPN2* | 19 | 33525145 | rs10409307 | A | G | | | | Myc,NRSF,PLAG1,Sp4,Zfx | | RHPN2,PDCD5,NUDT19,GPATCH1,FAAP24,CTD-2538C1.2,CEP89,CEBPA-AS1 | GPATCH1 |
| Known Loci | *RHPN2* | 19 | 33545042 | rs12462348 | C | T | | ESDR, GI, CRVX | GI,GI | Crx,Pitx2 | | CTD-2538C1.2,GPATCH1,NUDT19,PDCD5,RHPN2,WDR88 | GPATCH1 |
| Known Loci | *MAFB* | 20 | 39265737 | rs6029242 | T | C | | PANC | | | | | |
| Known Loci | *MAFB* | 20 | 39270795 | rs6124259 | G | A | | 5 tissues | ESDR,LIV | BDP1,Maf,NERF1a,Pbx3,Znf143 | | | |
| Known Loci | *MAFB* | 20 | 39272739 | rs4812449 | C | G | | 16 tissues | 6 tissues | CCNT2,GATA,SETDB1 | | | |
| Known Loci | *MAFB* | 20 | 39274534 | rs71193616 | G | GGGA | | ADRL, VAS | | ELF1,Egr-1,Ets,GATA,Irf,MZF1::1-4,PU.1,Pax-5,SP1,TATA | | | |
| Known Loci | *ERG* | 21 | 40031924 | rs9647180 | T | C | 6 tissues | 5 tissues | LIV | Ets,RXRA | | ERG | |
| Known Loci | *ERG* | 21 | 40034216 | rs59685837 | GA | G | 4 tissues | 8 tissues | 6 tissues | DMRT1,Irf,Zfp105 | | | |
| Known Loci | *ARVCF* | 22 | 19972118 | rs9606203 | C | A | 6 tissues | 20 tissues | 22 tissues | HNF4,RXRA,SIX5 | K562,HUVEC,HMEC,NHEK | ARVCF,CCDC188,COMT,TANGO2 | ARVCF,COMT |
| Known Loci | *ARHGAP8* | 22 | 45150659 | rs5765956 | C | T | IPSC, GI | 13 tissues | 7 tissues | CHD2,E2F,TATA | | ARHGAP8 | |

Supplementary Table 7. Novel loci containing cleft genes

| Phentotype | Locus Name | Chromsome | Position(b37) | rsid | GeneList (500kb flanked novel leadSN | Related Genes/Region | Syndromic (craniofacial phenoty | Animal Model_orofacial cleft (PMID) | Cranifacial pathway (PMID) |
|---|---|---|---|---|---|---|---|---|---|
| NSOC | *NTRK1* | 1 | 156809206 | rs4661229 | TSACC,RHBG,C1orf61,MIR9−1,MEF2D,IQGAP3,TTC24,APOA1BP,GPATCH4,HAPLN2,BCAN,NES,CRABP2, ISG20L2,RRNAD1,MRPL24,HDGF,PRCC,SH2D2A,NTRK1,INSRR,PEAR1,LRRC71,ARHGEF11,MIR765,ETV3L,ETV3,CYCSP52 | NTRK1 | cleft palate (PMID: 29421787) | | |
| NSCL/P | *HMGCR* | 2 | 220681758 | rs13011262 | ENC1,HEXB,GFM2,NSA2,FAM169A,GCNT4,ANKRD31,HMGCR,COL4A3BP,POLK,ANKDD1B | HMGCR | | 36320166 | 27488927 |
| NSOC | *CALD1* | 7 | 134530381 | rs4732060 | *AKR1B1,AKR1B10,AKR1B15,BPGM,CALD1,AGBL3,TMEM140,C7orf49,WDR91,MIR6509,STRA8,CNOT4* | 7q33 CNV | cleft palate (PMID: 29260337) | | |
| NSCL/P;NSCLP | *CALD1* | 7 | 134561819 | rs10488465 | *AKR1B1,AKR1B10,AKR1B15,BPGM,CALD1,AGBL3,TMEM140,C7orf49,WDR91,MIR6509,STRA8,CNOT4* | 7q33 CNV | cleft palate (PMID: 29260337) | | |
| NSOC;NSCL/P | *SHH* | 7 | 155819390 | rs4716972 | CNPY1,RBM33,SHH,LOC389602,LOC285889,LINC01006 | SHH | | 883,777,015,199,404 | 24590292 |
| NSOC;NSCL/P | *NRG1* | 8 | 32351333 | rs17645417 | NRG1,NRG1−IT1,NRG1−IT3 | | | | |
| NSCL/P | *PRICKLE1* | 12 | 42882443 | rs12817499 | GXYLT1,YAF2,ZCRB1,PPHLN1,PRICKLE1,LOC101927058 | *PRICKLE1* | | *24689077* | *2,468,907,735,809,770* |
| NSCLO | *ALX1* | 12 | 85979547 | rs565838209 | LRRIQ1,ALX1,RASSF9,MGAT4C | ALX1 | orofacial cleft (PMID: 35127681 | 35127681 | |
| NSCL/P | *SOX9* | 17 | 70279647 | rs62069766 | LOC102723505,LINC01152,LOC102723517,SOX9−AS1,LOC101928205,SOX9,LINC00673,LINC00511,SLC39A11 | SOX9 | Pierre Robin sequence (PMID: 1 | 11371614 | |
| NSOC;NSCL/P | *LINC00320* | 21 | 22165883 | rs13052576 | LINC00320,NCAM2,RNU6−67P | | | | |
| NSOC;NSCPO | *RUNX1* | 21 | 36203846 | rs2246738 | KCNE2,SMIM11,C21orf140,KCNE1,RCAN1,CLIC6,LINC00160,LOC100506385,RUNX1,RUNX1−IT1 | RUNX1 | | 19000669, 30046048,14516670,31171577,34931[illegible] | 19000669, 30046048 |
| NSCL/P | *MYH9* | 22 | 36684331 | rs5756130 | RBFOX2,APOL3,APOL4,APOL2,APOL1,MYH9,MIR6819,TXN2,FOXRED2,EIF3D,CACNG2,IFT27 | MYH9 | | PMC2740885 | |

Note:NSOC, non-syndromic orofacial cleft cases by combining NSCLP,NSCLO and NSCPO together; NSCL/P, non-syndromic cleft lip with or without palate (NSCLP&NSCLO); NSCLP, non-syndromic cleft lip and palate; NSCLO, non-syndromic cleft lip only; NSCPO, non-syndromic cleft palate only.

Supplementary Table 8. Heritabilty Estimated based on LD score regression by phenotypic group across population

| Population | Phenotype | H2 | 95%CI |
|---|---|---|---|
| Asian | NSOC | 0.12 | 0.087-0.15 |
| Asian | NSCL/P | 0.18 | 0.13-0.23 |
| Asian | NSCLP | 0.2 | 0.15-0.26 |
| Asian | NSCLO | 0.2 | 0.088-0.32 |
| Chinese | NSOC | 0.14 | 0.11-0.18 |
| Chinese | NSCL/P | 0.21 | 0.16-0.27 |
| Chinese | NSCLP | 0.19 | 0.14-0.25 |
| Chinese | NSCLO | 0.25 | 0.12-0.38 |
| Chinese | NSCPO | 0.26 | 0.19-0.33 |

Note: NSOC, non-syndromic orofacial cleft cases by combining NSCLP,NSCLO and NSCPO together; NSCL/P, non-syndromic cleft lip with or without palate (NSCLP&NSCLO); NSCLP, non-syndromic cleft lip and palate; NSCLO, non-syndromic cleft lip only; NSCPO, non-syndromic cleft palate only;95%CI, 95% confidence interval.

Supplementary Table 9. Genetic Correlation Analysis in Chinese and European Population

| Population | Phenotype_1 | Phenotype_2 | p-value | rg | 95%Lower_rg | 95%Upper_rg | se_rg | z | h2_obs | h2_obs_se | h2_int | h2_int_se | gcov_int | gcov_int_se |
|---|---|---|---|---|---|---|---|---|---|---|---|---|---|---|
| Chinese | NSCLO | NSCPO | 0.38 | 0.2 | -0.25 | 0.65 | 0.23 | 0.87 | 0.16 | 0.039 | 1.12 | 0.0094 | 0.27 | 0.0065 |
| | NSCLO | NSCLP | 1.92E-09 | 0.71 | 0.48 | 0.94 | 0.12 | 6 | 0.3 | 0.059 | 1.06 | 0.0094 | 0.12 | 0.0072 |
| | NSCPO | NSCLP | 0.65 | -0.05 | -0.26 | 0.17 | 0.11 | -0.45 | 0.32 | 0.052 | 1.05 | 0.0079 | 0.14 | 0.0055 |
| European | NSOC | BMI | 0.03 | 0.079 | 0.0076 | 0.15 | 0.04 | 2.17 | 0.21 | 0.007 | 1.03 | 0.033 | -0.013 | 0.0079 |
| | NSCL/P | BMI | 0.033 | 0.077 | 0.0063 | 0.15 | 0.04 | 2.14 | 0.21 | 0.007 | 1.03 | 0.033 | -0.015 | 0.0081 |
| | NSCLP | BMI | 0.04 | 0.072 | 0.0032 | 0.14 | 0.04 | 2.05 | 0.21 | 0.007 | 1.03 | 0.033 | -0.015 | 0.0081 |
| | NSCPO | BMI | 0.52 | -0.037 | -0.15 | 0.08 | 0.06 | -0.64 | 0.21 | 0.0072 | 1.03 | 0.034 | -0.0092 | 0.0083 |
| | NSOC | Fetal_BW | 0.79 | 0.014 | -0.087 | 0.11 | 0.05 | 0.26 | 0.098 | 0.0057 | 1.09 | 0.018 | -0.0033 | 0.0061 |
| | NSCL/P | Fetal_BW | 0.53 | 0.033 | -0.069 | 0.13 | 0.05 | 0.63 | 0.098 | 0.0057 | 1.09 | 0.018 | -0.0063 | 0.006 |
| | NSCLP | Fetal_BW | 0.49 | 0.036 | -0.065 | 0.14 | 0.05 | 0.7 | 0.098 | 0.0057 | 1.09 | 0.018 | -0.0042 | 0.0061 |
| | NSCPO | Fetal_BW | 0.44 | 0.064 | -0.099 | 0.23 | 0.08 | 0.77 | 0.096 | 0.0059 | 1.1 | 0.02 | -0.0019 | 0.0056 |
| | NSOC | Fetal_Effect | 0.33 | 0.076 | -0.077 | 0.23 | 0.08 | 0.98 | 0.03 | 0.003 | 1.04 | 0.01 | -0.0027 | 0.0054 |
| | NSCL/P | Fetal_Effect | 0.41 | 0.066 | -0.09 | 0.22 | 0.08 | 0.83 | 0.03 | 0.003 | 1.04 | 0.01 | -0.0056 | 0.0053 |
| | NSCLP | Fetal_Effect | 0.36 | 0.072 | -0.081 | 0.22 | 0.08 | 0.92 | 0.03 | 0.003 | 1.04 | 0.0099 | -0.0031 | 0.0053 |
| | NSCPO | Fetal_Effect | 0.38 | 0.11 | -0.13 | 0.35 | 0.12 | 0.89 | 0.029 | 0.003 | 1.05 | 0.01 | -0.0023 | 0.0047 |

Note: NSOC, non-syndromic orofacial cleft cases by combining NSCLP,NSCLO and NSCPO together; NSCL/P, non-syndromic cleft lip with or without palate (NSCLP&NSCLO); NSCLP, non-syndromic cleft lip and palate; NSCLO, non-syndromic cleft lip only; NSCPO, non-syndromic cleft palate only;Fetal_BW, Fetal birth weight.

Supplementary Table 10. Associations of Lead SNPs with Other Traits

| Group | SNP | Locus Name | Chromosome | Position(b37) | A1 | A2 | Trait | EFO | PMID | Ancestry | P_lowest | Beta | SE | N | Dataset |
|---|---|---|---|---|---|---|---|---|---|---|---|---|---|---|---|
| Orofacial clefts | rs9439713 | *PAX7* | 1 | 18972776 | A | G | Cleft lip with or without cleft palate;Orofacial clefts | HP_0000175;EFO_0003959;HP_0000202 | 28054174 | Mixed | 6E-13 | NA | NA | - | NHGRI-EBI_GWAS_Catalog |
| | rs66515264 | *ARHGAP29* | 1 | 94558110 | G | T | Cleft lip with or without cleft palate;Orofacial clefts | HP_0000175;EFO_0003959;HP_0000202 | 28054174 | Mixed | 4E-17 | NA | NA | - | NHGRI-EBI_GWAS_Catalog |
| | rs1109430 | *IRF6* | 1 | 209992501 | A | G | Orofacial clefts | HP_0000202 | 28054174 | Mixed | 1E-21 | NA | NA | - | NHGRI-EBI_GWAS_Catalog |
| | rs2064163 | *IRF6* | 1 | 210048819 | G | T | Nonsyndromic cleft lip with cleft palate;Cleft lip with or without cleft palate in Asian ancestry population;N | HP_0000175;EFO_0003959 | 28232668 | East Asian | 9E-19 | 0.2614 | 0.02954 | - | NHGRI-EBI_GWAS_Catalog;GRASP |
| | rs17242358 | *LINC00824* | 8 | 129964873 | A | G | Orofacial clefts | HP_0000202 | 28054174 | Mixed | 6E-37 | NA | NA | - | NHGRI-EBI_GWAS_Catalog |
| | rs55658222 | *LINC00824* | 8 | 129976136 | A | G | Cleft lip with or without cleft palate | HP_0000175;EFO_0003959 | 28054174 | Mixed | 8E-44 | NA | NA | - | NHGRI-EBI_GWAS_Catalog |
| | rs10886040 | *VAX1* | 10 | 118846294 | C | G | Orofacial clefts;Cleft lip with or without cleft palate | HP_0000202;HP_0000175;EFO_0003959 | 28054174 | Mixed | 8E-11 | NA | NA | - | NHGRI-EBI_GWAS_Catalog |
| | rs11841646 | *SPRY2* | 13 | 80679302 | A | T | Nonsyndromic cleft lip;Cleft lip with or without cleft palate;Orofacial clefts | EFO_0003959;HP_0000175;HP_0000202 | 22863734 | Mixed | 4.71E-11 | NA | NA | 2383 | GRASP;NHGRI-EBI_GWAS_Catalog |
| | rs9911652 | *NTN1* | 17 | 8943063 | C | T | Cleft lip with or without cleft palate | HP_0000175;EFO_0003959 | 28054174 | Mixed | 0.00000003 | NA | NA | - | NHGRI-EBI_GWAS_Catalog |
| | rs12944377 | *NTN1* | 17 | 8947708 | C | T | Cleft lip with or without cleft palate;Orofacial clefts | HP_0000175;EFO_0003959;HP_0000202 | 28054174 | Mixed | 8E-21 | NA | NA | - | NHGRI-EBI_GWAS_Catalog |
| | rs227731 | *NOG* | 17 | 54773238 | T | G | Cleft lip with or without cleft palate;Nonsyndromic cleft lip with cleft palate;Cleft lip;Orofacial clefts;Nonsy | HP_0000175;EFO_0003959;EFO_0004625 | 28054174;28232668 | Mixed;East Asian;European | 0.000000002 | NA | NA | - | NHGRI-EBI_GWAS_Catalog;dbGaP;GRASP |
| | rs6102085 | *MAFB* | 20 | 39281629 | A | G | Cleft lip with or without cleft palate in Asian ancestry population;Cleft lip with or without cleft palate | EFO_0003959 | 20436469 | Mixed | 1.22E-08 | NA | NA | 5589 | GRASP |
| Anthropometric Traits | rs3028171 | *IRF6* | 1 | 210043322 | A | C | Leg fat mass left;Leg fat mass right;Arm fat mass right;Arm fat mass left | - | UKBB | European | 2.526E-09 | -0.0142 | 0.002383 | 331275 | Neale-B_UKBB_EUR_2017 |
| | rs7516554 | *IRF6* | 1 | 210301331 | C | T | Hip circumference;Arm fat percentage right;Body mass index;Arm fat mass left;Arm fat mass right;Arm fa | EFO_0005093;EFO_0004340 | UKBB | European | 2.526E-10 | -0.01554 | 0.002457 | 336601 | Neale-B_UKBB_EUR_2017 |
| | rs2485896 | *IRF6* | 1 | 210352641 | C | G | Leg fat mass left;Arm fat mass left;Waist circumference;Body mass index;Leg fat mass right;Arm fat mass right;Trunk fat mass;Whole body fat mass;Arm fat percentage left;Hip circumference;Arm fat percentage right;Body fat percentage;Weight;Trunk fat percentage;Leg fat percentage left;Leg fat percentage right; | EFO_0004342 | UKBB | European | 4.573E-12 | -0.01704 | 0.002463 | 331275 | Neale-B_UKBB_EUR_2017 |
| | rs10488465 | *CALD1* | 7 | 134561819 | C | T | Impedance of arm left | - | UKBB | European | 1.605E-08 | 0.01057 | 0.001871 | 331292 | Neale-B_UKBB_EUR_2017 |
| | rs67763258 | *VIRMA* | 8 | 95739642 | G | T | Body mass index;Leg fat mass right;Leg fat mass left | EFO_0004340 | UKBB | European | 7.104E-09 | -0.0147 | 0.00254 | 336107 | Neale-B_UKBB_EUR_2017 |
| | rs1372450 | *LINC00824* | 8 | 129950399 | C | T | Height | EFO_0004339 | 25282103 | European | 0.000000033 | -0.017 | 0.003 | 252199 | GIANT_Height_EUR_2014 |
| | rs28377268 | *PTCH1* | 9 | 98225056 | G | T | Sitting height;Height;Trunk fat-free mass;Trunk predicted mass;Whole body fat-free mass;Whole body water mass;Leg fat-free mass right;Leg predicted mass right;Basal metabolic rate;Leg predicted mass left; Leg fat-free mass left;Arm predicted mass right;Arm predicted mass left;Arm fat-free mass right; Arm fat-free mass left;Birth weight;Impedance of leg right;Impedance of leg left;Comparative height size at age 10;Hip circumference;Impedance of whole body;Weight | EFO_0004339;EFO_0007777;EFO_0004344;EFO_000 | UKBB | European | 1.824E-53 | -0.04668 | 0.003032 | 336172 | Neale-B_UKBB_EUR_2017 |
| | rs4968248 | *WNT9B* | 17 | 44993128 | A | G | Sitting height | EFO_0004339 | UKBB | European | 7.176E-09 | 0.01114 | 0.001924 | 336172 | Neale-B_UKBB_EUR_2017 |
| | rs3760377 | *WNT9B* | 17 | 45042079 | A | G | Sitting height | EFO_0004339 | UKBB | European | 1.303E-08 | 0.0107 | 0.001882 | 336172 | Neale-B_UKBB_EUR_2017 |
| | rs227731 | *NOG* | 17 | 54773238 | T | G | Height;Comparative height size at age 10 | EFO_0004339 | 25282103;UKBB;28146470;237 | European;Mixed | 3.1E-18 | -0.026 | 0.003 | 252377 | GIANT_Height_EUR_2014;Neale-B_UKBB_EUR_2017;GIANT_Height_Mixed_2017;GIANT_Height_EUR_2017;GIANT_Height_EUR_2013 |
| | rs227727 | *NOG* | 17 | 54776955 | T | A | Height;Comparative height size at age 10 | EFO_0004339 | 25282103;UKBB;23754948 | European | 3.7E-18 | 0.027 | 0.003 | 252147 | GIANT_Height_EUR_2014;Neale-B_UKBB_EUR_2017;GIANT_Height_EUR_2013 |
| Blood cell | rs12120361 | *IRF6* | 1 | 209913473 | G | T | Red blood cell count | EFO_0004586 | 27863252 | European | 3.617E-09 | 0.02579 | 0.004371 | 173480 | Astle-W_Blood-Cell-Traits_EUR_2016 |
| | rs140385594 | *FAM49A* | 2 | 16705500 | - | GTGA | Mean platelet volume | EFO_0004586 | 27863252 | European | 5.34E-19 | NA | NA | 173480 | Astle-W_Blood-Cell-Traits_EUR_2016 |
| | rs6745357 | *FAM49A* | 2 | 16713395 | C | G | Mean platelet volume | EFO_0004586 | 27863252 | European | 1.561E-13 | -0.02941 | 0.003983 | 173480 | Astle-W_Blood-Cell-Traits_EUR_2016 |
| | rs5007483 | *FAM49A* | 2 | 16733525 | G | T | Mean platelet volume | EFO_0004586 | 27863252 | European | 1.741E-13 | -0.02983 | 0.004049 | 173480 | Astle-W_Blood-Cell-Traits_EUR_2016 |
| | rs28377268 | *PTCH1* | 9 | 98225056 | G | T | Neuroticism score | EFO_0007660 | UKBB | European | 7.101E-10 | -0.08667 | 0.01406 | 274108 | Neale-B_UKBB_EUR_2017 |
| | rs6029242 | *MAFB* | 20 | 39265737 | C | T | White blood cell count | EFO_0004586 | 27863252 | European | 1.784E-09 | 0.02238 | 0.00372 | 173480 | Astle-W_Blood-Cell-Traits_EUR_2016 |
| | rs6124259 | *MAFB* | 20 | 39270795 | A | G | White blood cell count;Monocyte count | EFO_0004586 | 27863252 | European | 4.565E-11 | 0.02451 | 0.003722 | 173480 | Astle-W_Blood-Cell-Traits_EUR_2016 |
| | rs4812449 | *MAFB* | 20 | 39272739 | C | G | Monocyte count;White blood cell count | EFO_0004586 | 27863252 | European | 2.287E-10 | -0.02312 | 0.003646 | 173480 | Astle-W_Blood-Cell-Traits_EUR_2016 |
| | rs9606203 | *ARVCF* | 22 | 19972118 | A | C | Plateletcrit;Platelet count;White blood cell count;Lymphocyte count;Monocyte count | EFO_0004586 | 27863252 | European | 1.819E-11 | 0.02715 | 0.00404 | 173480 | Astle-W_Blood-Cell-Traits_EUR_2016 |
| | rs756653 | *ARVCF* | 22 | 19976845 | G | A | Plateletcrit;Platelet count;White blood cell count;Lymphocyte count;Monocyte count | EFO_0004586 | 27863252 | European | 1.638E-11 | -0.02722 | 0.004042 | 173480 | Astle-W_Blood-Cell-Traits_EUR_2016 |
| Blood Pressure | rs4968248 | *WNT9B* | 17 | 44993128 | A | G | Systolic blood pressure | EFO_0006335 | UKBB | European | 1.269E-15 | -0.02016 | 0.002521 | 317754 | Neale-B_UKBB_EUR_2017 |
| | rs3760377 | *WNT9B* | 17 | 45042079 | A | G | Systolic blood pressure | EFO_0006335 | UKBB | European | 4.795E-09 | -0.01444 | 0.002466 | 317754 | Neale-B_UKBB_EUR_2017 |
| | rs9647180 | *ERG* | 21 | 40031924 | C | T | Diastolic blood pressure | EFO_0006336 | UKBB | European | 1.986E-10 | 0.01852 | 0.00291 | 317756 | Neale-B_UKBB_EUR_2017 |
| Bone | rs28377268 | *PTCH1* | 9 | 98225056 | G | T | Bone mineral density spine | EFO_0007701 | 26733130 | Mixed | 1E-11 | -0.088 | 0.01293 | - | NHGRI-EBI_GWAS_Catalog |
| | rs28570619 | *RHPN2* | 19 | 33517152 | A | G | Heel bone mineral density;Heel bone mineral density right;Heel bone mineral density left | EFO_0009270 | UKBB | European | 1.15E-42 | 0.1053 | 0.00769 | 194398 | Neale-B_UKBB_EUR_2017 |
| | rs9647180 | *ERG* | 21 | 40031924 | C | T | Heel bone mineral density | EFO_0009270 | UKBB | European | 6.462E-20 | 0.03366 | 0.003684 | 194398 | Neale-B_UKBB_EUR_2017 |
| Others | rs12568609 | *GRHL3* | 1 | 24667423 | A | G | Hair or balding pattern: pattern 4 | EFO_0007822 | UKBB | European | 4.01E-08 | -0.008989 | 0.001637 | 154988 | Neale-B_UKBB_EUR_2017 |
| | rs28377268 | *PTCH1* | 9 | 98225056 | G | T | Forced vital capacity;Forced vital capacity, best measure;Forced expiratory volume in 1-second | EFO_0004312;EFO_0004314 | UKBB | European | 2.16E-27 | -0.03455 | 0.003186 | 307638 | Neale-B_UKBB_EUR_2017 |
| | rs28377268 | *PTCH1* | 9 | 98225056 | G | T | Sensitivity or hurt feelings | EFO_0009599 | UKBB | European | 3.546E-11 | -0.01288 | 0.001946 | 327832 | Neale-B_UKBB_EUR_2017 |
| | rs28377268 | *PTCH1* | 9 | 98225056 | G | T | Worrier or anxious feelings | EFO_0009863 | UKBB | European | 4.745E-08 | -0.01064 | 0.001948 | 328717 | Neale-B_UKBB_EUR_2017 |
| | rs3760377 | *WNT9B* | 17 | 45042079 | A | G | Gene expression of GOSR2 in prefrontal cortex | GO_1901148 | 21909115 | European | 1E-52 | NA | NA | 69395 | GRASP |

Supplementary Table 11. Two Sample Mendelian randomization Analysis Between Maternal BMI and offspring orofacial clefts

| Phenotype | Method | No.SNP | Beta | Std.Error | p-value | OR(95%CI) |
|---|---|---|---|---|---|---|
| NSOC | MR Egger | 57 | 1.39 | 2.11 | 0.51 | 4.01(0.06-252.57) |
| **NSOC** | **Weighted median** | **57** | **1.83** | **1.08** | **0.089** | **6.24(0.76-51.32)** |
| NSOC | Inverse variance weighted | 57 | 0.46 | 0.74 | 0.53 | 1.59(0.37-6.74) |
| NSOC | Simple mode | 57 | 2.25 | 2.28 | 0.33 | 9.48(0.11-821.81) |
| NSOC | Weighted mode | 57 | 1.98 | 1.77 | 0.27 | 7.21(0.22-232.79) |
| NSCL/P | MR Egger | 57 | 2.48 | 2.36 | 0.3 | 11.93(0.12-1218.31) |
| **NSCL/P** | **Weighted median** | **57** | **2.06** | **1.17** | **0.079** | **7.87(0.79-78.7)** |
| NSCL/P | Inverse variance weighted | 57 | 0.56 | 0.82 | 0.49 | 1.76(0.35-8.79) |
| NSCL/P | Simple mode | 57 | 1.95 | 2.39 | 0.42 | 7.02(0.06-758.91) |
| NSCL/P | Weighted mode | 57 | 2.11 | 1.96 | 0.29 | 8.24(0.18-382.6) |
| NSCLP | MR Egger | 57 | 3.35 | 2.5 | 0.19 | 28.41(0.21-3823.31) |
| NSCLP | Weighted median | 57 | 1.4 | 1.28 | 0.27 | 4.04(0.33-49.25) |
| NSCLP | Inverse variance weighted | 57 | 0.52 | 0.87 | 0.55 | 1.68(0.3-9.23) |
| NSCLP | Simple mode | 57 | -0.68 | 2.57 | 0.79 | 0.51(0-78.03) |
| NSCLP | Weighted mode | 57 | 1.84 | 2.05 | 0.37 | 6.31(0.11-353.17) |
| NSCLO | MR Egger | 54 | 2.21 | 3.43 | 0.52 | 9.13(0.01-7629.86) |
| NSCLO | Weighted median | 54 | -0.1 | 1.75 | 0.96 | 0.91(0.03-27.87) |
| NSCLO | Inverse variance weighted | 54 | 0.17 | 1.18 | 0.89 | 1.18(0.12-11.93) |
| NSCLO | Simple mode | 54 | -0.52 | 3.45 | 0.88 | 0.59(0-513.91) |
| NSCLO | Weighted mode | 54 | -0.07 | 2.62 | 0.98 | 0.93(0.01-159.77) |

Note: NSOC, non-syndromic orofacial cleft cases by combining NSCLP,NSCLO and NSCPO together; NSCL/P, non-syndromic cleft lip with or without palate (NSCLP&NSCLO); NSCLP, non-syndromic cleft lip and palate; NSCLO, non-syndromic cleft lip only;OR, Odds ratio; 95%CI, 95% confidence interval.

Supplementary Table 12. Association Results of Training Dataset

| POFC1(Weight) | CHW_Axiom (Training) | Estimate | Std.Error | z_value | p-value |
|---|---|---|---|---|---|
| NSOC | NSOC | 0.18 | 0.071 | 2.5 | 0.012 |
| NSOC | NSCL/P | 0.18 | 0.071 | 2.45 | 0.014 |
| NSOC | NSCLP | 0.15 | 0.082 | 1.82 | 0.068 |
| NSOC | NSCLO | 0.18 | 0.098 | 1.81 | 0.07 |
| NSCL/P | NSOC | 0.2 | 0.072 | 2.8 | 0.0051 |
| NSCL/P | NSCL/P | 0.2 | 0.072 | 2.8 | 0.0051 |
| NSCL/P | NSCLP | 0.16 | 0.081 | 1.95 | 0.051 |
| NSCL/P | NSCLO | 0.22 | 0.1 | 2.25 | 0.025 |
| NSCLP | NSOC | 0.16 | 0.072 | 2.29 | 0.022 |
| NSCLP | NSCL/P | 0.16 | 0.072 | 2.24 | 0.025 |
| NSCLP | NSCLP | 0.12 | 0.082 | 1.45 | 0.15 |
| NSCLP | NSCLO | 0.2 | 0.1 | 2.02 | 0.044 |
| NSCLO | NSOC | 0.09 | 0.076 | 1.18 | 0.24 |
| NSCLO | NSCL/P | 0.1 | 0.076 | 1.3 | 0.19 |
| NSCLO | NSCLP | 0.08 | 0.086 | 0.89 | 0.37 |
| NSCLO | NSCLO | 0.11 | 0.11 | 0.97 | 0.33 |

Note: NSOC, non-syndromic orofacial cleft cases by combining NSCLP,NSCLO and NSCPO together; NSCL/P, non-syndromic cleft lip with or without palate (NSCLP&NSCLO); NSCLP, non-syndromic cleft lip and palate; NSCLO, non-syndromic cleft lip only;CHW, Western Chinese.

Supplementary Table 13. Association of PGS and Non-syndromic orofacial clefts by phenotypic group

| Dataset | Phenotype | Case/Control | Estimate | Std.Error | z_value | p-value | OR(95%CI) | R2_w_PGS | R2_wo_ | AUC(95%CI)_PGS | AUC(95%CI)_PGS&Sex |
|---|---|---|---|---|---|---|---|---|---|---|---|
| CHW_ZH | NSOC | 1620/947 | 0.085 | 0.043 | 1.98 | **0.047** | 1.09(1.00-1.19) | 0.12 | 0.12 | 0.52(0.5-0.55) | 0.57(0.54-0.59) |
| | NSCL/P | 762/947 | 0.016 | 0.052 | 0.31 | 0.76 | 1.02(0.92-1.13) | 0.16 | 0.16 | 0.51(0.48-0.54) | 0.60(0.57-0.63) |
| | NSCLO | 749/947 | 0.019 | 0.053 | 0.36 | 0.72 | 1.02(0.92-1.13) | 0.16 | 0.16 | 0.51(0.48-0.54) | 0.60(0.57-0.63) |
| | NSCPO | 858/947 | 0.13 | 0.049 | 2.73 | **0.0064** | 1.14(1.04-1.26) | 0.11 | 0.1 | 0.54(0.51-0.56) | 0.54(0.51-0.57) |
| CHE_Axiom&HSA | NSCL/P | 322/722 | 0.35 | 0.11 | 3.22 | **0.0013** | 1.43(1.15-1.77) | 0.66 | 0.65 | 0.61(0.58-0.65) | 0.87(0.85-0.90) |
| | NSCLP | 215/722 | 0.35 | 0.13 | 2.66 | **0.0078** | 1.41(1.10-1.82) | 0.65 | 0.65 | 0.61(0.56-0.65) | 0.87(0.84-0.91) |
| | NSCLO | 107/722 | 0.36 | 0.17 | 2.09 | **0.036** | 1.43(1.02-1.99) | 0.62 | 0.61 | 0.63(0.57-0.68) | 0.87(0.83-0.92) |
| CHS_ZH | NSOC* | 3000/11179 | 0.23 | 0.022 | 10.28 | **8.64E-25** | 1.26(1.20-1.31) | 0.021 | 0.009 | 0.57(0.56-0.58) | 0.60(0.59-0.62) |
| | NSCLP* | 2020/11179 | 0.33 | 0.026 | 12.63 | **1.46E-36** | 1.39(1.32-1.47) | 0.039 | 0.018 | 0.61(0.59-0.62) | 0.66(0.65-0.68) |
| | NSCPO* | 980/11179 | 0.017 | 0.036 | 0.47 | 0.64 | 1.02(0.95-1.09) | 0.004 | 0.004 | 0.50(0.49-0.52) | 0.58(0.56-0.60) |

Note: NSOC, non-syndromic orofacial cleft cases by combining NSCLP,NSCLO and NSCPO together; NSCL/P, non-syndromic cleft lip with or without palate (NSCLP&NSCLO); NSCLP, non-syndromic cleft lip and palate; NSCLO, non-syndromic cleft lip only;CHW, Western Chinese; CHE, Eastern Chinese; CHS, Southern Chinese;* indicate without Sex in the model;OR, Odds ratio; 95%CI, 95% confidence interval;R2, the proportion of variance explained by the fixed predictors; w, with; wo, without; AUC, Area under curve;PGS, polygenic genetic score.
